# For Better or Worse? Subjective Expectations and Cost-Benefit Trade-Offs in Health Behavior

**DOI:** 10.1101/2023.05.14.23289957

**Authors:** Gabriella Conti, Pamela Giustinelli

**Author notes:** We thank Mirko De Maria, Dylan D’Mello, Mirjam Otten, Martina Rovetto, and Elena Ashtari Tafti for able research assistance. Gabriella Conti has received funding from the European Research Council (ERC) under the European Union’s Horizon 2020 research and innovation programme (grant agreement No. 819752 DEVORHBIOSHIP - ERC-2018COG). This project has received full ethics approval by the UCL IOE Research Ethics Committee (REC 1348 “Perceived Costs and Benefits of COVID-19 Social Distancing Measures and Compliance Behaviours”). We are grateful to Sue Horton for discussing the paper at the 2021 AEA/ASSA Annual Meeting, to Michael Darden for his discussion at the 10th ASHEcon Annual Conference, tow Arthur Juet for his discussion at the Empirical Health Economics Workshop (EHEW) 2022, to Shuye Yu for his discussion at the 29^th^ European Workshop on Econometrics and Health Economics (EWEHE), and to Kevin Thom for his discussion at the 2022 Annual Health Econometrics Workshop (AHEW). We also thank conference participants to the 2021 AEA/ASSA Annual Meeting, the 10^th^ ASHEcon Annual Conference, the ECB-NYFed Conference on Expectations surveys, the IAAE 2022 Conference, the RHUL 2022 Workshop on “Global health, environment and labour”, the Empirical Health Economics Workshop (EHEW) 2022, the EEA-ESEM Congress 2022, the 29th European Workshop on Econometrics and Health Economics and the 2022 Annual Health Econometrics Workshop, as well as seminar participants at the University of Glasgow, UCL Social Research Institute, IZA Bonn, University of Wisconsin- Madison, Bocconi University, University of Copenhagen, University of Oxford, the Autonomous University of Barcelona, the European University Institute, NOVA University of Lisbon, NYU Abu Dhabi and the University of Innsbruck.

## Abstract

We provide a framework to disentangle the role of preferences and beliefs in health behavior, and we apply it to compliance behavior during the acute phase of the COVID-19 pandemic. Using rich data on subjective expectations collected during the spring 2020 lockdown in the UK, we estimate a simple model of compliance behavior with uncertain costs and benefits, which we employ to quantify the utility trade-offs underlying compliance, to decompose group differences in compliance plans, and to compute the monetary compensation required for people to comply. We find that, on average, individuals assign the largest disutility to passing away from COVID-19 and being caught transgressing, and the largest utility to preserving their mental health. But we also document substantial heterogeneity in preferences and/or expectations by vulnerability status, gender, and other individual characteristics. In our data, both preferences and expectations matter for explaining gender differences in compliance, whereas compliance differences by vulnerability status are mainly driven by heterogeneity in preferences. We also investigate the relationship between own and others’ compliance. When others fail to comply and trust breaks down, individuals respond heterogeneously depending on their own circumstances and characteristics. When others around them comply less, those with higher risk tolerance and those without prior COVID-19 experience plan to comply less themselves, while the vulnerables plan to comply more. When a high-level public figure breaches the rules, supporters of the opposing political party plan to comply less. These findings emphasize the need for public health policies to account for heterogenous beliefs, preferences, and responses to others in citizens’ health behaviors.

*JEL Codes*: C25, C83, D84, I12, I18.

## 1 Introduction

Health behaviors are actions individuals take that affect their health: protective behaviors such as exercising, eating healthy food, and having regular checkups, or risky behaviors such as smoking, eating junk food, and engaging in risky sex. Irrespective of their risky or protective nature, most health behaviors can have both positive and negative consequences for the individual, generating trade-offs in choice. Because the costs and benefits of alternative courses of action are *ex ante* uncertain, individual choices crucially depend on decision makers’ expectations over choice consequences as well as on how they resolve the trade-offs between expected costs and benefits.

Unpacking the roles of subjective expectations and preferences over the consequences of alternative actions, both of which may vary across individuals with different characteristics and experiences, is key to understand people’s health behaviors and to inform policy. Expectations may be influenced by information or sensitization interventions as well as through monetary or nonmonetary incentive schemes. Preferences, on the other hand, may be less malleable, and policies targeting them more controversial. While these issues have been examined with reference to specific behaviors such as smoking (see e.g. the research by Frank Sloan and coauthors, summarized in Sloan, Smith and Taylor (2003)’s book), the role played by expectations and preferences in explaining variation in health investments has been relatively understudied.^1^ In this paper, we contribute to filling this gap by laying out a portable framework that we apply within the setting of the coronavirus pandemic.

During the acute phases of the pandemic, social distancing and the more extreme forms of self-isolation introduced by the public health authorities were the main actions through which people could (and were required to) protect themselves and others from infection and its health-harming consequences. Most existing cost-benefit analyses consider only the health benefits of lockdowns (in terms of lives saved) vis-à-vis the costs deriving from aggregate losses in economic activity (e.g., Robinson, Sullivan and Shogren (2021), Viscusi (2020), Thunström et al. (2020)). Distancing and isolation, however, are not without costs or risks for individuals’ wellbeing. Psychological and/or financial distress (Chiesa et al., 2021), job loss (Gupta et al., 2023), and deterioration of social, psychological, and/or physical wellbeing (O’Connell, Smith and Stroud, 2022) are just a few prominent examples of the risks of prolonged social distancing and isolation. Upon forming expectations for the costs (risks) and benefits (returns) of distancing – and even isolation – versus those of more lenient conducts, citizens were thus confronted with the challenging task of resolving the cost-benefit trade-offs of compliance. In the first part of the paper, we elicit a variety of such expectations for the risks and returns of alternative compliance conducts, and we estimate utility parameters reflecting how individuals resolve the cost-benefit tradeoffs underlying compliance decisions.

Individual compliance decisions may not only depend on perceived costs and benefits – and how people resolve the tradeoffs among them – but also on the compliance behavior of others. We investigate this in two ways in the second part of the paper. First, we study the effect of others’ compliance behavior on individuals’ own compliance by eliciting respondents’ compliance probabilities under alternative hypothetical scenarios about the compliance behavior of others living in the same local authority. Second, we study the effect of the behavior of a high-level public figure on individuals’ own behavior by means of a randomized sensitization intervention that exploits a ‘naturally occurring’ event (the “Cummings’ affair”), and we investigate how exposure to the treatment (a screen showing the timeline of Dominic Cummings’ violations of lockdown rules) affects respondents’ compliance probabilities.^2^

Hence, our work also contributes to the economic literature on COVID-19, which has been growing fast. For instance, Brodeur et al. (2021) provide the first review of the economics of COVID-19, with over 700 citations on Google Scholar; Blundell et al. (2021) provide a review on the COVID-19 crisis in the UK. Many studies have elicited and analyzed beliefs and expectations related to the Coronavirus and the COVID-19, including Adams-Prassl et al. (2020), Akesson et al. (2022), Altig et al. (2020), Aucejo et al. (2020), Belot et al. (2021, 2020), Briscese et al. (2023), Bordalo et al. (2022, 2020), Bruine de Bruin and Bennett (2020), Ciancio et al. (2020), Collis et al. (2022), Delavande, Bono and Holford (2021), Kröger, Bellemare and de Marcellis-Warin (2020), Bravo and Sanz (2022), Papageorge et al. (2021), Savadoria and Lauriola (2022), Smith et al. (2020), Wise et al. (2020), among others. Differently from these (and other) studies, we connect subjective expectations on Coronavirus- and lockdown-related risks to compliance decisions (plans) in a formal model of discrete choice, in order to quantify the role of expectations and preferences in determining compliance.

Discrete choice analyses may be based on actual choices (“revealed preferences” or “RP” for short), stated choices (“stated preferences” or “SP” for short),^3^ or combinations of theirs (“RP & SP”).^4^ In this paper, we estimate a discrete choice model of compliance behavior using survey-elicited choice probabilities, which may be thought of as a probabilistic form of SP data.^5^ We use choice probabilities since our survey elicitation occurs at a time preceding the actual decision or behavior. We therefore enable respondents to express any uncertainty they might perceive regarding their behavior in the next four weeks by reporting interior instead of corner probabilities. Because compliance (or lack thereof) to the lockdown rules is a decision with uncertain consequences, we also use choice-contingent subjective probabilities over choice consequences on the right-hand side of the model.^6^

Discrete choice analyses based on SP data collected via a laboratory or survey experiment are also referred to as Discrete Choice Experiments (DCEs).^7^ In the second part of the paper, we use approaches germane to DCEs to study the effect of others’ compliance behavior on respondents’ compliance plans via hypothetical scenarios, and the effect of the behavior of a high-level public figure on respondents’ compliance plans via a randomized sensitization treatment exploiting the timeline of the Cummings affair. Hence, our work also contributes to the health economics literature on DCEs (e.g., Clark et al. (2014), Soekhai et al. (2019) and Haghani, Bliemer and Hensher (2021) in general, and Li et al. (2021), Filipe et al. (2022) for COVID-19 applications).

More specifically, in this paper we study the determinants of compliance decisions during the first lockdown in the UK (Spring 2020), using a formal framework of decision-making under subjective risk where decision makers maximize subjective expected utility, combined with rich survey data on individuals’ subjective expectations and other measures we collected in May 2020 from an age-gender- ethnicity representative sample of 1,100+ UK-based respondents on Prolific Academics.^8^

On March 23*^rd^* 2020, later than most European countries, the UK entered a strict first lockdown which remained effective until early June. “Stay home” was the single most important message and rule citizens were given by the UK authorities during this phase, with varying specifics and bindingness across different categories of citizens. All individuals were asked to minimize the time spent outside their home and, when outside, were required to stay (at least) 2 meters apart from anyone who did not belong to their own household. Leaving home was only allowed for essential activities or specific reasons.^9^ Special categories of citizens such as the vulnerables or the infected were subject to the stricter restrictions implied, respectively, by shielding and self-isolation.^10^ Transgressors were liable to prosecution and the police was given the power to fully enforce the rules through monetary fines, dispersion of gatherings, and even arrests.^11^ At the same time, monetary compensation schemes were gradually introduced for the self-isolating on low income.^12^ No other rules on specific protective or preventive behaviors such as face covering were introduced at this stage. All this has implications for how we conceptualize compliance (or lack thereof) in this setting.

We view compliance to the lockdown rules as the outcome of a decision under uncertainty, where individuals must form expectations about the costs (risks) and benefits (returns) of alternative actions and use their preferences to resolve the trade-offs between expected costs and benefits. For instance, a person may understand that going out – vis-à-vis never leaving home (the government’s recommendation) – implies higher risk of contracting the virus and, if not motivated by one of the essential activities or reasons allowed by the lockdown rules, a positive risk of sanctions. Yet, the person might weigh these risks against those of locking herself up at home. In fact, individuals may differ in their risk perceptions and/or in how they resolve cost-benefit trade-offs. These sources of heterogeneity may result in varying propensities to comply or, among people behaving similarly, they may represent different reasons for doing so. For example, some people may be induced to stay at home by a high perceived risk of contracting the Coronavirus or developing COVID-19 once infected, while others may be motivated by the fear of putting their family at risk or by that of sanctions. Some people may instead decide to go out more than permitted for fear of losing their job or for a strong need of fresh air and exercising.^13^

We formalize these ideas in Section 2, after introducing the main rules and features of the first UK lockdown our framework aims to capture. In the model, individuals can choose among four actions or conducts over a period of four weeks. These are: never leave home (A1); strict compliance with the rules (A2); general compliance but with some discretion, or discretionary compliance (A3); complete unconcern with the rules (A4). A1 corresponds to the government’s recommendation (“Stay Home”); it captures strict compliance for specific groups^14^ and “literal” or “over-compliance” for the remaining majority. A2 corresponds to careful adherence to the rules, which limited the frequency with which people could leave their home and the reasons for doing so, and required a physical distance of at least 2 meters between people not belonging to the same household. A3 corresponds to general or rough compliance with the rules, but with the possibility of using discretion depending on the situation or activity. Finally, A4 represents non-compliance, whereby the person disregards the rules and carries on with her own life as much as possible.^15^

We let people’s subjective expected utility of each action depend on a rich set of consequences or outcomes, some of which capable of generating trade-offs in compliance, consistent with recurring public discourse during the lockdown. These include contracting the Coronavirus; being unable to find ICU space in the hospital, having developed severe COVID-19; passing away for the complications of COVID-19; infecting others living with the person; infecting others not living with the person; being caught transgressing; expected fine for transgressing; becoming unhappy or depressed; gaining weight or becoming unfit; experiencing a deterioration of relationships with family, friends, or close colleagues; losing one’s job (for workers)/falling behind with exams (for students); running out of money.

We analyze respondents’ subjective expectations for these and related outcomes in Section 3, after describing our survey and sample. Following the survey expectations literature in economics on which we build, we elicited respondents’ subjective expectations probabilistically on a 0-100 numerical scale of percent chance. Most expectations were directly asked conditionally on alternative compliance behaviors (A1-A4), which is the form needed to study compliance decisions. For example, the within-person difference in the subjective probability of becoming depressed if one follows the rules with some discretion (A3) vis-à-vis never leaves home (A1), may be interpreted as the person’s perceived return to *not* following the rules strictly over never leaving home. For people who assign a lower likelihood of becoming depressed to A3 than A1, this return consists of a reduced perceived personal risk of depression. The within-person difference in the subjective probability of contracting the Coronavirus following A3 versus A1 can be given a similar interpretation. For people who assign a higher likelihood of contracting the virus following A3 than A1, the return consists of an increased perceived risk of own infection. Under standard preferences, this would be a “negative return”; alternatively, it could be interpreted as a (positive) return to never leaving home (A1) vis-à-vis following the lockdown’s rules with some discretion (A3).

Compliance behaviors too were elicited probabilistically, in this case as subjective choice probabilities. At the end of the expectations battery, we asked respondents their subjective probability, between 0 and 100 percent, of behaving according to each of the four conducts A1-A4 over the next four weeks. Respondents who feel sure about their conduct over the following month can express their certainty by reporting corner probabilities of 0 or 100 percent. Respondents who are not completely sure about their conduct during the upcoming month, perhaps because they recognize that circumstances might change or new information unfold, can express their uncertainty by reporting an interior probability.

In the descriptive analysis of Section 3, we present the empirical distribution of all expectations in levels and of the perceived returns computed as within-individual differences in choice-contingent expectations across actions. We do so both in the overall sample and for subsamples of respondents’ with different characteristics and experiences. We find significant differences in compliance probabilities by gender and vulnerability status,^16^ which we further investigate in terms of differences in the underlying preferences and beliefs.

In Section 4, we explain how we implement the model econometrically and we present parameter estimates. Because the choice-contingent expectations over choice consequences entering decision makers’ subjective expected utility are observed, as they were measured in the survey, the remaining unknown parameters of the model to be estimated are those capturing decision makers’ preferences over such choice consequences, that is, how decision makers resolve the cost-benefit trade-offs of compliance. Following the approach of Blass, Lach and Manski (2010), Arcidiacono et al. (2020), and others, we use choice probabilities for actions A1-A4 as our measures of compliance behavior on the left-hand side of the model. Under specific assumptions on the distribution of the unobservable components of decision makers’ subjective expected utility (discussed below), elicited choice probabilities can be inverted, leading to a tractable linear form estimable by least squares or least absolute deviations.

We find the largest estimated disutilities of non-compliance to be the psychological cost of being caught transgressing and that of passing away due to the health complications of COVID-19; whereas the largest utility weight associated with the benefits of non-compliance is that associated with avoiding becoming unhappy or depressed, i.e., our mental health outcome. We find significant heterogeneity in utility parameters (Subsection 4.3). For example, men have a greater disutility from becoming physically unfit (associated with the costs of compliance), while women from suffering a deterioration of their relationships; vulnerable individuals appear to perceive fewer tradeoffs than non-vulnerable ones. We also document heterogeneity in perceived returns (Subsection 4.4). Males have, in general, lower perceived risks, while vulnerable individuals have significantly higher perceived risks and lower perceived returns of leaving home.

Having documented that compliance probabilities and the underlying preferences and expected returns vary by gender and vulnerability status, in Subsection 4.5 we decompose group differences in compliance into components attributable to variation in expectations versus preferences. We find that differences in compliance probabilities across genders are explained by both differences in expectations and preferences, whereas differences in preferences seem the main source of variation explaining differences in compliance probabilities between vulnerable individuals and others.

In Subsection 4.6, we compute the compensation required for people to be isolated, using an indifference condition based on the model. We find that approximately a quarter of the sample requires compensation to be indifferent between never leaving home and their optimal choice, with substantial heterogeneity in the amount required. Notably, our model-based compensation for low-income individuals aligns well with the amount provided by the government within the ‘Test and Trace Support Payment’ for people on low incomes who have to self-isolate.

Lastly, in Section 5, we study the effects of the compliance behavior of others. We first investigate the influence of those living in the respondent’s local authority (LA) on the respondent’s own compliance via hypothetical scenarios. We find that when others fail to comply and trust breaks down, individuals respond heterogeneously depending on their own circumstances and characteristics. For example, when others around them comply less, those with higher risk tolerance and those without prior COVID-19 experience plan to comply less themselves, while the vulnerables plan to comply more. We also study the effect of compliance behavior of a high-level public figure, Dominic Cummings, via the randomized sensitization intervention mentioned above. We find that a group of respondents, those supporting the Labour party, react to the treatment’s negative prompt by lowering their subjective probability of never leaving home (A1) and increasing that of discretionary compliance (A3). Those randomized to the Cummings treatment additionally show a higher persistence of discretionary compliance in the next 4 weeks, independently of their political inclination.

Section 6 concludes.

## 2 Institutional Setting and Analytic Framework

### 2.1 “Stay Home”: First Lockdown’s Rules in the UK

The UK entered a strict first lockdown with a TV announcement by Prime Minister Boris Johnson on March 23rd 2020, later than most European countries.^17^ This first lockdown remained effective until early June 2020, when the restrictions began to be gradually lifted, a few weeks after Johnson’s announcement of a conditional phased lifting plan which was aired on May 10th. A detailed timeline is displayed in Figure A1 of the Supplementary Appendix.

“Stay home” was the single most important message and rule citizens were given by the UK authorities during the first lockdown. All citizens were asked to minimize the time spent outside their home and, when outside, were required to stay (at least) 2 meters apart from anyone who did not belong to their own household. A “Stay home” message was texted to all registered mobile phones in the United Kingdom (Figure 1); the ubiquitous logo is shown in Figure 2. In practice, the stay-home rule was applied with varying specifics and bindingness across the following four main categories of citizens:

1. *Self-isolating* – Individuals testing positive to the Coronavirus or manifesting symptoms consistent with COVID-19.
2. *Vulnerables* – Individuals over 70 years of age and/or affected by specific health conditions (e.g., severe lung or heart conditions, certain types of cancer) and/or undergoing specific medical treatments (e.g., cancer treatments or medicine that weakens one’s immune system). This category also included pregnant women.
3. *Key workers* – People working in critical sectors (e.g., the National Health Service or NHS).
4. *Others* – Everyone else.

**Figure 1:**
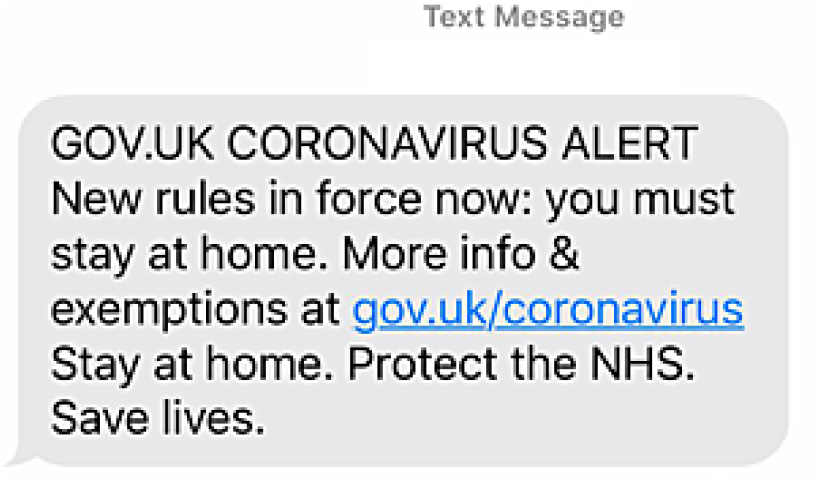
Coronavirus: SMS messages Notes: The government texted people across the UK to inform them of the new rules announced by the Prime Minister on 23 March 2020. This was made possible thanks to the cooperation of mobile phone operators.

**Figure 2:**
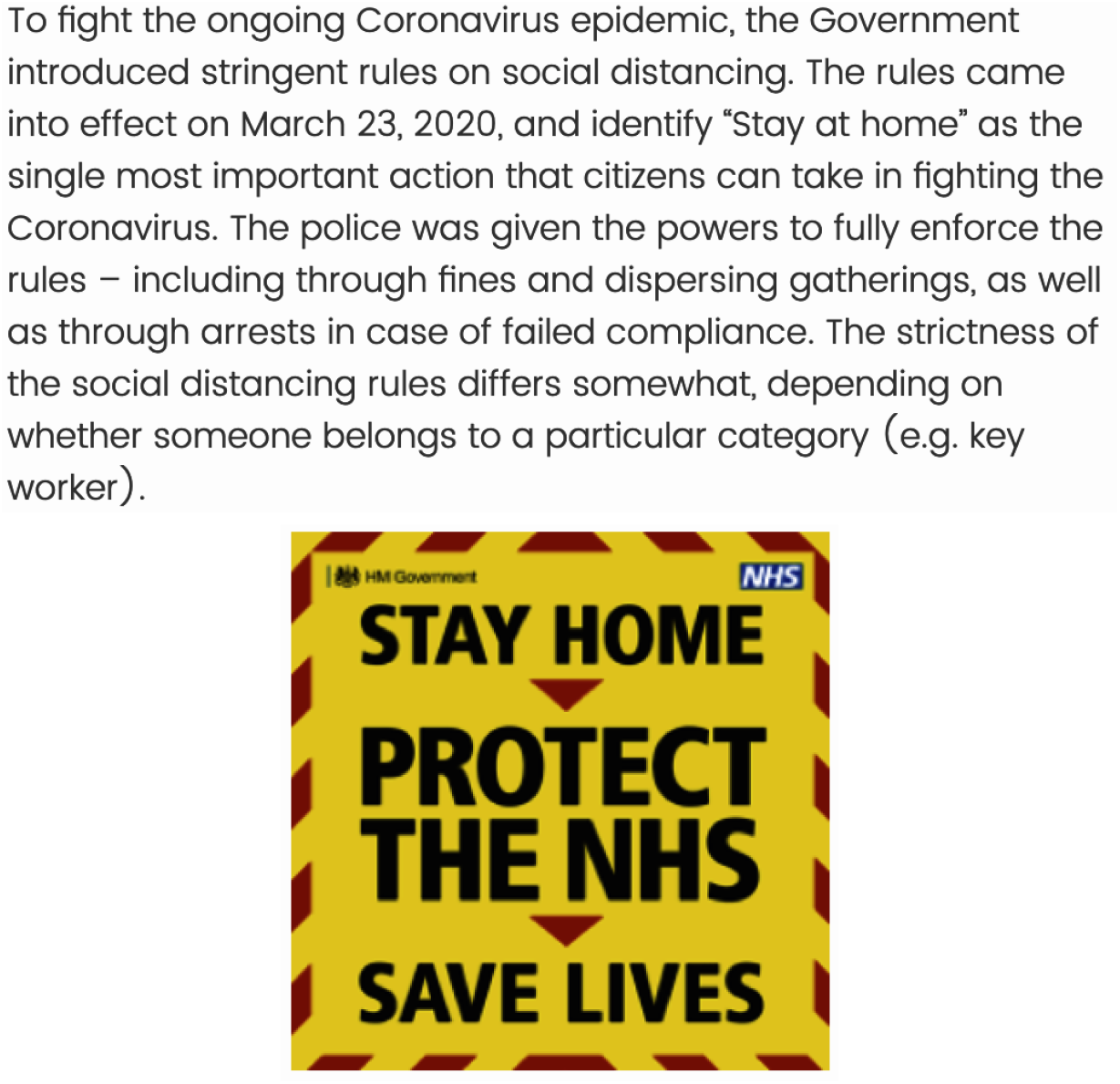
Introductory Screen of the Coronavirus Expectations Section

The first two groups were subject to the strictest rules. Self-isolating individuals were not allowed to leave their home for any reason for 7 consecutive days, self-isolating households for 14 consecutive days. Vulnerable individuals were expected not to leave their home for 12 consecutive weeks.^18^ The remaining groups were allowed to go out, but to a limited extent and only for the following specific reasons:

1. shopping for basic necessities (food and medicines);
2. one form of exercise per day (such as running, walking, or cycling), alone or with one’s household members;
3. any medical need (including donating blood or helping a vulnerable person);
4. attending the funeral of a close relative;
5. commuting to/from work, only for key workers and those who could not work from home;
6. taking children to/from school or childcare, only for key workers and parents of vulnerable children.

Transgressors were liable to prosecution and the police was given the power to fully enforce the rules through monetary fines, dispersion of gatherings, and even arrests. The fine schedule was £60 for the first penalty note (£30 if paid within 14 days); £120 for the second penalty note; doubled amount on each further repeat offence.

No additional rules on specific protective or preventive behaviors other than social distancing were given during this initial phase. Mandatory face covering in indoor settings, for instance, was not introduced until later in the Summer of 2020 (23 July 2020). All this has implications for how we think about compliance, including our modelling framework in Subsection 2.2 and our data collection and survey measures in Section 3.

To make things realistic but also keep them tractable, we allow (non-)compliance to take the form of one of four conducts of behavior or actions, the first two capturing compliance (including forms of over-compliance) and the other two non-compliance (partial or full).^19^ Specifically, we take the government’s “stay home” rule as the main benchmark. This was a binding rule for the vulnerables and self-isolating, and a strongly recommended behavior for everyone else. Accordingly, we define the status quo conduct or action as “Never leave home”. Key workers and other non-vulnerable individuals who were not self-isolating, (or after completing their quarantine period), were allowed to leave their home, albeit in a very restricted manner and only for the reasons specified by the lockdown rules. We define the conduct or action of individuals who closely follow the lockdown rules as “Strict compliance”. Some individuals may, occasionally or systematically, fail to comply with the rules. We define the behavior of those who keep the main rules in mind but apply them with some discretion, leading to occasional non-compliance, as “General compliance”. Finally, we define the behavior of those who carry out with their own life as much as possible without following the rules as “Non-compliance”.

### 2.2 Compliance Choice with Uncertain Consequences

We now introduce a modeling framework to analyze (non-)compliance with the rules just described. In Section 4, we give the framework econometric implementation and estimate model parameters using data on individuals’ subjective expectations for the consequences of alternative compliance conducts and for own compliance over the following month (early May-early June 2020), which we collected during the week of May 3-10 2020, right before Boris Johnson’s announcement of the conditional lifting plan. This and related data from the baseline survey are described in Section 3.

Citizens face a choice among a discrete set of J conducts or actions, *J* . The set *J* includes the four (non-)compliance behaviors described above (*J* = 4), with *j* = 1 denoting “Never leave home”, *j* = 2 denoting “Strict compliance”, *j* = 3 denoting “General compliance”, and *j* = 4 denoting “Non-compliance”. We assume that everyone can choose among the four choice alternatives, so the choice set is homogeneous across the population.

Individuals are forward looking, so their compliance decision depends on what they expect to result from it in the future. Each individual, denoted by *i*, derives utility *U_i_*(*θ⃗*), where 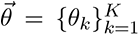 is a finite vector of consequences or outcomes. Because the elements of *θ⃗* are uncertain at the time of choice, the individual forms subjective probabilities, 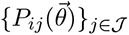, over the consequences of choosing each alternative, and then selects the SEU-maximizing alternative 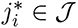,

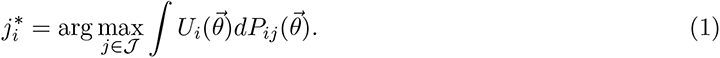

Following standard subjective expected utility theory, we specify (1) to be additively separable in the elements of *θ⃗* and, for each element of *θ⃗*, multiplicatively separable in the subjective probability and utility. Letting 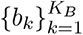 denote binary outcomes and 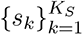 continuous ones, person i’s choice problem becomes:

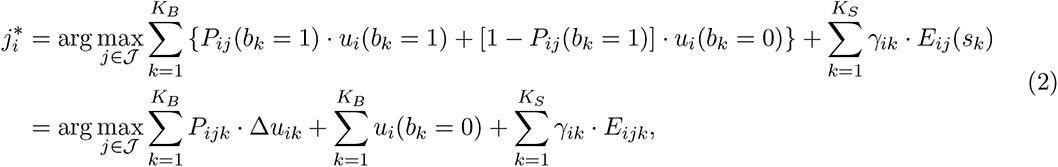

where *P_ijk_ ≡ P_ij_* (*b_k_* = 1) is i’s subjective probability that *b_k_* = 1 will result (e.g., *i* gets infected), if *j* is chosen; Δ*u_ik_* = *u_i_*(*b_k_* = 1) *− u_i_*(*b_k_* = 0) is the utility difference *i* derives from the occurrence of *b_k_* = 1 (e.g., *i* gets infected) relative to the occurrence of *b_k_* = 0 (e.g., *i* does not get infected); E*_ijk_ ≡* E*_ij_*(s*_k_*) is i’s subjective expectation for s*_k_* (e.g., monetary fine), if *j* is chosen; and *γ_ik_* represents the associated (dis)utility. Being constant across actions, the term 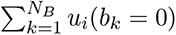 drops out of the choice.^20^,^21^

This specification allows the utility parameters 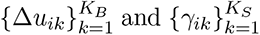 to vary across decision makers, but not across choice alternatives. The elements of 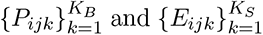, on the other hand, can vary unrestrictedly across individuals and alternatives.^22^ This modeling framework views compliance behavior as subjectively rational, in the sense that compliance decisions result from individuals maximizing their SEU. It does not imply, however, that individuals have correct or rational expectations over the consequences of alternative conducts of compliance. Put differently, our framework maintains that individuals make compliance decisions based on their expectations and utilities over the consequences of alternative conducts of compliance, but makes no assumptions about the rational or non-rational nature of decision makers’ expectations.

We specify i’s SEU in (2) as a function of the following probabilities (expectations):

- *k* = 1: Probability that i will contract the Coronavirus following *j*, *P_ij_*(Corona);
- *k* = 2: Probability that i will not find intensive care unit (ICU) space in the hospital while needing hospitalization due to the complications of COVID-19 following *j*, *P_i_*(no ICU space*|*acute COVID, Corona) *× P_i_*(acute COVID*|*Corona) *× P_ij_*(Corona);
- *k* = 3: Probability that i will die of COVID-19 following *j*, *P_i_*(dying of COVID*|*Corona)*× P_ij_*(Corona);
- *k* = 4: Probability that i will infect people with whom s/he lives following *j*, *P_ij_*(Infecting people living with) (for i’s living with others);
- *k* = 5: Probability that i will infect people s/he does not live with following *j*, *P_ij_*(Infecting people not living with);
- *k* = 6: Probability that i will be caught transgressing following *j*, *P_ij_*(caught);^23^
- *k* = 7: Expected monetary fine that i will receive if caught transgressing following *j*, E*_i_*(fine*|*caught) *× P_ij_*(caught);^24^
- *k* = 8: Probability that i will *not* become unhappy or depressed following *j*, 1 *− P_ij_*(Depressed);
- *k* = 9: Probability that i will *not* gain weight or become unfit following *j*, 1 *− P_ij_*(Gain weight/become unfit);
- *k* = 10: Probability that i’s relationships with family and close friends or colleagues will *not* deteriorate following *j*, 1 *− P_ij_*(Worse relationships);
- *k* = 11: Probability that i will *not* lose her job following *j*, 1 *− P_ij_*(Lose job) (for working i’s);
- *k* = 12: Probability that i will *not* fall behind with exams following *j*, 1*−P_ij_*(Fall behind with exams) (for studying i’s);
- *k* = 13: Probability that i will *not* run out of money following *j*, 1*−P_ij_*(Run out of £).

As explained shortly in Section 3, these probabilities are either directly elicited in the survey (those of outcomes 1, 4, 5, and 6), or are constructed from elicited ones (those of outcomes 2, 3, 7). For example, the probability of outcome k = 3 following action *j* is constructed by multiplying the probability of contracting the Coronavirus with or without symptoms following *j* by the probability of developing COVID-19 with acute symptoms and by the probability of passing away from COVID-19 conditional on contracting the Coronavirus, both of which were directly elicited by our survey. The elicited unconditional probabilities are listed in Section 3.4. The probability of each outcome k *∈* 8-13 is constructed as one minus the probability of the complement event, which was elicited in the survey. This choice of framing is made for communication. First, recall that in equation (2), Δ*u_ik_* represents the difference in utility that person *i* obtains from the occurrence of outcome k (*b_k_* = 1) relative to the occurrence of the complement outcome (*b_k_* = 0). For individuals with standard preferences, we expect Δ*u_ik_* to be negative for outcomes 1-7 (i.e., disutilities) and positive for outcomes 8-13 (i.e., utilities). In Section 4, we will estimate the model parameters, thus empirically testing these hypotheses.

Second, in the econometric implementation of Section 4, we take A1 (“never leave home”) as the reference action and we model choice of A2 (strict compliance), A3 (general compliance), or A4 (noncompliance) relative to A1 (the government’s benchmark). As a result, the relevant variables for choice of A2, A3, or A4 over A1 are the perceived returns to A2, A3, and A4 relative to A1.^25^ As argued by example in Section 1, for reasonable configurations of individuals’ expectations, the differences in the probabilities (or expectations) for outcomes 1-7 following actions A2, A3, and A4 versus action A1 are likely to capture increased perceived risks of negative outcomes (or “negative perceived returns”), whereas those for outcomes 8-13 are likely to represent increased perceived likelihoods of positive outcomes (or “positive perceived returns”). We analyze these perceived returns and the underlying subjective probabilities in Sections 3 and 4.

## 3 Survey Design and Data Description

### 3.1 Data Collection

To recruit our sample we use Prolific Academic, an online platform providing participants for web-based research, including web surveys.^26^ It is considered to be a source of high-quality participants and data (e.g., Peer et al. (2017)), superior to otherwise similar research platforms.^27^ Prolific Academic has been increasingly used for web-based research in economics and other social sciences, including for COVID-related research (e.g., Akesson et al. (2020), Buso et al. (2020), and Campos-Mercade et al. (2021)). A useful feature of Prolific Academic is the possibility of requesting age-gender-ethnicity representative samples for the UK and the US. For our study, we took advantage of this feature and requested an age-gender-ethnicity representative sample of the UK population during early May of 2020.^28^

We collected our data by means of two web surveys: a lengthier baseline, which we fielded on May 3-10, 2020, and a short follow-up, which we fielded on May 28, 2020. We describe each of them in turn.

#### 3.1.1 Baseline Survey

**Baseline overview.**The baseline survey is structured in five main sections, as follows:

A. *You and Your Health* – This section covers the respondent’s age, gender, self-rated health, health conditions and history, height and weight to construct BMI, including changes since the start of the pandemic.
B. *Corona Knowledge* – This section measures the respondent’s awareness of the situation (e.g., existence of the Coronavirus and related COVID-19 disease, lockdown status) and the respondent’s familiarity with various aspects of the ongoing pandemic (e.g., COVID-19 symptoms, protective behaviors, pandemic statistics, lockdown rules).
C. *Corona Experience* – This section asks questions about the respondent’s experience with the Coronavirus/COVID-19, in first-person and through family, friends, or acquaintances.
D. *Corona Behaviors* – This section elicits respondent’s habits during the lockdown (e.g., number of days the person went out, specific behaviors when outside, etc.).
E. *Corona Expectations* – This sections measures: (i) risk perceptions related to the Coronavirus over the following 4 weeks (e.g., unconditional probability of contracting the virus, of developing COVID-19 symptoms conditional on contracting the virus, etc.); (ii) perceptions of risks related to Coronavirus and risks related to the lockdown, under alternative scenarios regarding the respondent’s compliance behavior in the following 4 weeks (e.g., the probability of contracting the Coronavirus conditional on actions A1-A4 described above, the probability of developing depression conditional on actions A1-A4, etc.); (iii) the respondent’s subjective probability of following each of the four compliance conducts (A1-A4); (iv) the respondent’s probability of compliance under hypothetical scenarios of the compliance behavior of others living in his/her same local authority, and an estimate of the proportion (between 0% and 100%) of people living in their local authority who would follow each of the A1-A4 conducts in the following 4 weeks. This section includes also some questions eliciting the respondent’s familiarity with the concepts of chance and percent and the respondent’s interpretation of the non-compliance scenarios (A3-A4).
F. *Background Information* – This section covers additional demographics (e.g., marital status, parental status, and household structure); information on the respondent’s socioeconomic status (e.g., education, employment status, work mode, and income bracket, including changes since the start of the pandemic); IQ (via Raven’s matrices); time and risk preferences.

**Expectations battery.** After eliciting respondents’ knowledge, experience, and behavior with regard to various aspects of the pandemic and lockdown, we began the Corona Expectations section (E) with an introductory screen reporting basic information about the government’s lockdown rules. This includes the main “Stay Home” rule to protect the NHS and save lives, some information on enforcement, and a note on the fact that specifics may vary across categories of citizens: the introductory screen is reported in Figure 2; more detailed information on citizens’ categories and category-specific rules followed.^29^ Sharing basic information about the lockdown and its rules at the beginning of Section E was a deliberate design choice we made. Because the baseline was fielded about 5 weeks into the lockdown, we thought it important to gather data on respondents’ familiarity with main aspects of the pandemic and lockdown to learn if/to what extent it differed across respondents. Moreover, in order for the expectations questions and scenarios we posed in Section E to be understandable by and meaningful to everyone, we thought it important to create a common knowledge base on the main lockdown rules and on the meaning of compliance or lack thereof for different types of citizens.

All expectations for binary or discrete events of Section E were elicited using a visual 0-100 linear scale of percent chance, with a clickable slider to minimize response anchoring.^30^ This format has been also found to have desirable properties with respect to the use of “focal” responses (0, 50, 100)^31^ and rounding of reports.^32^ As an example, Figure 3 displays the survey screen with the question eliciting the percent chance of contracting the Coronavirus in the next 4 weeks.

**Figure 3:**
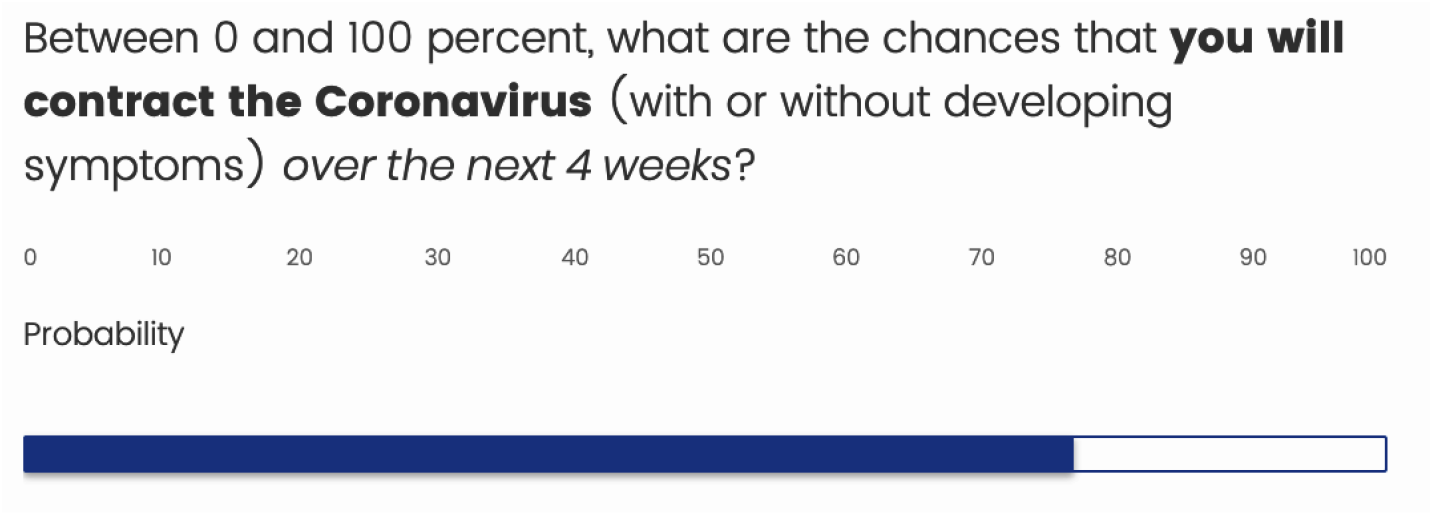
Example of Expectation Elicitation Question, Unconditional

#### 3.1.2 Follow-up Survey

Using the launch of the NHS Test and Trace Service (TTS) as pretense, we fielded a short follow-up survey on May 28th 2020, which we used to implement a randomized sensitization intervention based on the “Cummings scandal”. Designed to protect the NHS while helping the country return to a more normal life, the Test and Trace Service (TTS) was introduced on May 28th 2020 to trace the spread of the virus and isolate new infections, in an important monitoring and early-warning role both locally and nationally.^33^ Figure A2 shows the introductory screen of our follow-up survey, which includes the screenshot of the notice published by the Department of Health and Social Care on May 27th 2020.

By the time the TTS was launched, the Cummings scandal had just reached its peak. Figure A3 displays the “Cummings Screen”, which we used to implement a randomized sensitization intervention whereby treatment respondents were shown the screen at the beginning of the follow-up, whereas control respondents were shown the screen at the end of it.^34^ The screen goes over the main events of the Cummings affair, from Boris Johnson’s national lockdown announcement on March 23rd, followed by Dominic Cummings’ first alleged violation of the lockdown rules on March 27th, to the Downing Street rose garden press conference where Dominic Cummings defended his conduct as reasonable and legal.

After asking respondents to provide an assessment of whether Cummings had or not broken the rules,^35^ we re-elicited respondents’ citizen category, compliance probabilities over actions A1-A4 in the following 4 weeks, and the estimate of the proportion of people living in their LA who would follow each of the four compliance behaviors (from Section E of the baseline survey). Finally, we asked a new compliance probability question related to the TTS: specifically, we elicited respondents’ subjective probability that they will self-isolate (even with no symptoms) if the contact tracers (as part of the new NHS Test and Trace service) told them that they had been in contact with someone with the virus over the previous 14 days; we also asked them which factors (Including the Cummings affair) they considered in their answer.

### 3.2 Sample Description

Our sample consists of 1,100+ adults living in the UK on May 3-10, 2020. The sample is representative with respect to age, gender, and ethnicity.

Table 1 shows sample statistics for gender, age, ethnicity, education, household income, location, citizen category (i.e., vulnerable, self-isolating, and key worker), employment status, cohabitation status, prior experience with the Coronavirus/COVID-19, degree of literacy regarding the Coronavirus/COVID- 19, willingness to take risks, and patience.

**Table 1:**
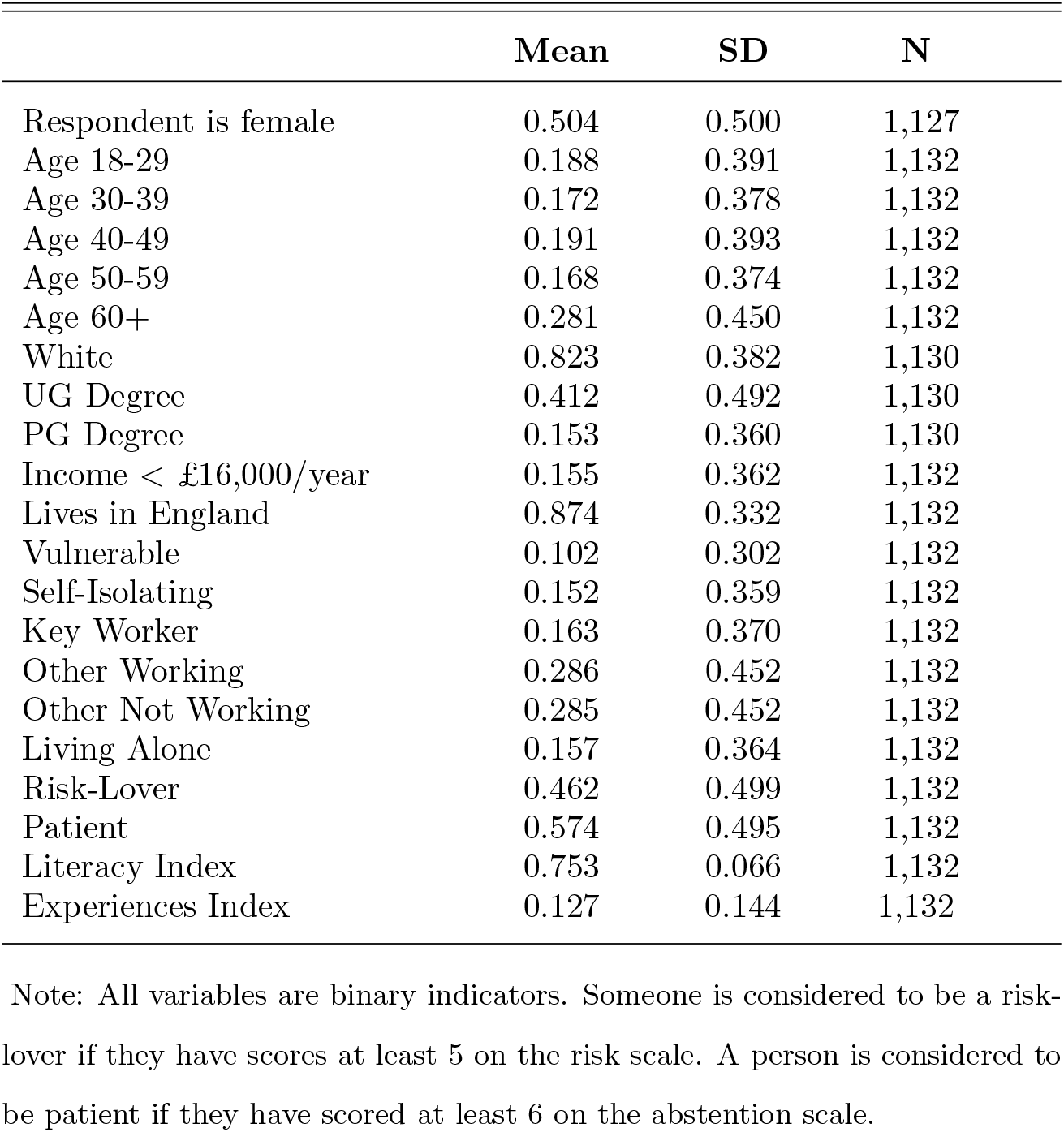
Summary Statistics

We confirm that the sample is representative by gender, age, and ethnicity. We have 41% of the respondents with at least an undergraduate degree (slightly more educated than the population), 15% of the respondents on low income (less than £16,000/year), 10% of the respondents self-identifying as belonging to the vulnerable category, 15% self-isolating, 16% key workers, an equal percentage (28.5%) of other working and other not working, and 16% of respondents living alone.

Using the responses to our questions in Section (C) of the survey (*Corona Experience*), we created a COVID-19 experience index, capturing the respondent’s prior experience with the Coronavirus/COVID- 19, obtained through personal experience or through that of family and/or friends.^36^ The index ranges between 0 and 1, where 0 means no prior experience with the Coronavirus/COVID-19 and 1 means that the respondent indicated having some form or degree of experience with the Coronavirus/COVID-

19 in all questions of the experience battery. The sample distribution of the COVID-19 experience index ranges from 0 to 0.762, with nearly 31% of respondents reporting no prior experience with the Coronavirus/COVID-19. The sample mean is 0.127 and the sample standard deviation is 0.144, indicating that at the time of the survey respondents had limited prior experience with the Coronavirus/COVID-19 on average, but also revealing substantial heterogeneity across respondents in their prior experience with the virus and related illness.

Similarly, we used the responses to our questions in Section (B) of the survey (*Corona Knowledge*) to create a COVID-19 literacy index, capturing the respondent’s degree of knowledge and familiarity with respect to the Coronavirus/COVID-19 pandemic.^37^ The index ranges between 0 and 1, where 0 means complete unawareness of the Coronavirus/COVID-19 pandemic and 1 means that the respondent displayed awareness about the ongoing pandemic and answered correctly all questions requiring familiarity with statistics regarding the virus, the related illness, and the lockdown rules. The sample distribution of the COVID-19 literacy index ranges from 0.492 to 0.915. The sample mean is 0.753 and the sample standard deviation is 0.066, indicating that at the time of the survey respondents had a relatively high degree of knowledge and familiarity with the pandemic on average, and that COVID-19 literacy is much less dispersed than COVID-19 experience in our sample.

Respondents’ willingness to take risks and their degree of patience were elicited using Falk et al. (2016)’s risk and patience scales. Essentially, respondents are asked to rate their willingness to take risks and their willingness to abstain from something today in order to afford more tomorrow on a 0-10 scale. 46% of respondents rate their willingness to take risks to be 5 or higher and 57% of respondents rate their patience 6 or higher. Hence, we constructed two binary indicators for willingness to take risks to be *≥*5 (‘High Willingness to Take Risks’) and willingness to abstain from something today to be *≥* 6 (‘High Patience’).

### 3.3 Compliance Plans As Subjective Choice Probabilities

We start from the end of the expectations section (E) and begin by describing respondents’ compliance plans for the upcoming month, which we elicited probabilistically as the respondent’s likelihood of following each compliance conduct or action in A1-A4. In Section 4, we use these data as left-hand-side variables in our model of compliance behavior.

#### Elicitation

As for all probabilistic expectations we elicited in our survey, compliance probabilities were measured on a 0-100 percent chance scale using clickable sliders. In this case, the question screen displayed four sliders, one for each of the four compliance conducts or actions, A1-A4. For each conduct or action, the respondent was asked to select on the corresponding slider a number between 0 percent and 100 percent reflecting the likelihood that she would follow that conduct over the next 4 weeks. At the bottom of the screen, the sum of the four responses was displayed to help the respondent select four probabilities summing to 100 percent. The question screen is shown in Figure 4.

**Figure 4:**
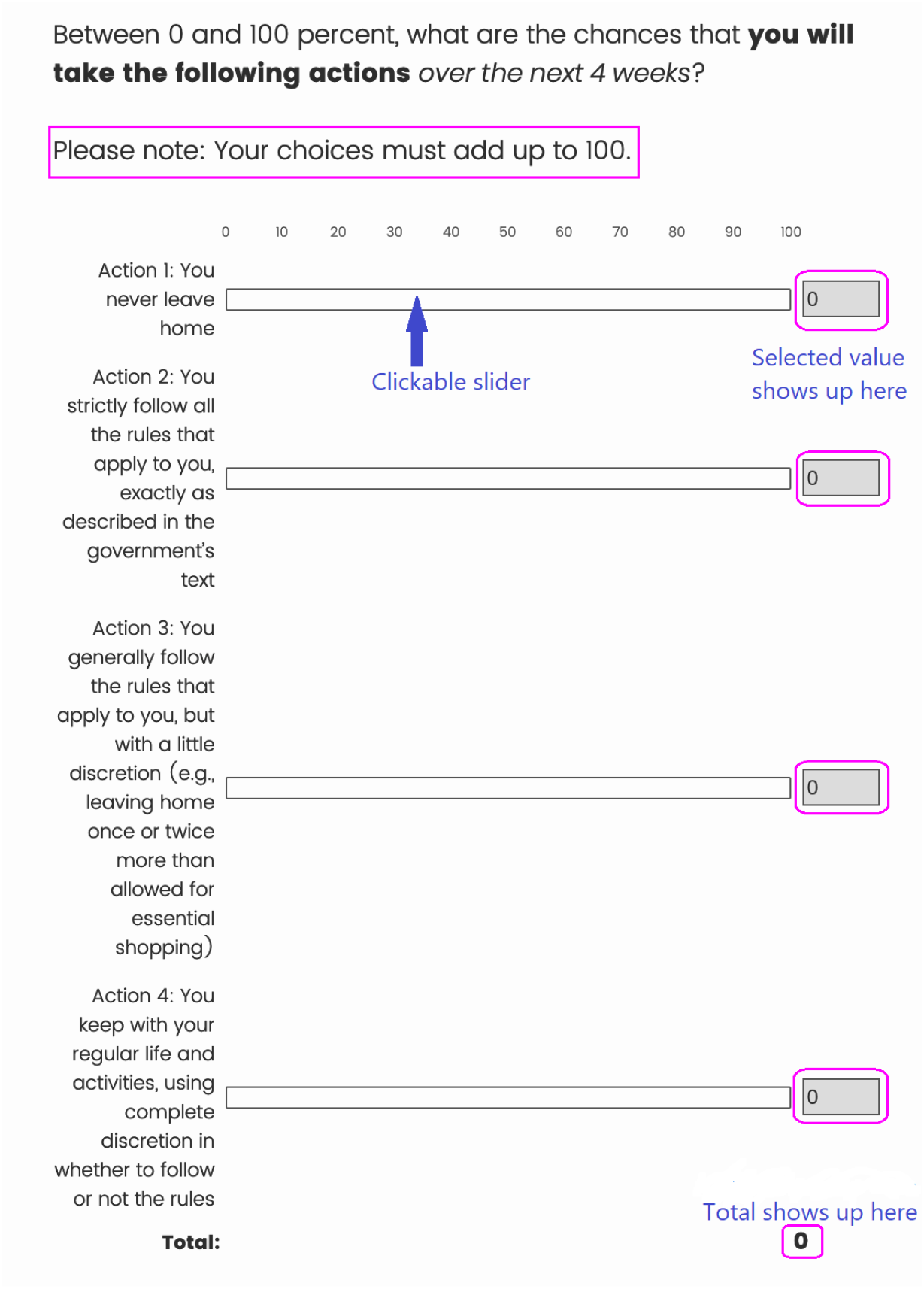
Elicitation of Choice Probabilities

#### Description

Table 2 reports various features of the empirical distribution of respondents’ compliance probabilities for each action in A1-A4. Complete histograms are shown in Appendix Figure A4. Strict compliance (A2) is the conduct with the highest probability of being chosen on average, with both mean and median around 54-55 percent. Never leave home (A1) and General compliance (A3) follow, with a mean of 22 percent for A1 and 19 percent for A3, and a median of 10 percent for both actions. The mean probability of Non-compliance (A4) is around 4 percent, the median 0 percent.

**Table 2:** Compliance Probabilities for Actions A1-A4

| Actions | min | p10 | p25 | p50 | p75 | p90 | max | mean | sd | N |
| --- | --- | --- | --- | --- | --- | --- | --- | --- | --- | --- |
| A1 - Never leave home | 0 | 0 | 0 | 10 | 38 | 75 | 100 | 22.25 | 29.39 | 1,132 |
| A2 - Strict compliance | 0 | 8 | 25 | 54.5 | 84.5 | 96 | 100 | 54.15 | 32.30 | 1,132 |
| A3 - General compliance | 0 | 0 | 0 | 10 | 28.5 | 55 | 100 | 19.31 | 24.37 | 1,132 |
| A4 - Non-compliance | 0 | 0 | 0 | 0 | 2 | 13 | 100 | 4.28 | 11.55 | 1,132 |

The statistics shown in Table 2 reveal substantial heterogeneity in respondents’ compliance probabilities, the extent of which varies between compliance and non-compliance actions. Specifically, for each of the four actions the smallest reported probability of choosing that action is 0 percent and the largest is 100 percent, spanning the whole 0-100 scale. However, the choice probability distributions for compliance actions A1-A2 have larger standard deviations (29 and 32 percent, respectively) than those of the choice probability distributions for non-compliance actions A3-A4 (24 and 12 percent, respectively).

The statistics of Table 2 represent cross-sectional distributions of marginal subjective choice probabilities. It is also of interest to describe person-specific patterns in choice probabilities across compliance actions. Around 11% of respondents displayed firm compliance plans by assigning the whole probability mass (100 percent) to one of the four actions: 3.18% to A1 (Never leave home), 6.45% to A2 (Strict compliance), 1.24% to A3 (General compliance), and 0.18% to A4 (Non-compliance). The remaining 89% of respondents displayed some uncertainty over their future compliance behavior by assigning a positive choice probability to two or more alternative conducts of compliance. In particular, nearly 33% of respondents split the whole probability mass between two actions, 28% between three actions, and 28% between all four actions.

If we take the lockdown rules in general and the government’s “Stay Home” guidance in particular as benchmarks, these statistics imply that only about 3% of respondents planned to never leave home for sure over the next month and about 28% of them planned to never leave home and/or strictly comply for sure, where the latter group includes the former. Over 72% of respondents assigned a strictly positive probability to either or both non-compliance conducts (A3-A4). However, only 2.2% of respondents assigned the whole probability mass to A3 and/or A4.

At this point, it is useful to discuss whether there might be social desirability bias in reports of compliance probabilities. While we don’t have mobility tracking apps linked to our respondents, we can provide some pieces of evidence which speak against such concerns. First of all, there is abundant evidence from multiple sources that, during the first lockdown, compliance with the COVID-19 public health measures was very high: existing sources report levels of compliance around 95% (e.g., Keyworth et al. (2021) and Ganslmeier, Van Parys and Vlandas (2022)).^38^ Second, such evidence is corroborated by the patterns observed in the mobility data based on GPS-powered devices such as smartphones, for example in the Google mobility data.^39^ Third, we further validate our elicited compliance plans with actual behavior elicited in the follow-up survey (see paragraph “Interpretation and validation” later in this section and related Table 4).^40^

#### Heterogeneity

As shown in Table 2, there is substantial heterogeneity in respondents’ compliance plans expressed as choice probabilities over alternative compliance conducts. It is therefore natural to ask whether and, if so, how compliance probabilities vary with respondents’ characteristics and circumstances. We begin by investigating whether compliance probabilities vary across categories of citizens, who faced lockdown rules of varying strictness. Table 3 shows the mean and standard deviation of the reported choice probabilities for each action, this time disaggregated by vulnerability status (left panel), gender (middle panel), and COVID-19 experience index (right panel).^41^

**Table 3:** Compliance Probabilities for Actions A1-A4, by Vulnerability, Gender, and Experience

| Vulnerables | mean | sd | N | Males | mean | sd | N | COVID-19<br>Exp. Index > 0 | mean | sd | N |
| --- | --- | --- | --- | --- | --- | --- | --- | --- | --- | --- | --- |
| A1 - Never leave home | 43.33 | 35.44 | 287 | A1 - Never leave home | 20.86 | 28.63 | 559 | A1 - Never leave home | 21.79 | 28.93 | 783 |
| A2 - Strict compliance | 34.85 | 32.29 | 287 | A2 - Strict compliance | 52.91 | 32.36 | 559 | A2 - Strict compliance | 54.06 | 31.84 | 783 |
| A3 - General compliance | 17.77 | 24.03 | 287 | A3 - General compliance | 20.93 | 24.86 | 559 | A3 - General compliance | 19.76 | 24.18 | 783 |
| A4 - Non-compliance | 4.05 | 11.56 | 287 | A4 - Non-compliance | 5.30 | 13.89 | 559 | A4 - Non-compliance | 4.39 | 11.92 | 783 |
| Non-Vulnerables | mean | sd | N | Females | mean | sd | N | COVID-19<br>Exp. Index = 0 | mean | sd | N |
| A1 - Never leave home | 17.95 | 25.67 | 845 | 1 - Never leave home | 23.57 | 30.02 | 568 | A1 - Never leave home | 23.28 | 30.43 | 349 |
| A2 - Strict compliance | 58.77 | 31.41 | 845 | 2 - Strict compliance | 55.50 | 32.18 | 568 | A2 - Strict compliance | 54.35 | 33.34 | 349 |
| A3 - General compliance | 19.54 | 24.50 | 845 | 3 - General compliance | 17.60 | 23.64 | 568 | A3 - General compliance | 18.32 | 24.81 | 349 |
| A4 - Non-compliance | 3.74 | 10.70 | 845 | 4 - Non-compliance | 3.33 | 8.61 | 568 | A4 - Non-compliance | 4.05 | 10.71 | 349 |

As expected, among the vulnerables the probability of action A1 (Never leave home) is substantially higher (mean = 43.33 percent) than among the other respondents (mean = 17.95 percent). The higher mean probability of action A1 among the vulnerables is almost exactly balanced by a lower (of approximately the same amount) probability of action A2 (Strict compliance). These differences in mean probabilities of actions A1 and A2 between vulnerables and non-vulnerables are statistically significant, and so are the differences in mean probabilities of actions A3 and A4 between males and females shown in the middle panel. Specifically, women report lower probabilities of non-compliance (A3-A4) than men on average.

Moving to the individual-level patterns of choice probabilities across compliance conducts or actions, a higher proportion of vulnerables assigns the whole probability mass to action A1 (Never leave home) than the corresponding percentage of non-vulnerables: 18.26% against 1.47%. Similarly, a higher percentage of vulnerables assigns the whole probability mass to A1 *and/or* A2 than the corresponding proportion of non-vulnerables: 50.44% against 25.17%.^42^ Yet, nearly 50% of vulnerables assign a strictly positive probability to either or both non-compliance actions (A3 and/or A4); the corresponding figure among the non-vulnerables is 75%. Finally, the percentages of vulnerables and non-vulnerables assigning the whole probability mass to A3 and/or A4 are similar: 3.48% among the vulnerables and 2.07% among the non-vulnerables. One may wonder how vulnerables choose between A1 and A2. To address this question, we compute the ratio between the probability of action 1 and the sum of the probabilities of action 1 and 2 for each respondent. This statistic has mean 0.5, which implies that, on average, vulnerables assign equal probability to either action. While the standard deviation of this ratio is quite large (0.35), we find that this heterogeneity is not explained by a variety of observable characteristics, including age, gender, education, time and risk preferences, knowledge and experience with coronavirus, and health.

The middle panel of Table 3 shows the mean and standard deviation of compliance probabilities by gender. Women (middle-bottom panel) report on average higher compliance probabilities than men (A1 mean = 24 vs. 21 percent and A2 mean = 56 vs. 53 percent) and lower non-compliance probabilities (A3 mean = 18 vs. 21 percent and A4 mean = 3 vs. 5 percent). In addition to having a higher mean, the distributions of A3 and A4 probabilities are also more dispersed among men than among women.

Finally, the right panel of Table 3 shows the mean and standard deviation of compliance probabilities by COVID-19 experience index (no experience in the right-bottom panel vs. any amount of experience in the right-top panel). The distributions are very similar between the two groups, although respondents with no prior experience with COVID-19 do report slightly higher compliance probabilities (A1-A2) and slightly lower non-compliance probabilities (A3-A4) than the other respondents on average.

To sum up, there are statistically significant differences in choice probabilities by vulnerability status and gender, but not by prior experience with COVID-19.

#### Interpretation and validation

Conduct A1 (Never leave home) is objectively defined and should have the same meaning for everyone. Conduct A2 (Strict compliance with the lockdown rules) is also objectively defined, as long as one is familiar with the rules and with how those vary across citizens. Since we had our respondents familiarize themselves with the main lockdown rules at the beginning of Section E, we assume that everyone understood the meaning and implications of A2 for them when they answered the compliance expectations questions and the other expectations questions contingent on compliance. The meaning of A3 and A4, on the other hand, might vary somewhat across respondents. To understand how respondents interpreted non-compliance and learn what they thought about when we asked them to hypothetically entertain the two non-compliance scenarios A3 and A4, towards the end of the expectations section we asked them the following open question: “*What non-compliance behaviour did you think about?* ”. The bar graph shown in Figure A5 of the Appendix reports the most named activities mentioned by respondents. In order from the 1*^st^* to the 5*^th^* most mentioned, they include: (i) visiting relatives, (ii) exercising more than once a day, (iii) meeting with friends, (iv) sunbathing, and (v) going to the second house. Figure A6 of the Appendix shows a snippet of quotes from a sample of respondents.

At the end of the expectations section, we additionally asked respondents to rate on a scale between 0 and 100 their understanding of the concept of chance and their familiarity with percent chance scale. Reassuringly, the mean and median ratings are quite high (78 and 83, respectively). The full histogram of self-rated familiarity is shown in Figure A7 of the Appendix.

Finally, we take advantage of specific questions we fielded within the Cummings follow-up survey on May 28th 2020, asking respondents their habits during the previous week, to construct measures of compliance behavior that can be compared to the compliance probabilities we elicited at baseline and used to estimate the discrete choice model of compliance behavior. Specifically, we first asked respondents how many days they stayed at home all day long in the past 7 days. To those who reported that they stayed at home less than 7 days, we then asked to think about the day/s on which they went outside and to indicate the number of days they engaged in a series of behaviors.^43^ We used the (4) Worn disposable gloves (5) Avoided public transportation (6) Worn a face mask (surgical/N95/FFP2/FFP3) (7) Worn responses to these questions to create two dummy variables for each individual: the first dummy, called stay-home dummy, is equal to 1 if the individual reported staying at home all day long for the entire past week, and is equal to 0 otherwise; the second dummy, called no-compliance dummy, is equal to 1 if the individual reported engaging in at least one behavior that was in violation of the lockdown rules, and is equal to 0 otherwise. In Table 4, we report the estimates of a linear probability model regressing the stay-home dummy on the baseline probability of staying at home, or action A1 (top panel), and those of a linear probability model regressing the no-compliance dummy on the baseline probability of non-compliance, or action A4 (bottom panel). These regressions are estimated in the panel sample, linking individuals’ responses to the baseline survey (week of 03-10 May 2020) with their responses to the Cummings follow-up (28 May 2020), and are shown for the full sample as well as separately by gender, vulnerability status, and prior experience with the COVID-19.^44^

**Table 4:** Validation of Compliance Probabilities

|  | Dependent Variable: Stay-Home Dummy |  |  |  |  |  |  |
| --- | --- | --- | --- | --- | --- | --- | --- |
| | Full Sample | Male | Female | Vulnerable | Non-Vulnerable | COVID-19<br>Exp ≠ 0 | COVID-19<br>Exp = 0 |
| **Prob(A1)** | 0.536***<br>(0.033) | 0.527***<br>(0.047) | 0.536***<br>(0.046) | 0.683***<br>(0.097) | 0.471***<br>(0.036) | 0.564***<br>(0.038) | 0.479***<br>(0.062) |
| **Constant** | 0.022*<br>(0.012) | 0.022<br>(0.017) | 0.022<br>(0.018) | 0.047<br>(0.057) | 0.025***<br>(0.012) | 0.007<br>(0.014) | 0.053**<br>(0.024) |
| **t-stat**<br>**slope=1** | -14.27 | -10.08 | -10.14 | -3.27 | -14.86 | -11.53 | -8.44 |
| **F-stat const=0**<br>**& slope=1** | 138.75 | 67.31 | 71.59 | 8.10 | 145.88 | 98.64 | 71.15 |
| **N** | 1,041 | 516 | 520 | 105 | 936 | 712 | 329 |
| **R<sup>2</sup>** | 0.208 | 0.197 | 0.209 | 0.325 | 0.158 | 0.238 | 0.156 |
|  | Dependent Variable: Non-Compliance Dummy |  |  |  |  |  |  |
| | Full Sample | Male | Female | Vulnerable | Non-Vulnerable | COVID-19<br>Exp ≠ 0 | COVID-19<br>Exp = 0 |
| **Prob(A4)** | 0.267**<br>(0.117) | 0.265*<br>(0.140) | 0.311<br>(0.220) | -<br>- | 0.298**<br>(0.126) | 0.310**<br>(0.136) | 0.142<br>(0.232) |
| **Constant** | 0.693***<br>(0.015) | 0.679***<br>(0.021) | 0.705***<br>(0.020) | -<br>- | 0.658***<br>(0.016) | 0.697***<br>(0.017) | 0.682***<br>(0.027) |
| **t-stat**<br>**slope=1** | -6.25 | -5.24 | -3.13 | - | -5.57 | -5.09 | -3.69 |
| **F-stat const=0**<br>**& slope=1** | 1213.33 | 553.98 | 651.96 | - | 927.15 | 868.94 | 345.51 |
| **N** | 1,131 | 558 | 568 | - | 1,016 | 782 | 349 |
| **R<sup>2</sup>** | 0.005 | 0.006 | 0.004 | - | 0.006 | 0.007 | 0.001 |

Notes. The two dependent variables are measured at follow-up (28 May 2020). Stay-Home Dummy=1 if the respondent reports to have never left the house in the past week. Non-compliance Dummy = 1 if the number of transgressions performed in the last week is non-zero. The regressors are measured at the baseline (w/c 3 May 2020). Prob(A1) = subjective probability of never leaving home in the next four weeks. Prob(A4) = subjective probability of non-complying with the rules in the next four weeks.
The vulnerable column in the bottom half is blank because there is no vulnerable person for which the dummy of non-compliance is equal to 1. Standard errors in parenthesis. \*\*\*: $p < 0.01$ ; \*\*: $p < 0.05$ ; \*: $p < 0.1$ .

Before inspecting the estimates, it is important to keep in mind that the comparison between the choice probabilities elicited at baseline and the self-reported behavior elicited in the Cummings follow-up is less than ideal. First, the choice probabilities were asked with reference to a 4-week horizon, whereas the behavior is reported with reference to the third week following the baseline survey (week before 28 May 2020). Second, the realized behavior was not elicited in terms of the same compliance and non-compliance categories used to elicit respondents’ choice probabilities at baseline. This is especially true of the no-compliance dummy, which likely underestimates the incidence of non-compliance in the particular week for which the question was asked and, hence, also in the four weeks which the compliance probability questions refer to. There are at least two reasons why this might be the case: first, the questions used to construct the no-compliance dummy cover a limited set of behaviors the individual might have engaged in; second, the baseline survey was fielded right before Johnson’s announcement (on May 10, 2020) of the gradual lifting of the lockdown starting in June, wheres the follow-up survey was fielded after the announcement (on May 28, 2020). Therefore, by the end of May citizens likely had a more relaxed approach in anticipation of the phasing out of the lockdown. Indeed, by comparing the subjective compliance probabilities between waves (Figure A8), we find that the percentage of those reporting to never leave home almost halved, from 22.3% to 13.3%; most of the mass moved to A2, as the percentage of those reporting to comply strictly increased from 54.2% to 60.7%. The percentage of those planning general compliance remained unchanged (19.5%), while the percentage of those planning non-compliance to the rules slightly increased, from 4.28% to 6.35%. We find a similar pattern in the respondents’ perceptions of compliance behavior of others living in their local authority (Figure A9).

We estimate the regressions shown in Table 4 in the spirit of a validation exercise; however, for the reasons just explained, the estimates should be interpreted with caution. Perhaps unsurprisingly, the subjective probability of staying at home (A1) is a better predictor of actually staying at home in the top panel of Table 4 than the subjective probability of not complying (A4) is of the no-compliance dummy in the bottom panel of the table. In the top panel (stay-home regression), the estimated slope coefficient ranges between 0.471 and 0.683, and it is strongly statistically significantly different from 0. It is also statistically different from 1, which would be the theoretical value expected under rational expectations. The estimated constant is usually not statistically different from 0, which is the theoretical value expected under rational expectations (with no aggregate shocks). Vulnerable respondents are better than non-vulnerables at predicting staying at home, both looking at the slope coefficient (*p*=0.012) and at the *R*^2^; however, the point estimates are not significantly different across the other two groups (gender and COVID-19 experience). In the bottom panel (no-compliance regression), the estimated slope coefficient ranges between 0.142 and 0.311. It is usually significantly different from 0, but not in all groups. It is clearly significantly smaller than 1. Also, the estimated constants are all significantly different from 0.

### 3.4 Perceived Coronavirus and Related Risks, Not Conditioned on Compliance

We now turn to the other questions of Section E, eliciting respondents’ perceptions of Coronavirus-related risks. We begin with the unconditional expectations and then continue with those contingent on alternative compliance conducts in the next subsection.

#### Elicitation

We asked unconditional expectations for four binary or discrete events and one continuous outcome, as follows.

1. The percent chance that the person will contract the Coronavirus (with or without symptoms) over the next 4 weeks.
2. The percent chance that the person would develop No symptoms/ At most mild symptoms/ Severe-to-acute symptoms of the disease (COVID-19) that require hospitalization, if she were to contract the Coronavirus over the next 4 weeks.^45^
3. The percent chance that the person would be able find space in a hospital with Intensive Care Unit (ICU), if she were to contract the Coronavirus and develop severe-to-acute symptoms of the disease (COVID-19) that require hospitalization with intensive care.
4. The percent chance that the COVID-19 would be fatal for the person, if she were to contract the Coronavirus.
5. The amount the person expects to be fined in GBP, if caught transgressing.

#### Description

Main features of the sample distributions of these expectations are reported in Table 5. On average, respondents assigned a probability of 25 percent to the event of contracting the Coronavirus during the month of May 2020 (median 20 percent). Conditional on hypothetically contracting the Coronavirus, they assigned a mean probability of 31 percent to the event of not developing COVID- 19 symptoms, 44 percent to the event of developing COVID-19 with at most mild symptoms, and 25 percent to the event of developing COVID-19 with severe-to-acute symptoms requiring hospitalization. (The corresponding median probabilities are 25, 42, and 18 percent.) Conditional on hypothetically contracting the Coronavirus and developing severe-to-acute COVID-19 symptoms requiring intensive care hospitalization, respondents assigned a probability of 29 percent to the possibility of finding space in a hospital with ICU (median 20 percent). Finally, conditional on hypothetically contracting the Coronavirus, the mean probability of passing away due to the complications of COVID-19 is 29 percent (median 20 percent). As for respondents’ expectations over the monetary sanctions for transgressing the lockdown rules, the mean expected fine is £136.5 (median £61).

**Table 5:** Perceived Coronavirus and Related Risks, not Conditioned on Compliance

|  | min | p10 | p25 | p50 | p75 | p90 | max | mean | sd | N |
| --- | --- | --- | --- | --- | --- | --- | --- | --- | --- | --- |
| PC of contracting Coronavirus | 0 | 3 | 9 | 20 | 40 | 51 | 100 | 24.89 | 21.07 | 1,132 |
| PC of developing no symptoms,<br>if contract Coronavirus | 0 | 5 | 11 | 25 | 47.5 | 64 | 100 | 30.88 | 22.76 | 1,132 |
| PC of developing mild symptoms<br>if contract Coronavirus | 0 | 18 | 30 | 42 | 60 | 73 | 100 | 43.91 | 20.69 | 1,132 |
| PC of developing severe symptoms<br>if contract Coronavirus | 0 | 3 | 9 | 18 | 35.5 | 60 | 100 | 25.21 | 23.08 | 1,132 |
| PC of not finding space in ICU, if contract<br>Coronavirus and severe symptoms | 0 | 0 | 7 | 20 | 49 | 71 | 100 | 29.15 | 27.16 | 1,132 |
| PC of passing away, if contract Coronavirus | 0 | 3 | 8 | 20 | 50 | 70 | 100 | 29.48 | 25.76 | 1,132 |
| Expected fine (GBP), if caught | 0 | 44 | 60 | 61 | 123.5 | 301 | 1,000 | 136.5 | 178.1 | 1,132 |

Note: PC = Percent Chance.

Further inspection of Table 5 reveals substantial heterogeneity in reported expectations around the means. The empirical distributions of subjective probabilities range between 0 and 100 percent for all Coronavirus-related risks. The empirical distribution of the expected fine, conditional on being caught transgressing, ranges between £0 and £1,000. The empirical standard deviation ranges between 21 and 27 percent for the subjective probabilities and equals £178 for the expected fine. (The difference between 90th and 10th percentile ranges between 48 and 71 percent for the subjective probabilities and equals £257 for the expected fine.)

### 3.5 Perceived Risks and Benefits of Non-Compliance, As Choice-Conditioned Subjective Probabilities

We now describe the expectations that we elicited conditional on alternative compliance conducts.

**Elicitation.** We elicited expectations for four Coronavirus-related events or risks, under four alternative and mutually exclusive hypothetical scenarios corresponding to each of the four compliance conducts or actions A1-A4. The expectations are:

1. The percent chance that the person would contract the Coronavirus (with or without symptoms) over the next 4 weeks.
2. The percent chance that the person would infect with Coronavirus someone living with her over the next 4 weeks.
3. The percent chance that the person would infect with Coronavirus someone *not* living with her over the next 4 weeks.
4. The percent chance that the person would be caught transgressing over the next 4 weeks.^46^

To illustrate, Figure A10 shows the survey screen with the question eliciting the subjective probability of contracting the Coronavirus over the next 4 weeks under the four compliance conducts. The display is similar to that for elicitation of compliance probabilities, with the difference that in this case the four probabilities do not need to some up to one.

We additionally elicited expectations for five events or outcomes, capturing aspects of a person’s wellbeing in different domains (personal health, personal relationships, work/study, and personal finances). As for the Coronavirus-related risks, each of these questions was asked under four alternative and mutually exclusive hypothetical scenarios corresponding to each of the four compliance conducts or actions A1-A4. The expectations are:

1. The percent chance that the person would become unhappy or depressed over the next 4 weeks.
2. The percent chance that the person would gain weight or become unfit over the next 4 weeks.
3. The percent chance that the person’s relationship with family, friends, and/or colleagues would deteriorate over the next 4 weeks.
4. The percent chance that the person would lose her job (for working respondents) or fall behind with exams (for students) over the next 4 weeks.
5. The percent chance that the person (and her family) would run out of money over the next 4 weeks.

For the latter set of outcomes, moving forward we work with the probabilities of the complement events. As previously explained, this is done for interpretation. In particular, we tend to think of the first set of outcomes as Coronavirus-related risks, which are likely higher under non-compliance than under compliance, thus capturing the perceived costs or risks of non-compliance. On the other hand, we tend to think of the second set of outcomes as other kinds of risks, which are likely lower under non-compliance than under compliance, thus capturing the perceived benefits or returns to non-compliance.

#### Description

Tables 6 and 7 report means and standard deviations (the latter in parenthesis under the means) of the empirical distributions of these expectations (columns 1-4) and of their differences within respondent and between pairs of compliance conducts (columns 5-7). All differences are taken with respect to conduct A1 (Never leave home), which we use as a reference action since it was the status quo behavior recommended by the authorities. Table 6 refers to the first set of events or outcomes, which we interpret as the costs or risks of non-compliance; Table 7 refers to the second set of events or outcomes, which we interpret as the benefits of, or returns to, non-compliance.

**Table 6:** Perceived Risks of Non-Compliance, As Choice-Conditioned Subjective Probabilities

|  | Never out<br>home<br>(A1) | Strict<br>compl.<br>(A2) | General<br>compl.<br>(A3) | Non-<br>compl.<br>(A4) | A2-A1 | A3-A1 | A4-A1 |
| --- | --- | --- | --- | --- | --- | --- | --- |
| PC of contracting Coronavirus over next month | 10.14<br>(18.65) | 19.61<br>(23.39) | 27.74<br>(21.15) | 54.35<br>(28.72) | 9.47<br>(17.81) | 17.60<br>(22.35) | 44.21<br>(35.71) |
| PC of being unable to find ICU with acute COVID | 0.86<br>(2.85) | 1.64<br>(4.22) | 2.29<br>(4.53) | 4.19<br>(7.38) | 0.78<br>(3.12) | 1.43<br>(4.12) | 3.34<br>(7.15) |
| PC of passing away for COVID | 3.62<br>(8.77) | 6.42<br>(11.35) | 9.16<br>(12.31) | 17.21<br>(19.68) | 2.79<br>(7.65) | 5.54<br>(11.41) | 13.59<br>(19.71) |
| PC of infecting someone living with you over next month | 7.95<br>(17.98) | 15.38<br>(21.65) | 26.96<br>(22.69) | 52.56<br>(31.65) | 7.43<br>(15.94) | 19.01<br>(22.12) | 44.62<br>(35.48) |
| PC of infecting someone not living with you over next month | 4.71<br>(15.50) | 11.78<br>(19.51) | 22.32<br>(21.11) | 47.07<br>(30.83) | 7.07<br>(14.89) | 17.62<br>(21.62) | 42.36<br>(34.75) |
| PC of being caught transgressing | 0 | 0 | 15.31<br>(20.08) | 38.10<br>(31.56) | 0 | 15.31<br>(20.08) | 38.10<br>(31.56) |
| Expected fine if caught transgressing | 0 | 0 | 21.89<br>(54.83) | 51.17<br>(88.82) | 0 | 21.89<br>(54.83) | 51.17<br>(88.82) |

Note: PC=Percent Chance. N=1,132. Means and standard deviations (in parentheses). The last three columns display means of within-person differences.

**Table 7:** Perceived Returns to Non-Compliance, As Choice-Conditioned Subjective Probabilities

|  | Never out<br>home<br>(A1) | Strict<br>compl.<br>(A2) | General<br>compl.<br>(A3) | Non-<br>compl.<br>(A4) | A2-A1 | A3-A1 | A4-A1 |
| --- | --- | --- | --- | --- | --- | --- | --- |
| PC of not becoming unhappy or depressed over next month | 52.50<br>(34.63) | 62.90<br>(30.46) | 68.78<br>(26.08) | 73.90<br>(26.90) | 10.39<br>(20.44) | 16.28<br>(26.15) | 21.39<br>(36.30) |
| PC of not gaining weight or becoming unfit over next month | 48.33<br>(34.41) | 61.16<br>(30.39) | 67.33<br>(27.13) | 77.80<br>(22.78) | 12.82<br>(22.08) | 19.00<br>(25.42) | 29.47<br>(33.03) |
| PC of relationship not deteriorating over next month | 74.45<br>(30.58) | 77.49<br>(27.31) | 78.21<br>(24.35) | 74.03<br>(29.82) | 3.04<br>(14.02) | 3.76<br>(21.84) | -0.428<br>(37.48) |
| PC of not losing job | 75.83<br>(34.18) | 83.37<br>(26.84) | 84.03<br>(24.89) | 84.08<br>(24.89) | 7.54<br>(23.95) | 8.19<br>(26.37) | 8.24<br>(31.75) |
| PC of not running behind with exams | 69.33<br>(33.61) | 71.87<br>(28.70) | 72.43<br>(23.14) | 71.91<br>(27.88) | 2.55<br>(14.91) | 3.10<br>(20.49) | 2.58<br>(30.46) |
| PC of not running out of money over the next month | 81.27<br>(30.50) | 83.97<br>(26.92) | 85.12<br>(24.89) | 86.26<br>(23.64) | 2.71<br>(17.17) | 3.86<br>(19.38) | 5.00<br>(25.74) |

Note: PC=Percent Chance. N=1,132. Means and standard deviations (in parentheses). The last three columns display means of within-person differences.

There are clear gradients of subjective expectations across compliance conducts. In Table 6, all Coronavirus-related probabilities as well as the expected fine increase on average from left to right across the first four columns, that is, from Never leave home (A1) to Non-compliance (A4). The last three columns of Table 6 display the sample mean and standard deviation of the within-respondent differences in subjective expectations across pairs of actions, where A1 is always used as reference action. The mean difference is always positive (higher perceived Coronavirus-related risks for actions A2, A3, and A4 relative to A1) and increasing across actions (higher for increasing degrees of non-compliance). These statistics are also shown graphically in Figure A11 of the Appendix.

In Table 7 the probabilities of *not* experiencing negative outcomes increase on average from left to right across the first four columns, that is, from A1 to A4, only for the personal health domain. Instead, among the set of outcomes for personal relationships and personal finances, the average gradients look quite modest. Since 1st of March 2020, the government had introduced the Coronavirus Job Retention Scheme in an effort to help employers avoid the need to make mass redundancies as a result of the impact of COVID-19.^47^ This should have lessened individuals’ perceived risks of losing their job and of running out of money.^48^ The last three columns display the sample mean and standard deviation of the within-respondent differences in subjective probabilities across pairs of actions, which display a similar pattern. These statistics are also shown graphically in Figure A12 of the Appendix.

## 4 Econometric Implementation and Estimation Results

### 4.1 Econometric Implementation

*At the time of choice*, the decision problem of person *i* has the form

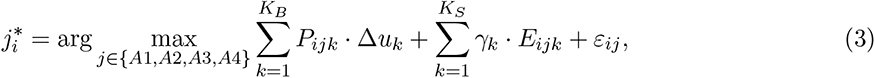

where compared to (2), we have suppressed the subscript *i* from the utility parameters, now assumed homogeneous across individuals,^49^ and have introduced an additive term, ε*_ij_*, capturing components of the decision maker’s SEU that are unobserved by the econometrician.^50^ Under the standard revealed preference argument, whereby each person’s compliance behavior coincides with her SEU-maximizing conduct or action,

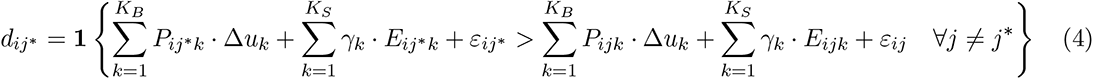

where **1** {*·*} is the indicator function and d*_ij_*= 0 *∀j ̸*= *j^∗^*, observation of 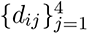 in a population or sample of individuals along with the individuals’ subjective expectations over the uncertain consequences of alternative compliance conducts, 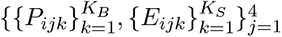, enables identification of the unknown utility parameters, 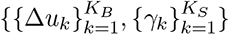, given assumptions on the distribution of unobservables, 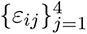 across the population.

Recall, however, that our survey elicits respondents’ compliance plans for the next 4 weeks in the form of choice probabilities. *At the time of the survey*, the decision problem of person *i* has the form

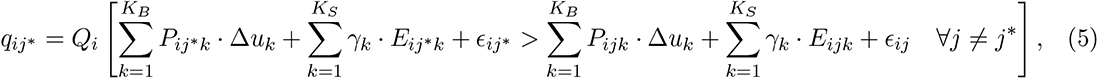

where q*_ij_∗* is person i’s subjective probability of choosing action *j^∗^* in the next 4 weeks and where, for simplicity, no explicit notation for time is used. The right-hand side of equation (5) provides a subjective random utility interpretation of elicited choice probabilities, 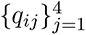, which are probabilistic versions of stated choices. It says that person *i* holds subjective probability q*_ij_∗* that following compliance conduct *j^∗^* will be optimal over the next 4 weeks, in the sense that it will yield a higher SEU than any of the other feasible compliance conducts.

The term ɛ*_ij_* in problem (5) has a partially different interpretation from that of term ε*_ij_* in problem (3)-(4). ɛ*_ij_* may be thought of as a composite term, ɛ*_ij_* = ϑ*_ij_* +ξ*_ij_*, where ϑ*_ij_* captures factors unknown to the econometrician but known to the decision maker (like ε*_ij_* in (3)-(4)), whereas ξ*_ij_* represents factors unknown to *both* the decision maker and the econometrician. In the taxonomy of Manski (1999) (see also Blass, Lach and Manski (2010) and Arcidiacono et al. (2020)), ξ*_ij_* represents *resolvable uncertainty*. As such, it captures all those factors that are unknown to person *i* when she is asked to make predictions 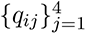, but would be known to her in the actual choice situation, that is, when she decides what compliance conduct to follow.

Person i’s subjective distribution Q*_i_* over 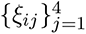 expresses the person’s resolvable uncertainty on her hypothetical or future optimal action. It is however important to note that if a person perceives no resolvable uncertainty about her optimal course of action at the time she is asked to predict compliance, she can express her certainty by reporting corner probabilities equal to 1 (for the optimal conduct she is certain to follow) or 0 (for the remaining conducts). Hence, eliciting choice probabilities is more informative than asking stated choices.

To implement (5) econometrically and provide estimates of the model parameters, we follow Arcidiacono et al. (2020) and assume that the components of 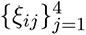 are i.i.d. Type 1 Extreme Value according to both the econometrician and decision maker. Under this assumption, the choice probabilities, 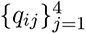, have the familiar exponential closed form

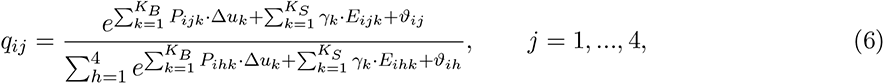

and are therefore invertible by applying the natural logarithm to each side of (6).^51^ Thus, applying the log-odds transformation to (6) yields the following linear specification:

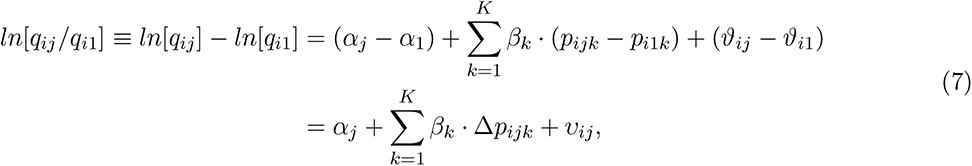

where “Never leave home” is the reference action (j = 1 or A1); the alternative-specific constant for A1 is normalized to 0 (α_1_ = 0); β*_k_* denotes a generic element of the vector of utility parameters, 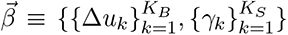, to be estimated; Δ*P_ijk_* denotes a generic element of the vector of person i’s perceived returns (or risks) of choosing each of actions A2, A3, or A4 over the reference action A1, 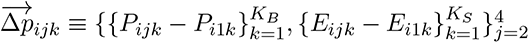.

We estimate the parameters of (7) by least squares, using data on respondents’ subjective probabilities over choices on the left-hand side and on respondents’ subjective probabilities (expectations) over choice consequences on the right-hand side. That is, to estimate the basic specification with homogeneous utility parameters, we use the data 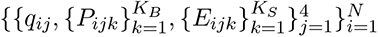, where N denotes sample size.^52^

### 4.2 Basic Specification With Homogeneous Utility Parameters

Table 8 presents least squares (LS) estimates of model (7). The first five coefficients represent the (dis-)utility weights attached to the corresponding Coronavirus-related risks, which we view as costs or risks of non-compliance. The sixth coefficient captures the nonmonetary (psychological) cost of being fined and the seventh the monetary one. These, too, may be viewed as costs or risks of non-compliance, since there are no sanctions, monetary or otherwise, for staying at home or strictly complying to the lockdown rules. The last six coefficients represent the utility weights attached to the corresponding health, relationship, and financial outcomes, which we view as benefits or returns of non-compliance.

**Table 8:** Model With Homogeneous Utilities – LS Estimates

| $\beta_k$ | Exp. Sign | Estimate |
| --- | --- | --- |
| **Risks** |  |  |
| $\beta_1$ (contracting Coronavirus) | - | 0.557 (0.468) |
| $\beta_2$ (unable to find ICU with acute COVID) | - | -1.129 (2.063) |
| $\beta_3$ (passing away for COVID) | - | -2.005 (0.934)** |
| $\beta_4$ (infecting people living with) | - | -0.899 (0.420)** |
| $\beta_5$ (infecting people not living with) | - | -1.419 (0.521)*** |
| $\beta_6$ (being caught transgressing) | - | -3.408 (0.362)*** |
| $\beta_7$ (expected fine) | - | -0.003 (0.001)** |
| **Benefits** |  |  |
| $\beta_8$ (not unhappy/depressed) | + | 1.618 (0.327)*** |
| $\beta_9$ (not unfit/gain weight) | + | 0.409 (0.359) |
| $\beta_{10}$ (no worse relationships) | + | 0.232 (0.316) |
| $\beta_{11}$ (not losing job) | + | 1.130 (0.459)** |
| $\beta_{12}$ (not running behind with exams) | + | 0.703 (1.331) |
| $\beta_{13}$ (not running out of money) | + | -0.688 (0.513) |
| Constant |  | 0.816 (0.126)*** |
| Observations |  | 1,132 |

Note: k=1: subjective probability of contracting Coronavirus; k=2: subjective probability of not finding space in ICU after contracting Corona & getting COVID-19 with severe symptoms; k=3: subjective probability of dying after contracting Coronavirus; k=4: subjective probability of infecting someone living with you; k=5: subjective probability of infecting someone not living with you; k=6: subjective probability of being caught transgressing; k=7: expected fine (weighted by subjective probability of being caught transgressing); k=8: subjective probability of not becoming unhappy/depressed; k=9: subjective probability of not gaining weight/becoming unfit; k=10: subjective probability of relationship not deteriorating; k=11: subjective probability of not losing job; k=12: subjective probability of not falling behind with exams; k=13: subjective probability of not running out of money. Standard errors clustered at the individual level in parentheses.
\*\*\*: $p < 0.01$ ; \*\*: $p < 0.05$ ; \*: $p < 0.1$ .

As expected, Coronavirus-related risks have negative associated utility weights. The only exception is the utility coefficient of contracting the Coronavirus with or without symptoms (*β*_1_), whose estimate is positive but statistically insignificant. A possible interpretation is that there is no disutility from contracting the Coronavirus per se, that is, beyond the disutility associated with its health-harming consequences, which are captured by the other risks included in the utility specification.^53^

The largest estimated disutilities are those from being caught transgressing (*β*_6_)^54^ and from passing away due to the health complications of COVID-19 (*β*_3_), followed by those from infecting non-cohabiting and cohabiting others (*β*_5_ and *β*_4_).^55^ The smallest albeit statistically significant coefficient among the first group is that multiplying the expected fine (*β*_7_). The estimated disutility of not being able to find the needed ICU space (*β*_2_) is sizable in magnitude but not statistically significant.

Moving to the benefits, the largest utility weight is *β*_8_, associated with avoiding becoming unhappy or depressed, the mental health outcome.^56^ Its estimate is of a similar magnitude (in absolute value) to the estimated disutility of infecting non-cohabiting others (*β*_5_). The utility weight associated with not losing one’s job (*β*_11_) follows; the coefficient’s magnitude is similar to the disutility of not finding ICU space, but its estimate is statistically significant.^57^ The utilities associated to the remaining outcomes are smaller in magnitude and statistically insignificant.

As a robustness check, Table A1 in the Supplementary Appendix reports least absolute deviations (LAD) estimates of model (7). The LAD estimates are very similar to the LS ones in both signs and magnitudes, but display some differences in estimation precision. For example, the disutility of passing away due to the complications of COVID-19 is no longer statistically significant in Table A1. On the other hand, the utility of avoiding becoming unfit/gaining weight, that of avoiding a deterioration of personal relationships and that of avoiding running out of money are now statistically significant.

Taken together, these estimates reveal the existence of trade-offs underlying compliance decisions and provide a first quantification of them. It is of course possible that individuals with different characteristics or circumstances use different sets of utility weights to resolve the trade-offs underlying compliance decisions. We investigate this aspect next.

### 4.3 Investigating Heterogeneity in Utilities

In Table 9, we re-estimate the model by allowing the utility parameters to vary by gender, vulnerability status, and prior experience with COVID-19.^58^ Each preference parameter Δ*u_k_*, where k indexes the outcomes listed by row (k *∈* 1*−*13), is modelled as *β*_const_+*β*_male_**1***_i_ {*male*}*+*β*_vulnerable_**1***_i_ {*vulnerable*}*+*β*_COVID-19 experience > 0_**1***_i_ {*COVID-19 experience > 0*}*, where **1***_i_ {*male*}* equals 1 if respondent *i* is male and 0 otherwise; **1***_i_ {*vulnerable*}* equals 1 if respondent *i* is vulnerable and 0 otherwise; and **1***_i_ {*COVID-19 experience > 0*}* equals 1 if respondent *i* has prior experience with COVID-19 and 0 otherwise. Thus, the estimates shown in the first column of Table 9 refer to the utility coefficients of the reference group, corresponding to non-vulnerable female respondents without prior COVID-19 experience. The estimates shown in the following columns represent the utility parameters of the remaining groups, corresponding to seven possible gender-vulnerability-COVID-19 experience combinations described in the columns’ labels.

**Table 9:** Model With Heterogeneous Utilities by Gender, Vulnerability, and COVID-19 Experience – LS Estimates

| $\beta_k$ | Female<br>Non-Vulnerable<br>No COVID Exp. | Female<br>Non-Vulnerable<br>w/ COVID Exp. | Female<br>Vulnerable<br>No COVID Exp. | Female<br>Vulnerable<br>w/ COVID Exp. | Male<br>Non-Vulnerable<br>No COVID Exp. | Male<br>Non-Vulnerable<br>w/ COVID Exp. | Male<br>Vulnerable<br>No COVID Exp. | Male<br>Vulnerable<br>w/ COVID Exp. |
| --- | --- | --- | --- | --- | --- | --- | --- | --- |
| **Risks** |  |  |  |  |  |  |  |  |
| $\beta_1$ (contracting Coronavirus) | -1.112<br>(1.086) | 0.197<br>(0.653) | -5.145**<br>(2.21) | -3.836*<br>(2.131) | 0.262<br>(0.876) | 1.571*<br>(0.818) | -3.772*<br>(2.155) | -2.462<br>(2.194) |
| $\beta_2$ (unable to find ICU with acute COVID) | -4.097<br>(3.958) | 0.271<br>(3.441) | -0.693<br>(5.129) | 3.675<br>(3.991) | -4.303<br>(4.047) | 0.065<br>(3.918) | -0.899<br>(5.964) | 3.469<br>(5.290) |
| $\beta_3$ (passing away for COVID) | 1.735<br>(2.057) | -1.695<br>(1.316) | 6.248**<br>(3.018) | 2.818<br>(2.669) | 0.078<br>(1.846) | -3.353**<br>(1.611) | 4.591<br>(3.027) | 1.160<br>(2.978) |
| $\beta_4$ (infecting people living with) | -0.388<br>(0.981) | -0.422<br>(0.585) | -4.044**<br>(1.701) | -4.077***<br>(1.566) | -0.433<br>(0.890) | -0.467<br>(0.703) | -4.089**<br>(1.779) | -4.122**<br>(1.746) |
| $\beta_5$ (infecting people not living with) | -2.264<br>(1.534) | -1.925***<br>(0.648) | 1.345<br>(2.150) | 1.683<br>(1.773) | -2.205*<br>(1.191) | -1.866**<br>(0.893) | 1.404<br>(2.086) | 1.742<br>(2.046) |
| $\beta_6$ (being caught transgressing) | -3.638***<br>(0.784) | -2.885***<br>(0.500) | -4.930***<br>(1.519) | -4.178***<br>(1.446) | -4.037***<br>(0.806) | -3.284***<br>(0.669) | -5.329***<br>(1.503) | -4.576***<br>(1.483) |
| $\beta_7$ (expected fine) | 0.0008<br>(0.002) | -0.003<br>(0.002) | -0.001<br>(0.005) | -0.005<br>(0.005) | 0.0002<br>(0.002) | -0.003<br>(0.002) | -0.002<br>(0.005) | -0.005<br>(0.005) |
| **Benefits** |  |  |  |  |  |  |  |  |
| $\beta_8$ (not unhappy/depressed) | 1.346**<br>(0.639) | 1.742***<br>(0.496) | -0.151<br>(1.332) | 0.245<br>(1.316) | 1.606**<br>(0.669) | 2.002***<br>(0.544) | 0.109<br>(1.338) | 0.505<br>(1.326) |
| $\beta_9$ (not unfit/gain weight) | -0.080<br>(0.853) | -0.273<br>(0.525) | 0.867<br>(1.116) | 0.674<br>(1.173) | 0.726<br>(0.803) | 0.533<br>(0.573) | 1.673<br>(1.271) | 1.480<br>(1.372) |
| $\beta_{10}$ (no worse relationships) | 0.587<br>(0.821) | -0.095<br>(0.489) | 3.065**<br>(1.349) | 2.382**<br>(1.175) | 0.474<br>(0.740) | -0.209<br>(0.513) | 2.951**<br>(1.278) | 2.268*<br>(1.161) |
| $\beta_{11}$ (not losing job) | 4.289***<br>(1.193) | 1.589***<br>(0.593) | 1.387<br>(3.058) | -1.314<br>(3.034) | 2.533**<br>(1.103) | -0.169<br>(0.711) | -0.369<br>(3.000) | -3.071<br>(3.062) |
| $\beta_{12}$ (not running behind with exams) | -3.369**<br>(1.442) | -1.455<br>(2.127) | -4.873<br>(5.409) | -2.960<br>(4.854) | 1.024<br>(2.677) | 2.937*<br>(1.513) | -0.481<br>(6.587) | 1.433<br>(5.511) |
| $\beta_{13}$ (not running out of money) | -2.743**<br>(1.189) | -0.568<br>(0.698) | -2.521<br>(2.615) | -0.346<br>(2.568) | -2.356**<br>(1.022) | -0.181<br>(0.893) | -2.134<br>(2.674) | 0.041<br>(2.754) |
| Constant | 0.883***<br>(0.123) |  |  |  |  |  |  |  |
| **Observations** | 1,127 |  |  |  |  |  |  |  |
| **Pseudo R<sup>2</sup>** | 0.182 |  |  |  |  |  |  |  |

Note: k=1: subjective probability of contracting Coronavirus; k=2: subjective probability of not finding space in ICU after contracting Corona & getting COVID-19 with severe symptoms; k=3: subjective probability of dying after contracting Coronavirus; k=4: subjective probability of infecting someone living with you; k=5: subj prob of infecting someone not living with you; k=6: subjective probability of being caught transgressing; k=7: expected fine (weighted by subjective probability of being caught transgressing); k=8: subjective probability of not becoming unhappy/depressed; k=9: subjective probability of not gaining weight/becoming unfit; k=10: subjective probability of relationship not deteriorating; k=11: subjective probability of not losing job; k=12: subjective probability of not running behind with exams; k=13: subjective probability of not running out of money. Standard errors in parentheses. \*\*\*: $p < 0.01$ ; \*\*: $p < 0.05$ ; \*: $p < 0.1$ .

The estimates in Table 9 provide evidence of heterogeneity in utility parameters by person’s characteristics and circumstances. In terms of risks, vulnerable respondents have large in magnitude and statistically significant disutilities of contracting the Coronavirus and of infecting people they live with, whereas nonvulnerable respondents have a larger and statistically significant disutility of infecting people they do not live with; also, male nonvulnerable respondents with prior COVID-19 experience have a larger disutility of passing away for COVID-19. In terms of benefits, vulnerable respondents have a larger utility of avoiding deterioration of relationships, while nonvulnerable ones have a larger utility of becoming unhappy/depressed, avoiding losing their job, and running behind with exams.^59^

In addition to being a function of utilities, choice probabilities are also a function of perceived returns to (non-)compliance, which too may vary across individuals. In the next two subsections, we investigate the predictors of individuals’ perceived returns to (non-)compliance and we decompose observed group differences in choice probabilities between a component explained by variation in utilities and a component explained by variation in expectations.

### 4.4 Investigating Heterogeneity in Expectations

In Table 10, we estimate best linear predictors of perceived risks of noncompliance (top panel) and perceived returns to noncompliance (bottom panel) conditional on gender, vulnerability status, and prior experience with COVID-19. Perceived risks and returns are defined and constructed as individual-level differences in subjective probabilities over choice consequences across pairs of compliance actions. For each consequence, k = 1, …, 13, the differences in the corresponding choice-contingent probabilities are computed relative to the benchmark conduct *j* = 1 or A1 (Never leave home). Each column corresponds to a separate perceived risk or return: for each of them, the probability differences across pairs of actions (A4-A1, A3-A1, A2-A1) are pooled.

**Table 10:** Heterogeneity in Perceived Risks and Returns by Gender, Vulnerability, and COVID-19 Experience

| height | Subjective probability of [consequence $k$] if Action $j$ vs Action 1 (= Never Leave Home) | | | | | | |
| --- | --- | --- | --- | --- | --- | --- | --- |
|  | (1)<br>Contracting<br>Coronavirus | (2)<br>No ICU with<br>acute COVID | (3)<br>Passing away<br>for COVID | (4)<br>Infecting People<br>Living With | (5)<br>Infecting People<br>Not Living With | (6)<br>Being Caught<br>Transgressing | (7)<br>Expected<br>Fine |
| **Male** | -0.040***<br>(0.011) | -0.004**<br>(0.002) | -0.018***<br>(0.005) | -0.049***<br>(0.012) | -0.044***<br>(0.010) | -0.015<br>(0.009) | -1.718<br>(2.215) |
| **Vulnerable** | -0.026<br>(0.017) | 0.015***<br>(0.003) | 0.061***<br>(0.008) | 0.010<br>(0.019) | -0.024<br>(0.017) | -0.026*<br>(0.015) | -0.801<br>(3.636) |
| **COVID-19**<br>**Exp. Index > 0** | 0.032***<br>(0.011) | -0.0009<br>(0.002) | 0.005<br>(0.005) | 0.047***<br>(0.013) | 0.047***<br>(0.011) | 0.032***<br>(0.009) | 4.744**<br>(2.394) |
|  | (8)<br>Not Unhappy<br>Or Depressed | (9)<br>Not Unfit<br>or Gain Weight | (10)<br>No Worse<br>Relationships | (11)<br>Not Losing<br>Job | (12)<br>Not Running<br>Behind With<br>Exams | (13)<br>Not Running<br>Out of<br>Money |  |
| **Male** | 0.014<br>(0.009) | -0.0005<br>(0.009) | 0.032***<br>(0.009) | -0.020<br>(0.012) | 0.043*<br>(0.025) | -0.001<br>(0.007) |  |
| **Vulnerable** | -0.104***<br>(0.016) | -1.119***<br>(0.016) | -0.093***<br>(0.015) | -0.071***<br>(0.027) | 0.005<br>(0.067) | -0.041***<br>(0.012) |  |
| **COVID-19**<br>**Exp. Index > 0** | 0.015<br>(0.011) | 0.039***<br>(0.010) | -0.025**<br>(0.009) | 0.022<br>(0.014) | 0.078**<br>(0.031) | 0.003<br>(0.008) |  |

Note: Each column reports the estimated coefficients from a separate regression of an outcome on the variables listed in the first column. The outcomes in columns (1)-(7) at the top are the subjective probabilities of the risks in the top row if Action $j$ versus Action 1 (=Never Leave Home) is chosen, while the outcomes in columns (8)-(13) are the subjective probabilities of the returns in the bottom row if Action $j$ versus Action 1 (=Never Leave Home) is chosen. Standard errors clustered at the individual level in parentheses. \*\*\*: $p < 0.01$ ; \*\*: $p < 0.05$ ; \*: $p < 0.1$ .

We find evidence of significant heterogeneity in perceived risks and returns by gender, vulnerability status, and prior experience with COVID-19. For example, vulnerable respondents have higher perceived risks of not finding ICU space with acute COVID and of passing away from COVID associated to leaving home (A2-A4) versus staying home (A1), a lower perceived risk of being caught transgressing, and lower perceived returns to noncompliance for nearly all consequences. Male respondents have lower perceived risks in general (i.e., for all consequences), associated to leaving versus staying home, and higher perceived returns of avoiding deterioration of relationships. Respondents with prior COVID-19 experience have higher perceived risks (for nearly all consequences), associated to leaving home, and selected higher perceived returns (e.g., avoid becoming unfit/gaining weight and losing job) or lower ones (e.g., avoid relationships deterioration).

### 4.5 Decomposing Group Differences in Choice Probabilities: Expectations versus Preferences

In Subsection 3.3, we have documented that compliance probabilities vary by gender and vulnerability status. We now apply an Oaxaca (1973)-Blinder (1973) decomposition to the model in order to decompose the observed differences in the (log of the) probability of choosing action *j ∈* 2-4 relative to action *j* = 1 between genders and vulnerability statuses into the share explained by group differences in perceived risks/returns (expectations) and the share explained by group differences in utility parameters (preferences).

Table 11 shows the results of the decomposition. The higher average compliance probabilities – and corresponding lower average noncompliance probabilities – of female respondents relative their male counterparts are explained by both differences in expectations and preferences; whereas, the higher average noncompliance probabilities – and corresponding lower average compliance probabilities – of nonvulnerable respondents relative to their vulnerable counterparts seems to be completely driven by differences in preferences between the two groups.

**Table 11:** Decomposition of (Non-)Compliance Probabilities into Expectations VS. Preferences

|  | **Differences in (log of) Subjective Probabilities of A2-A4 vs A1** |  |
| --- | --- | --- |
|  | **between...** |  |
|  | Females<br>VS. Males | Not Vulnerables<br>VS. Vulnerables |
| Overall Difference | -0.713*** | 2.056*** |
| Share Expectations | 0.399*** | 0.164 |
| Share Preferences | 0.762*** | 0.913*** |
| Share Interaction | -0.164 | -0.078 |

Note: Oaxaca-Blinder decomposition results. \*\*\*: $p < 0.01$ ; \*\*: $p < 0.05$ ; \*: $p < 0.1$ . The difference in choice probabilities among respondents with and without prior COVID-19 experience is not statistically significant.

### 4.6 Do People Need “Stay Home” Compensation?

At the peak of the pandemic, worried about further spread of infections, the UK Government introduced a debated compensation scheme for the self-isolating on low income. According to this scheme, workers on low income in parts of England with a high incidence of Coronavirus cases (e.g., Blackburn, Darwen, Pendle, and Oldham) could claim money. The scheme started off with a trial amount of £130 for eligible individuals who tested positive to the Coronavirus and had to self-isolate for 10 days, plus £182 to other members of their household who had to self-isolate for 14 days as a consequence.^60^ After the introduction of the NHS Test and Trace Service (TTS) at the end of May 2020, the trial compensation was eventually transformed into a £500 Test and Trace Support Payment for people on low incomes who had to self-isolate due to Coronavirus (for England only).^61^

Given the heated debate generated by the existence and amount of this compensation, we use our model and estimates to shed some light on the issue. In particular, following Delavande (2008), we use an indifference condition based on the model to compute for each individual in our sample the amount of money, 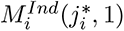, that makes her indifferent between her optimal compliance conduct, *j_i_^*^*, and the government’s “Stay Home” benchmark, *j* = 1,

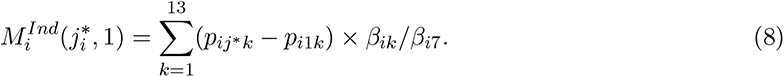

The empirical distribution of 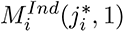 is shown in Figure 5. About 25% of the sample requires compensation to be indifferent between staying home and their optimal choice. The estimated mean compensation is £300-350 for four weeks.^62^ Consistent with the heterogeneity in preferences and expectations described earlier, vulnerable respondents are less likely to need compensation (22%) and require less-than-average compensation (£169-206).^63^ Respondents with prior COVID-19 experience are more likely to need compensation (26-27%) and require more-than-average compensation (£356-412). Male respondents are less likely to need compensation (20%), but require more-than-average compensation (£466-523). Of particular relevance to the public debate, we find that respondents on low income are less likely to need compensation (21%), but require more-than-average compensation (£556-577). This amount is in the ballpark of the amount granted by the Government in the trial phase (£130 over 10 days, plus an additional amount for family members), but substantially lower than the amount eventually granted at regime (£500 over 10 days).^64^

**Figure 5:**
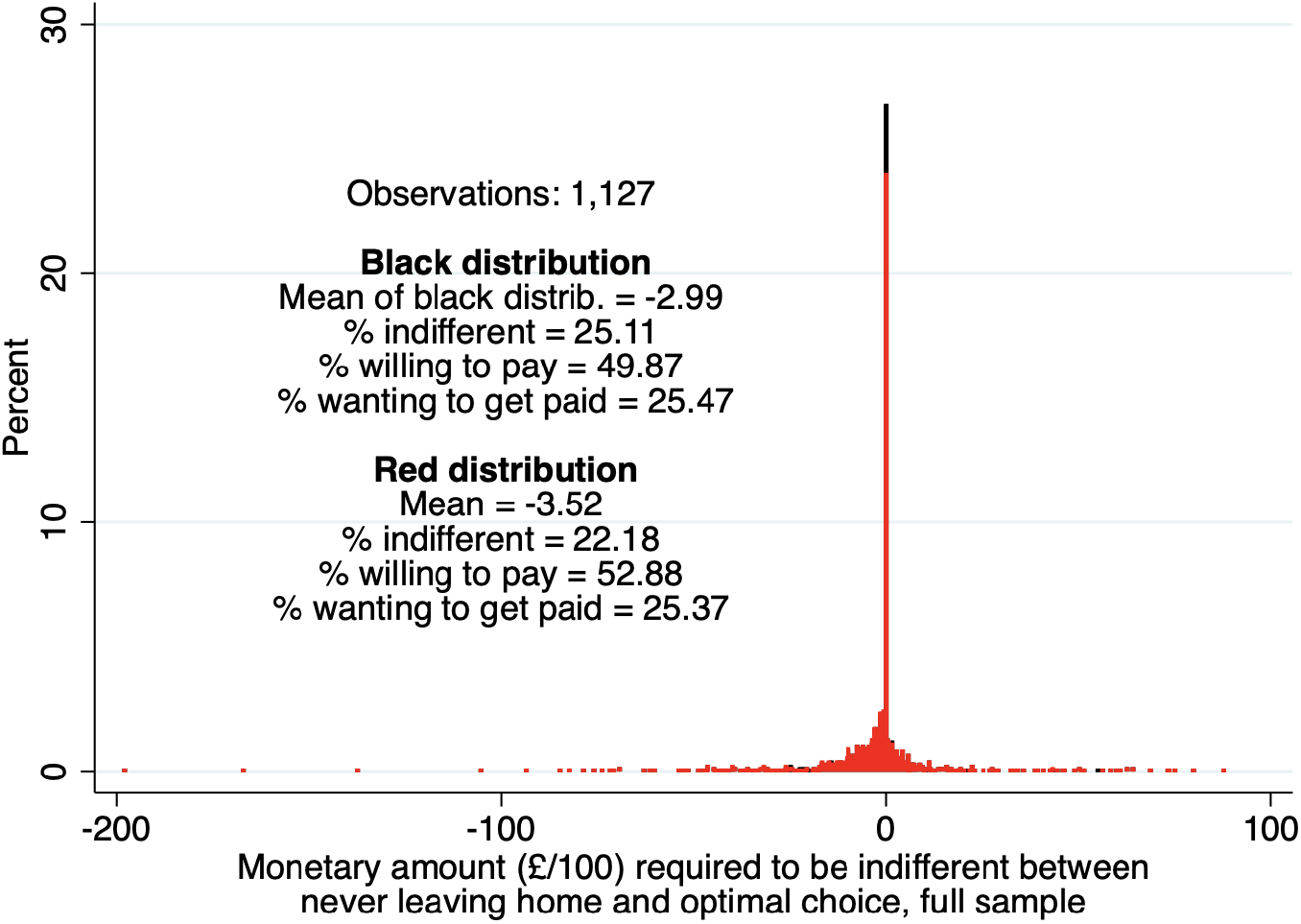
Distribution of the Monetary Amount Required to be Indifferent Between Never Leaving Home and Optimal Choice

## 5 The Role of Compliance Behavior of Others

Individual compliance decisions may not only depend on perceived personal costs and benefits – and how people resolve the tradeoffs among them – but also on the compliance behavior of others. In this section, we investigate how respondents’ compliance probabilities respond to others’ failure to comply in two ways. First, we study respondents’ perceived effect of the compliance behavior of others living in the same local authority (LA) on their own compliance plans. Second, we study the effect of the behavior of a high-level public figure, Dominic Cummings, on respondents’ own compliance.

### 5.1 The Effect of Others Living in the Same Municipality

In our baseline survey, we elicit respondent’s beliefs about the proportion of people living in their LA who will follow each of the four actions A1-A4 in the subsequent four weeks. We additionally elicit respondents’ subjective probabilities of following actions A1-A4 under alternative hypothetical scenarios about the compliance behavior of others living in their LA. We specify two scenarios. In a low-compliance scenario, the hypothesized distribution of others’ behavior is 10% A1, 15% A2, 25% A3, and 50% A4 – or 25% compliance versus 75% non-compliance. In a high-compliance scenario, the hypothesized distribution is 50% A1, 25% A2, 15% A3, and 10% A4 – or 75% compliance versus 25% non-compliance.

We use the latter measures to construct within-respondent differences in action-specific probabilities between low- and high-compliance scenarios, that is, P(A*_j_|*Low compliance) *−* P(A*_j_|*High compliance) for *j* = 1, 2, 3, 4.^65^ We additionally aggregate the latter across compliance actions (A1 + A2) and non-compliance actions (A3 + A4). Table 12 reports mean and standard deviation of the empirical distributions of these measures in the overall sample and in selected sub-samples (by vulnerability status, gender, COVID-19 experience, COVID-19 literacy, and risk tolerance). On average, moving from a high to a low scenario of others’ compliance implies a decrease in the respondent’s own probability of strict compliance (A2) and an increase in both the probability of staying at home (A1) and the probabilities of non-compliance (A3-A4). Thus, the average response is non-monotonic across actions, suggesting that when surrounding others comply less and trust breaks down, some individuals expect to do the same (increased likelihood of A3-A4), while others expect to engage in protective behavior (increased likelihood of A1).

**Table 12:** Perceived Effects of Others’ Compliance (Low vs. High) on Own Compliance

| All | mean | sd | N | Low risk tolerance | mean | sd | N | COVID-19<br>Lit. Index > 0 | mean | sd | N |
| --- | --- | --- | --- | --- | --- | --- | --- | --- | --- | --- | --- |
| A1 - Never leave home | 0.95 | 15.89 | 1,132 | A1 - Never leave home | 2.21 | 16.23 | 609 | A1 - Never leave home | 0.90 | 16.77 | 558 |
| A2 - Strict compliance | -2.91 | 16.35 | 1,132 | A2 - Strict compliance | -4.07 | 16.46 | 609 | A2 - Strict compliance | -2.84 | 16.60 | 558 |
| A3 - General compliance | 0.61 | 13.42 | 1,132 | A3 - General compliance | 0.85 | 13.29 | 609 | A3 - General compliance | 0.78 | 13.42 | 558 |
| A4 - Non-compliance | 1.35 | 9.45 | 1,132 | A4 - Non-compliance | 1.01 | 7.75 | 609 | A4 - Non-compliance | 1.15 | 8.72 | 558 |
| A1 + A2 - Compliance | -1.96 | 15.78 | 1,132 | A1 + A2 - Compliance | -1.86 | 14.84 | 609 | A1 + A2 - Compliance | -1.94 | 15.70 | 558 |
| A3 + A3 - Non-compliance | 1.96 | 15.78 | 1,132 | A3 + A3 - Non-compliance | 1.86 | 14.84 | 609 | A3 + A3 - Non-compliance | 1.94 | 15.70 | 558 |
|  |  |  |  | High risk tolerance | mean | sd | N | COVID-19<br>Lit. Index = 0 | mean | sd | N |
|  |  |  |  | A1 - Never leave home | -0.52 | 15.38 | 523 | A1 - Never leave home | 1.00 | 15.01 | 574 |
|  |  |  |  | A2 - Strict compliance | -1.55 | 16.13 | 523 | A2 - Strict compliance | -2.98 | 16.13 | 574 |
|  |  |  |  | A3 - General compliance | 0.33 | 13.57 | 523 | A3 - General compliance | 0.44 | 13.42 | 574 |
|  |  |  |  | A4 - Non-compliance | 1.74 | 11.11 | 523 | A4 - Non-compliance | 1.54 | 10.12 | 574 |
|  |  |  |  | A1 + A2 - Compliance | -2.07 | 16.81 | 523 | A1 + A2 - Compliance | -1.98 | 15.86 | 574 |
|  |  |  |  | A3 + A3 - Non-compliance | 2.07 | 16.81 | 523 | A3 + A3 - Non-compliance | 1.98 | 15.86 | 574 |
| Vulnerables | mean | sd | N | Males | mean | sd | N | COVID-19<br>Exp. Index > 0 | mean | sd | N |
| A1 - Never leave home | 0.31 | 13.13 | 115 | A1 - Never leave home | 1.04 | 15.13 | 559 | A1 - Never leave home | 1.39 | 16.67 | 783 |
| A2 - Strict compliance | 0.39 | 8.86 | 115 | A2 - Strict compliance | -2.89 | 15.23 | 559 | A2 - Strict compliance | -3.08 | 16.98 | 783 |
| A3 - General compliance | -0.19 | 8.42 | 115 | A3 - General compliance | 0.48 | 12.93 | 559 | A3 - General compliance | .238 | 13.61 | 783 |
| A4 - Non-compliance | -0.51 | 7.04 | 115 | A4 - Non-compliance | 1.38 | 9.27 | 559 | A4 - Non-compliance | 1.45 | 9.87 | 783 |
| A1 + A2 - Compliance | 0.70 | 11.55 | 115 | A1 + A2 - Compliance | -1.86 | 15.08 | 559 | A1 + A2 - Compliance | -1.69 | 15.90 | 783 |
| A3 + A3 - Non-compliance | -0.70 | 11.55 | 115 | A3 + A3 - Non-compliance | 1.86 | 15.08 | 559 | A3 + A3 - Non-compliance | 1.69 | 15.90 | 783 |
| Non-Vulnerables | mean | sd | N | Females | mean | sd | N | COVID-19<br>Exp. Index = 0 | mean | sd | N |
| A1 - Never leave home | 1.02 | 16.18 | 1,017 | A1 - Never leave home | 0.72 | 16.47 | 568 | A1 - Never leave home | -0.03 | 13.96 | 349 |
| A2 - Strict compliance | -3.28 | 16.96 | 1,017 | A2 - Strict compliance | -2.86 | 17.37 | 568 | A2 - Strict compliance | -2.52 | 14.86 | 349 |
| A3 - General compliance | 0.70 | 13.87 | 1,017 | A3 - General compliance | 0.88 | 13.75 | 568 | A3 - General compliance | 1.44 | 12.95 | 349 |
| A4 - Non-compliance | 1.56 | 9.67 | 1,017 | A4 - Non-compliance | 1.27 | 9.57 | 568 | A4 - Non-compliance | 1.11 | 8.45 | 349 |
| A1 + A2 - Compliance | -2.26 | 16.16 | 1,017 | A1 + A2 - Compliance | -2.14 | 16.36 | 568 | A1 + A2 - Compliance | -2.55 | 15.49 | 349 |
| A3 + A3 - Non-compliance | 2.26 | 16.16 | 1,017 | A3 + A3 - Non-compliance | 2.14 | 16.36 | 568 | A3 + A3 - Non-compliance | 2.55 | 15.49 | 349 |

Note: The table reports statistics (mean and standard deviation) of the distributions of within-respondent differences in compliance probabilities between two hypothetical scenarios – a low-compliance one and a high-compliance one – describing the compliance behavior of people living in the same local authority as the respondent. In the low-compliance scenario, the distribution of behavior in the respondent's local authority is 10% A1, 15% A2, 25% A3, and 50% A4 (or 25% compliance vs. 75% non-compliance). In the high-compliance scenario, the distribution is 50% A1, 25% A2, 15% A3, and 10% A4 (or 75% compliance vs. 25% non-compliance).

To investigate this further, we look at patterns across sub-groups. We find that vulnerable respondents react by increasing the probability of both compliance actions (A1-A2) and decreasing the probability of non-compliance actions (A3-A4). Conversely, respondents with no prior COVID-19 experience and those with high risk tolerance react by decreasing the probability of both compliance actions (A1-A2) and increasing the probability of both non-compliance actions (A3-A4).

Individuals’ expected response may also depend on their prior belief about the compliance behavior of those living around them and on the extent to which the hypothesized scenario differs from it. To study this possibility we construct two ‘shock’ measures. The first is defined as “25% - the respondent’s perceived percentage of others’ complying (A1+A2)”, where 25% is the hypothesized proportion of compliers (A1+A2) in the low-compliance scenario. The second is defined as “75% - the respondent’s perceived percentage of others’ complying”, where 75% is the hypothesized proportion of compliers in the high-compliance scenario. In the left panel of Table 13, we regress respondents’ own probability of complying (A1+A2) under the scenario of others’ low compliance on the first shock measure. We do so in the overall sample (column 1), and also separately for respondents whose hypothetical shock is negative (column 2) and for respondents whose hypothetical shock is positive (column 3). The former group is more common than the latter, as on average respondents believe that about 60% of others around them comply to the lockdown rules (A1+A2); see Figure A9. Indeed, for the low-compliance case, the average hypothetical shock in the sample is about -34%. In the right panel of Table 13, we estimate similar regressions using the compliance probability and shock measures under the scenario of others’ high compliance. Obviously, in this case the most common hypothetical shock is positive, with the average hypothetical shock in the sample being about 16%.

**Table 13:** Response of Own PC of Complying (A1+A2) in Low/High Scenario to How Scenario Differs from Own Belief about Others’ Compliance (“Shock”)

|  | **PC of Complying (A1+A2)**<br>**if Low Others' Compliance** |  |  | **PC of Complying (A1+A2)**<br>**if High Others' Compliance** |  |  |
| --- | --- | --- | --- | --- | --- | --- |
| | All<br>(1) | Shock < 0<br>(2) | Shock ≥ 0<br>(3) | All<br>(4) | Shock < 0<br>(5) | Shock ≥ 0<br>(6) |
| Shock | -0.489***<br>(0.038) | -0.467***<br>(0.042) | 0.356<br>(0.545) | -0.515***<br>(0.037) | -0.075<br>(0.150) | -0.590***<br>(0.058) |
| Constant | 57.650***<br>(1.738) | 58.769***<br>(1.980) | 47.971***<br>(5.801) | 84.476***<br>(0.684) | 88.173***<br>(1.896) | 86.817***<br>(1.362) |
| N | 1,132 | 1,036 | 96 | 1,132 | 299 | 833 |

Note: The shock is defined as "25% - R's perceived percentage of others' complying (A1+A2)" for the low-compliance scenario and as "75% - R's perceived percentage of others' complying (A1+A2)" for the high-compliance scenario.

We find that, on average, individuals respond by lowering their compliance probability in both scenarios; see columns 1 and 4. In the low-compliance scenario, the result is driven by respondents whose hypothetical shock is negative (column 2), while in the high-compliance scenario it is driven by respondents whose hypothetical shock is positive (column 6). Respondents in column 2 have prior beliefs about others’ compliance that are more optimistic than the behavior of others hypothesized in the low-compliance scenario they are given. These respondents plan to react to the lower-than-expected compliance of others by lowering their own compliance. On the other hand, respondents in column 6 have prior beliefs about others’ compliance that are more pessimistic than the behavior of others hypothesized in the high-compliance scenario they are given. These respondents, too, plan to react to the higher-than-expected compliance of others by lowering their own compliance.

### 5.2 The “Cummings Effect”

In this last sub-section, we study the effect of compliance behavior of a high-level public figure: the former Prime Minister Boris Johnson’s chief aide, Dominic Cummings. As mentioned in 3.1.2, we used the launch of the NHS Test and Trace Service (TTS) on May 28th 2020 as pretense to field a short follow-up survey and implement a randomized sensitization intervention based on the “Dominic Cummings scandal”. A random half of the sample was shown the “Cummings screen” showcasing the timeline of the scandal (see Figure A3) at the beginning of the survey, and the other half of the sample was shown the screen at the end of the survey. We estimate the impact of this treatment on respondents’ compliance probabilities; given the political salience of our treatment, we do so separately for respondents supporting the Labour party and respondents supporting the Conservative party (measured at baseline).

Treatment effects estimates for the follow-up sample and the panel sample are shown in Tables 14 and 15, respectively. We find that reported compliance probabilities are sensitive to negative prompts. In Table 14, respondents randomized to the Cummings treatment (i.e., those who were shown the negative prompt before the expectation questions) report a lower probability (-7.6 p.p.) of A1 (never leave home) in the next 4 weeks, and a corresponding higher probability (+7.4 p.p.) of A3 (general compliance), but only if they support the Labour party. In Table 15, we exploit the panel structure of the data and evaluate whether the treatment changed the persistence of respondents’ compliance probabilities between baseline and follow-up. We find that those randomized to the Cummings treatment show a higher persistence of A3 (general compliance) in the next 4 weeks, independently of their political inclination.

**Table 14:**
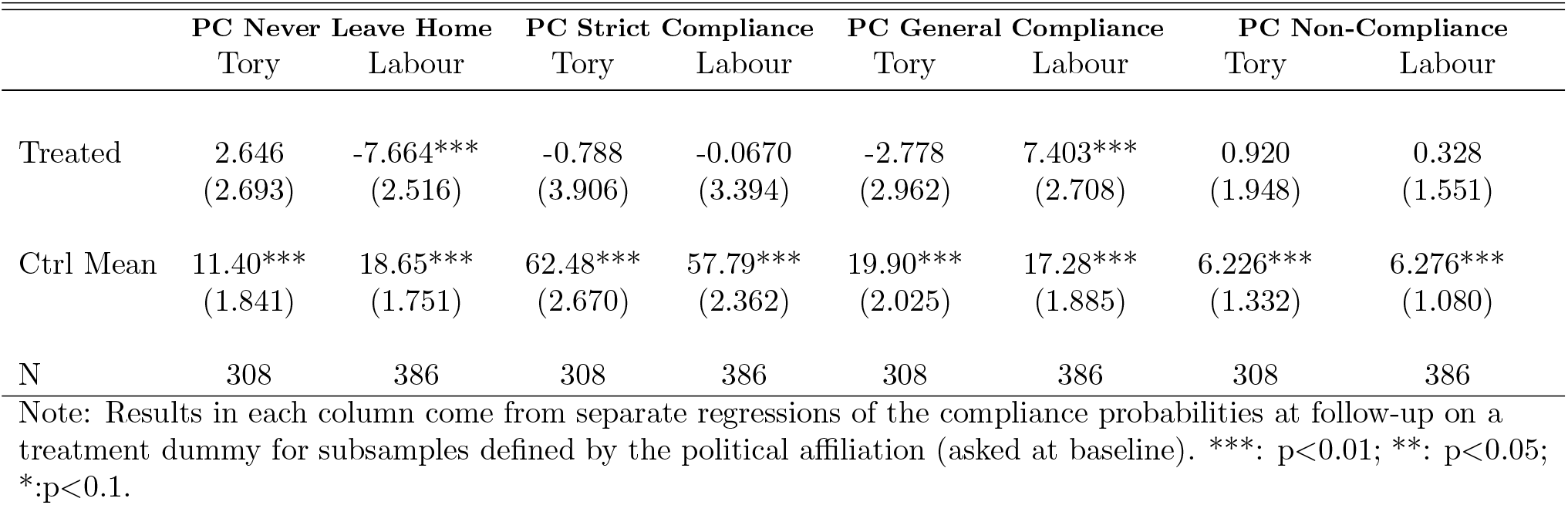
Treatment Effects, Cummings Sensitization Treatment – Follow-up Sample

|  | PC Never Leave Home |  | PC Strict Compliance |  | PC General Compliance |  | PC Non-Compliance |  |
| --- | --- | --- | --- | --- | --- | --- | --- | --- |
|  | Tory | Labour | Tory | Labour | Tory | Labour | Tory | Labour |
| Treated | 2.646<br>(2.693) | -7.664***<br>(2.516) | -0.788<br>(3.906) | -0.0670<br>(3.394) | -2.778<br>(2.962) | 7.403***<br>(2.708) | 0.920<br>(1.948) | 0.328<br>(1.551) |
| Ctrl Mean | 11.40***<br>(1.841) | 18.65***<br>(1.751) | 62.48***<br>(2.670) | 57.79***<br>(2.362) | 19.90***<br>(2.025) | 17.28***<br>(1.885) | 6.226***<br>(1.332) | 6.276***<br>(1.080) |
| N | 308 | 386 | 308 | 386 | 308 | 386 | 308 | 386 |

Note: Results in each column come from separate regressions of the compliance probabilities at follow-up on a treatment dummy for subsamples defined by the political affiliation (asked at baseline). \*\*\*: $p < 0.01$ ; \*\*: $p < 0.05$ ; \*: $p < 0.1$ .

**Table 15:** Effects of Cummings Sensitization Treatment – Panel Sample

| | $P(A1_{t1})$ | $P(A2_{t1})$ | $P(A3_{t1})$ | $P(A4_{t1})$ |
| --- | --- | --- | --- | --- |
| Treated | 0.626<br>(1.736) | -2.954<br>(4.044) | -1.164<br>(2.037) | -0.325<br>(1.127) |
| $P(A1_{t0})$ | 0.475***<br>(0.0318) | | | |
| $Treated \times P(A1_{t0})$ | -0.0675<br>(0.0465) | | | |
| $P(A2_{t0})$ | | 0.439***<br>(0.0444) | | |
| $Treated \times P(A2_{t0})$ | | 0.0188<br>(0.0640) | | |
| $P(A3_{t0})$ | | | 0.357***<br>(0.0462) | |
| $Treated \times P(A3_{t0})$ | | | 0.189***<br>(0.0670) | |
| $P(A4_{t0})$ | | | | 0.191***<br>(0.0642) |
| $Treated \times P(A4_{t0})$ | | | | 0.165<br>(0.101) |

Note: Results in each column come from separate regressions of the compliance probabilities at follow-up ( $t1$ ) on a treatment, dummy, the compliance probabilities at baseline ( $t0$ ) and their interactions. $t1$ = follow-up, data collected 28 May 2020 $t0$ = baseline, data collected 3-10 May 2020. N=905. \*\*\*: $p < 0.01$ ; \*\*: $p < 0.05$ ; \*: $p < 0.1$ .

The vast majority of respondents (95.72%) reported being aware about the Cummings episode, and a significant majority (81.94%) thought that Cummings had broken the lockdown rules.^66^ Hence, we view our sensitization treatment as increasing the salience – rather than the awareness – of the Cummings episode. In this light, we interpret our findings as indicative of the role that trust in the government plays in affecting compliance behavior. We enrich the existing evidence, which has either documented associations between trust and compliance (e.g., Wright, Steptoe and Fancourt (2021), Fancourt, Steptoe and Wright (2020), and Bird et al. (2023)),^67^ or willingness to comply (e.g., Pagliaro et al. (2021) and Burton et al. (2022)), or emphasized the role of trust as moderator of compliance (Bargain and Aminjonov, 2020).^68^ Ours is the first evidence based on a randomized treatment.

## 6 Conclusion

Understanding why some individuals engage more in healthy behaviors than others is a fundamental question in the health sciences, still being actively explored across several disciplines. Understanding drivers of healthy behaviors, and the trade-offs individuals face in terms of costs and benefits of alternative courses of actions that are *ex ante* uncertain, is crucial for designing effective and sustainable behavioral change interventions. In this paper, we have studied the role of individual beliefs, individual preferences, and others’ behaviors in explaining differences in individuals’ health behaviors within the context of the first COVID-19 lockdown in the UK – a time of significant uncertainty, when the consequences of compliance (or lack thereof) to the social distancing and self-isolation rules could make the difference between health and (a potentially fatal) disease.

We have collected rich survey data on respondents’ compliance plans, their perceived risks and returns of alternative compliance behaviors, and individual characteristics and attributes at the peak of the first UK lockdown (3-10 May 2020). In the first part of the paper, we have used these data to estimate a simple model of compliance behavior with uncertain costs and benefits. Our estimates enabled us to quantify the utility trade-offs underlying compliance, to decompose group differences in compliance plans, and to compute the monetary compensation required for people to be isolated.

Overall, we have found large disutilities from dying of COVID-19 and being caught transgressing, and large utilities from preserving a good mental health. We have also documented substantial heterogeneity in preferences and beliefs. For instance, vulnerable individuals have higher disutilities from contracting the Coronavirus and higher perceived risks associated with leaving home, while males have higher disutilities from becoming physically unfit and in general lower perceived risks from non-compliance. Differences in compliance probabilities across genders are explained by both differences in expectations and preferences, whereas differences in preferences seem the main source of variation explaining differences in compliance probabilities between vulnerable and non-vulnerable individuals. This suggests that interventions providing information on the actual risks of non-compliance could potentially improve adherence to the compliance rules while also supporting individuals in bearing the costs of compliance (e.g., see Burton et al. (2022) and Ryan et al. (2021)).

Using an indifference condition based on the model, we have computed the compensation required for people to comply and have found that approximately a quarter of the sample requires compensation to be indifferent between never leaving home (the conduct recommended by the government) and their optimal choice, with substantial heterogeneity in the amount required. Notably, our model-based compensation for low-income individuals aligns well with the amount provided by the government for self-isolating people on low incomes, providing a sound basis for the advocates of financial support to increase adherence to the public health guidelines (e.g., see Smith et al. (2021) and Ryan et al. (2021)).

In the second part of the paper, we have studied the relationship between own compliance and the compliance of others – those residing in the same local authority as the respondent as well as a high-level public figure, Dominic Cummings – using hypothetical scenarios and a randomized sensitization intervention.

We have found that, on average, moving from a high to a low scenario of others’ compliance implies a decrease in the respondent’s own probability of strict compliance (A2) and an increase in both the probability of staying at home (A1) and the probabilities of non-compliance (A3-A4). Thus, the average response is non-monotonic across actions, possibly suggesting that when surrounding others comply less, some individuals expect to do the same (increased likelihood of A3-A4), while others expect to engage in protective behavior (increased likelihood of A1). Indeed, this average responses mask different patterns across sub-groups, such as the vulnerables, who engage in more protective behaviors when exposed to lower compliance of others, or those with high risk tolerance, who engage in more relaxed behavior.

Lastly, we have found that a group of respondents, those supporting the Labour party, react to the Cummings treatment’s negative prompt by lowering their subjective probability of never leaving home and increasing that of discretionary compliance – providing a behavioral basis to all the work pointing to the role of political affiliation and trust in the government in compliance behavior. We have also found that those randomized to the Cummings treatment show a higher persistence of A3 (general compliance) in the next 4 weeks, independently of their political inclination.

These findings emphasize the need for public health policies to account for heterogenous beliefs, preferences, and responses to others in citizens’ health behaviors. While the COVID-19 pandemic is no longer a public health emergency, our analysis provides valuable insights for management of future pandemics as well as a portable framework applicable to other health behaviors under uncertainty, where it might be useful or of interest to disentangle individuals’ preferences and beliefs over the consequences of alternative behaviors, to compute possible subsidies aimed at improving the take-up of positive behaviors, and to design behavioral-change interventions by targeting (incorrect) beliefs.

## Data Availability

All data produced in the present study are available upon reasonable request to the authors, after publication.

## A Supplementary Online Appendix Not for Publication

**Figure A1:**
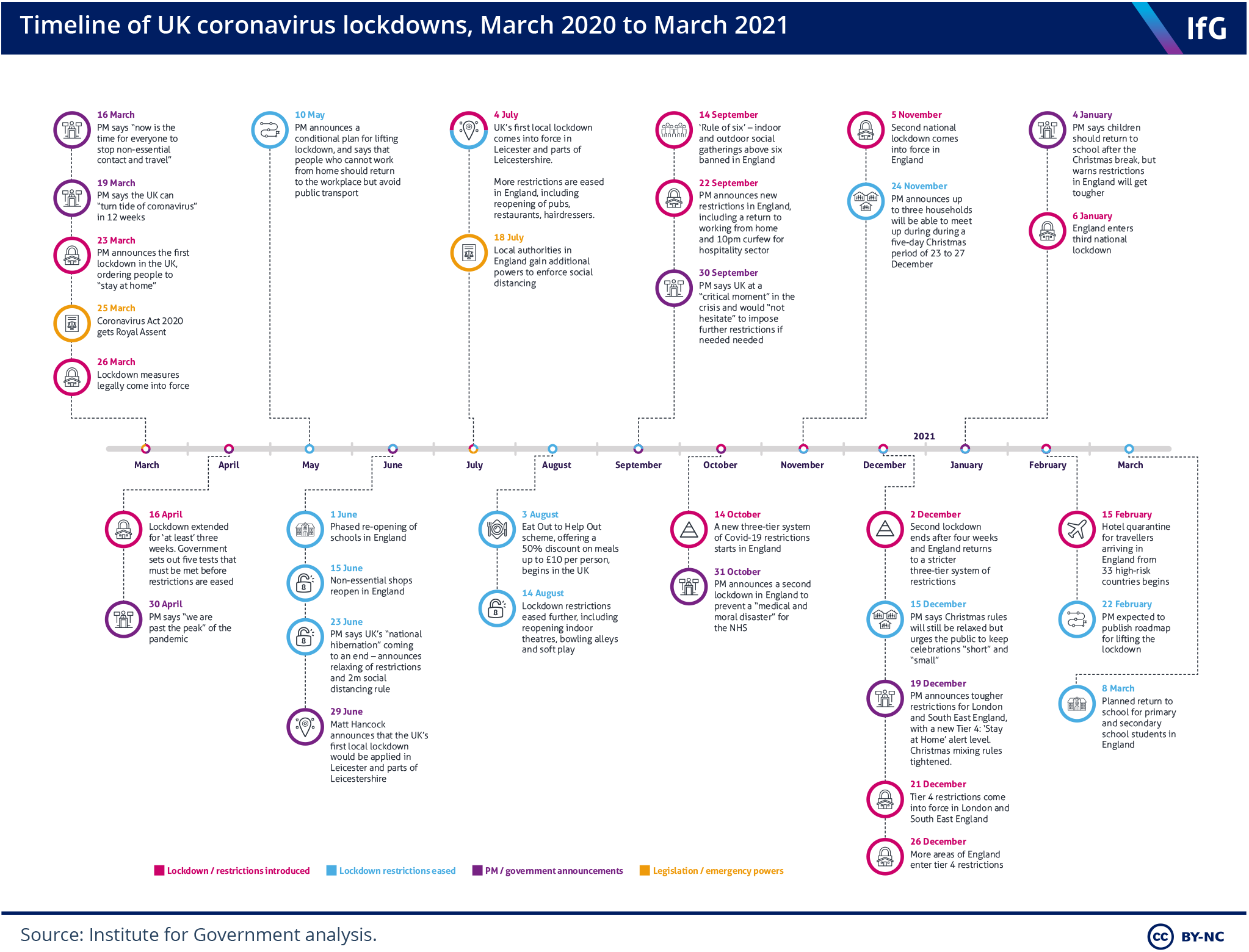
Timeline of UK COVID-19 Lockdowns

**Figure A2:**
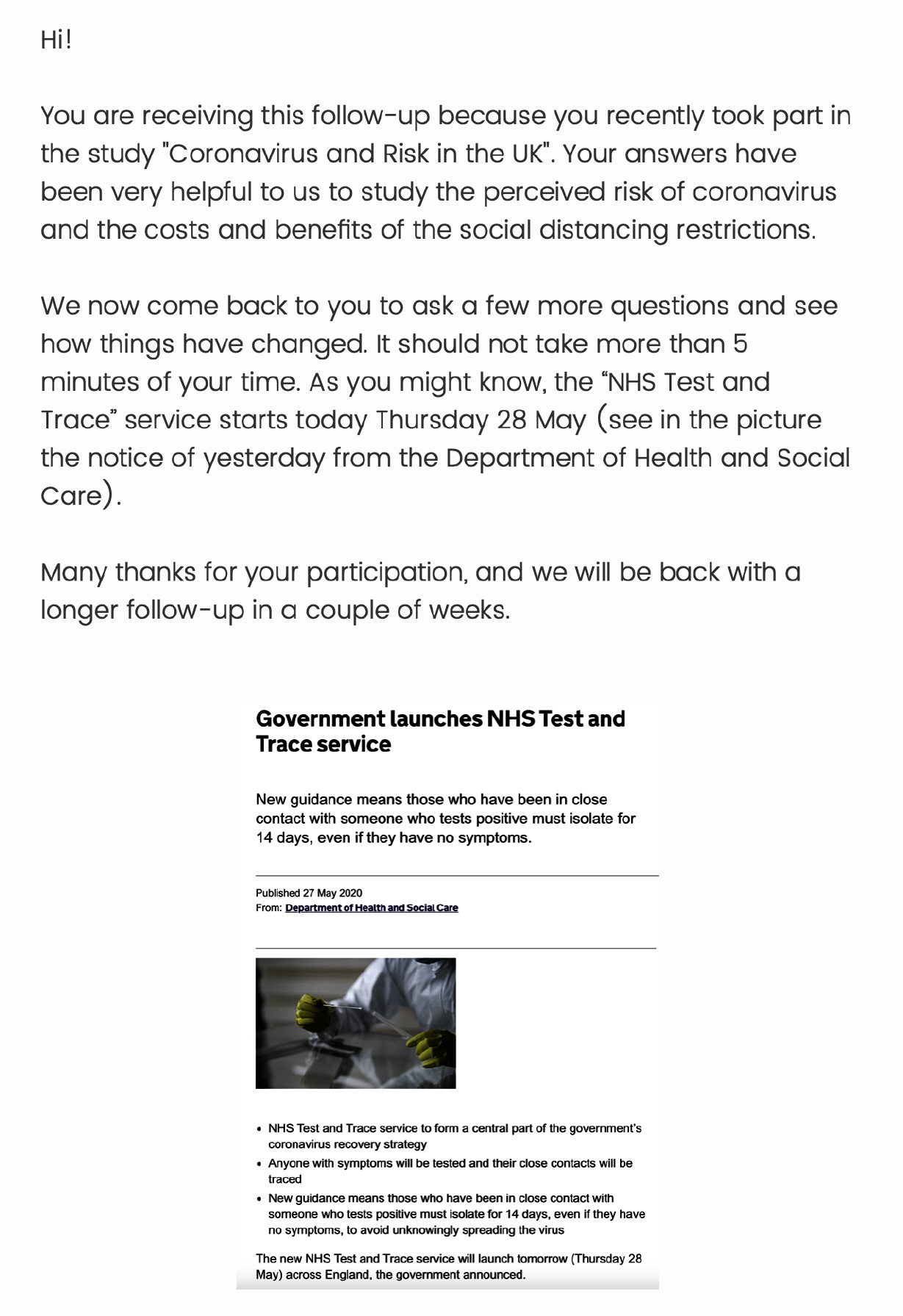
Introductory Screen to the Cummings Follow-Up

**Figure A3:**
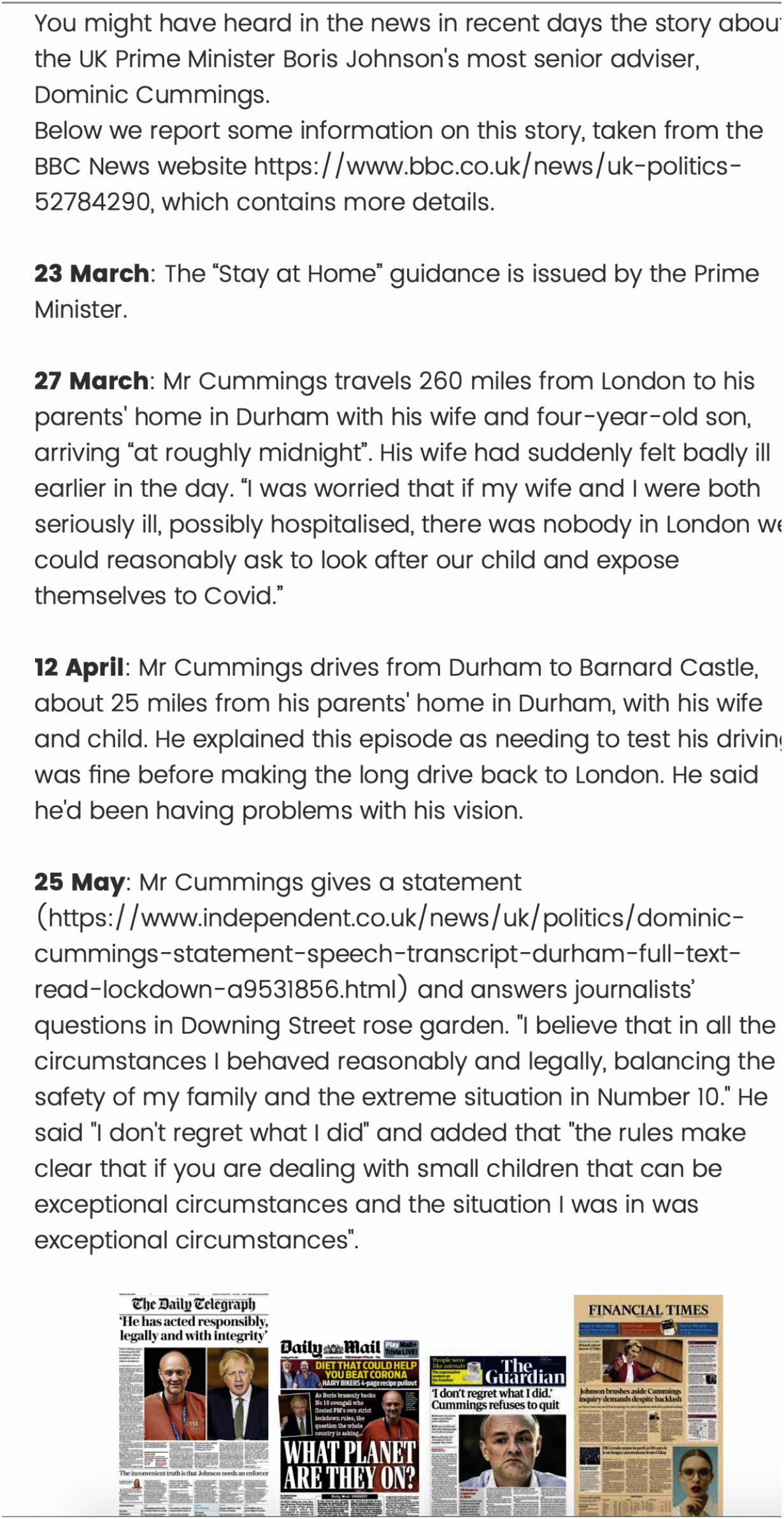
Timeline of the Cummings Affair

**Figure A4:**
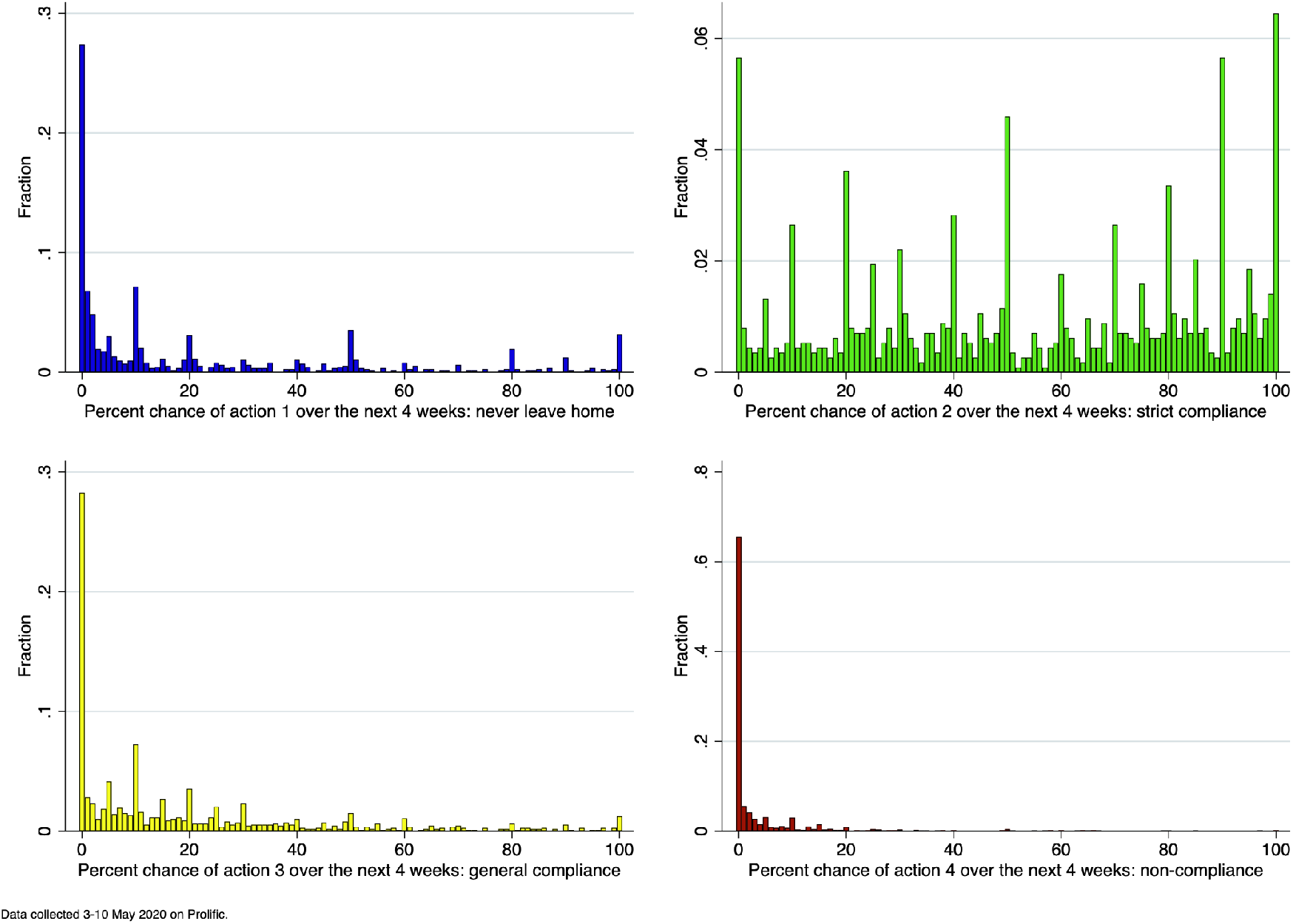
Percent Chances of Actions 1, 2, 3, and 4

**Figure A5:**
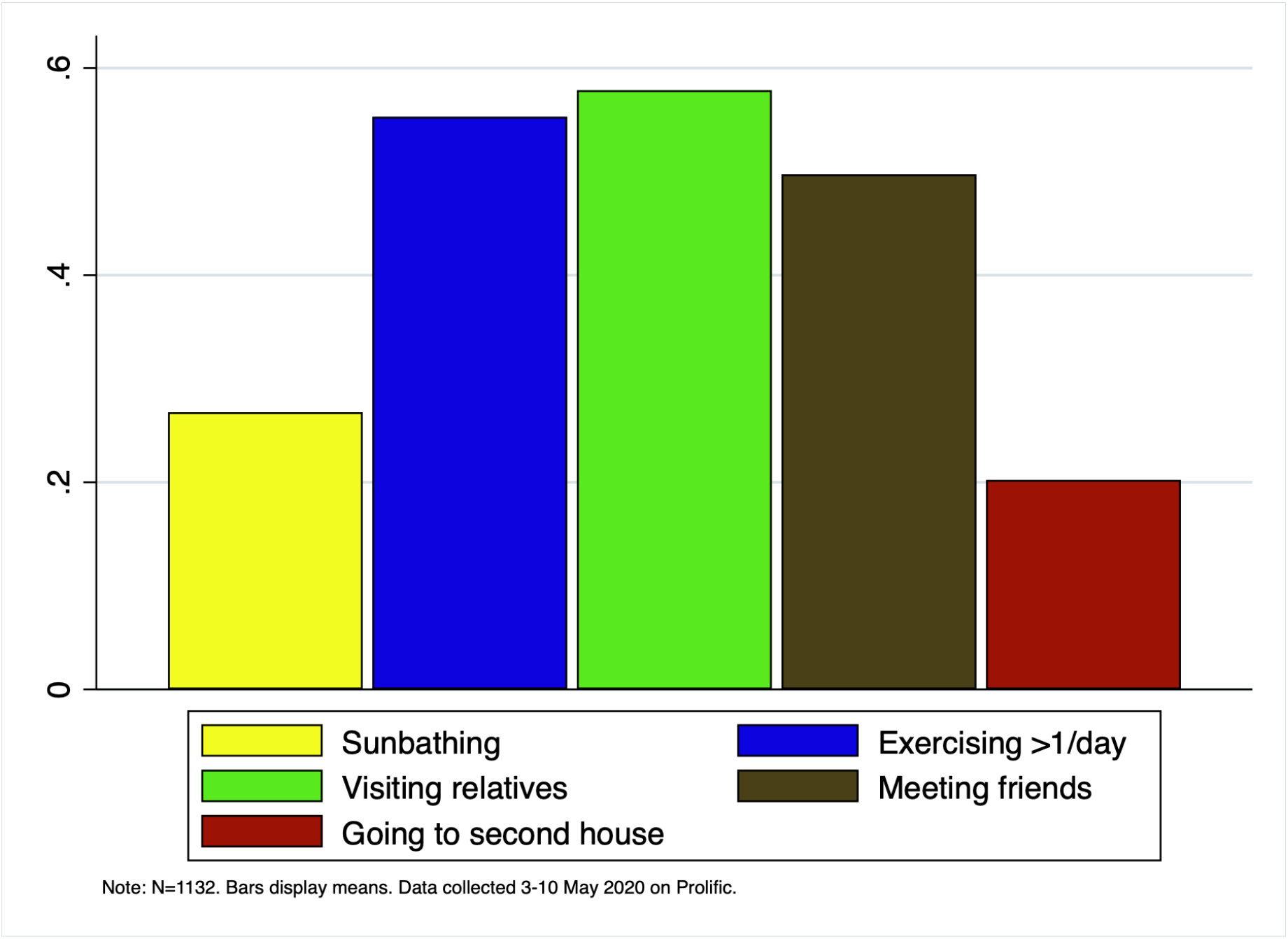
Most Common Self-Reported Non-Compliance Behaviors

**Figure A6:**
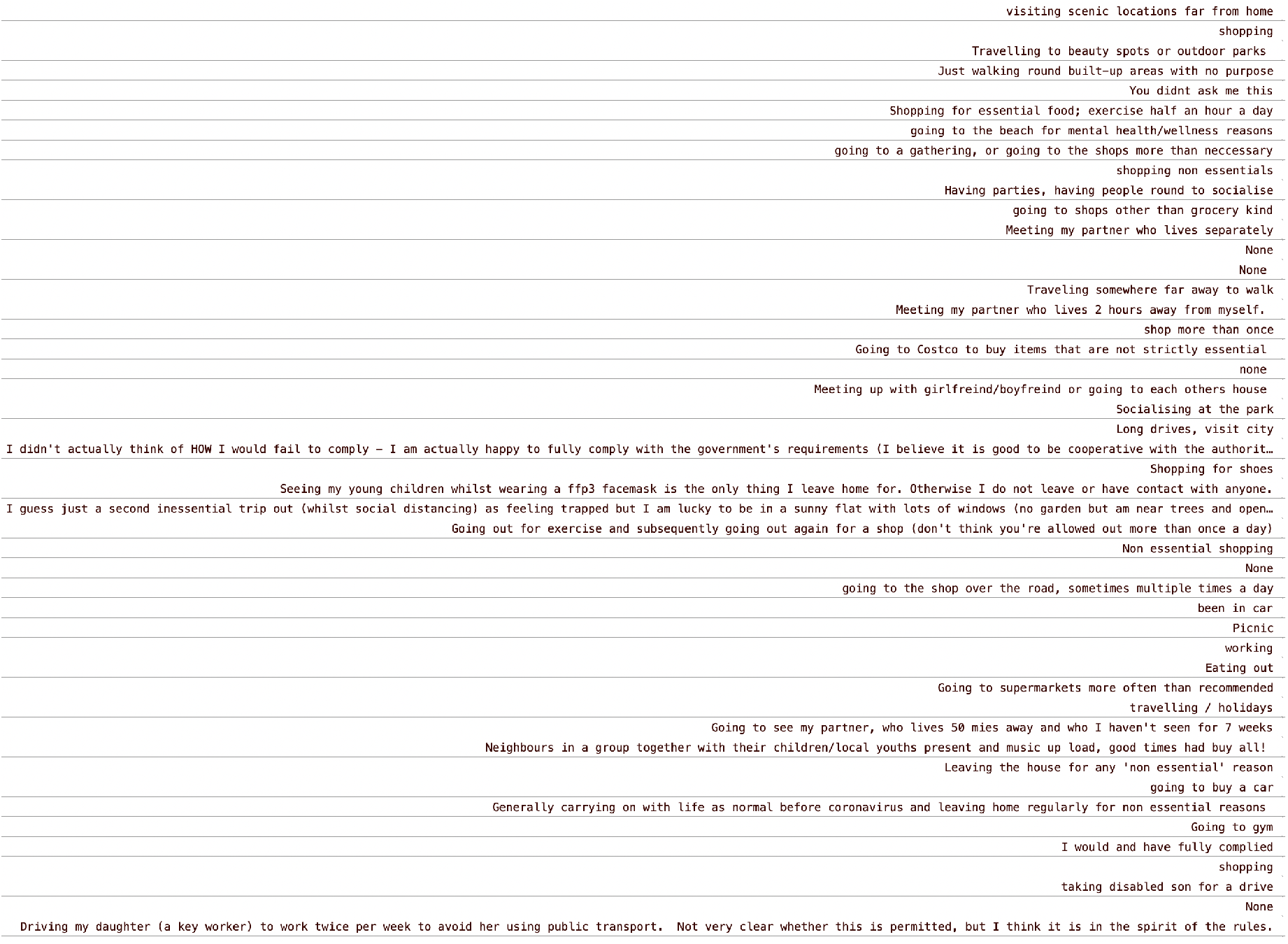
Quotes of Self-Reported Non-Compliance Behaviors

**Figure A7:**
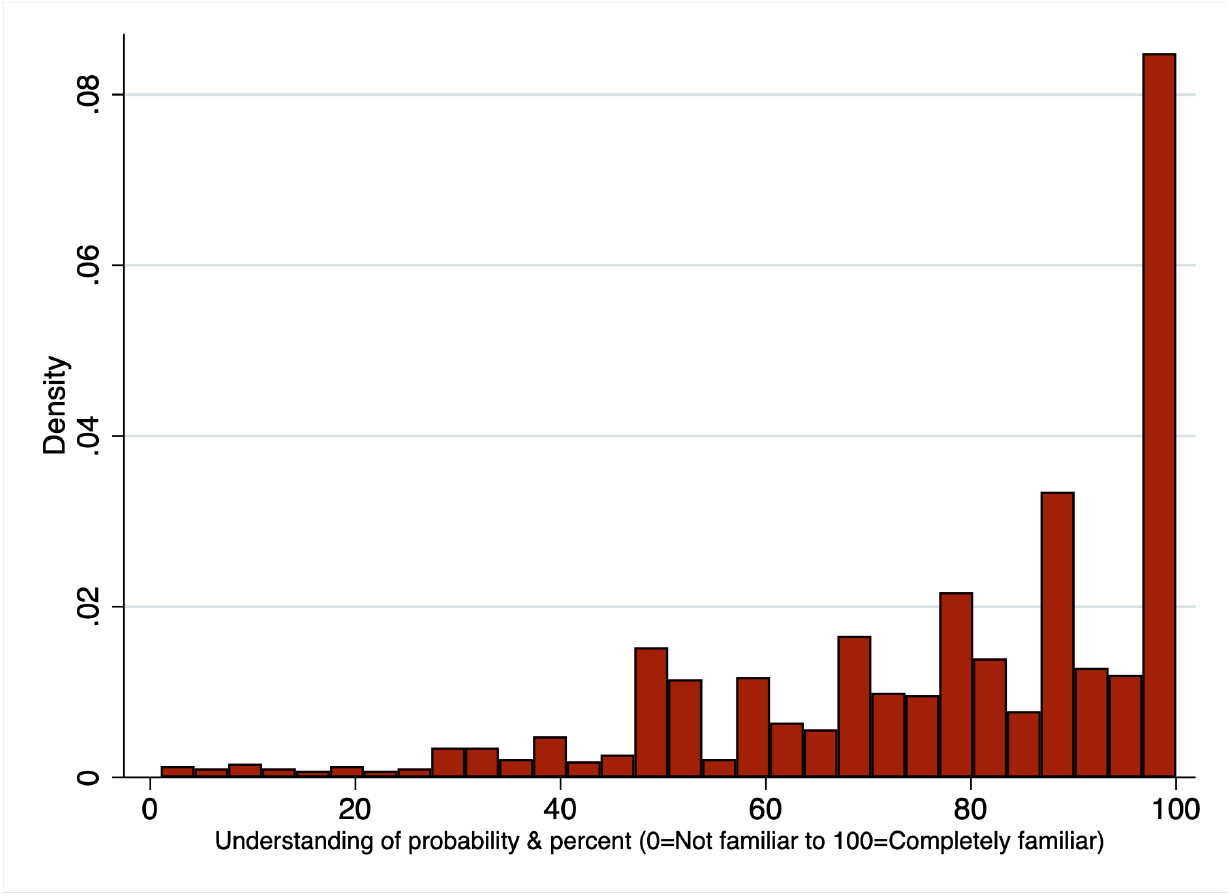
Histogram of Self-Reported Understanding of Probability

**Figure A8:**
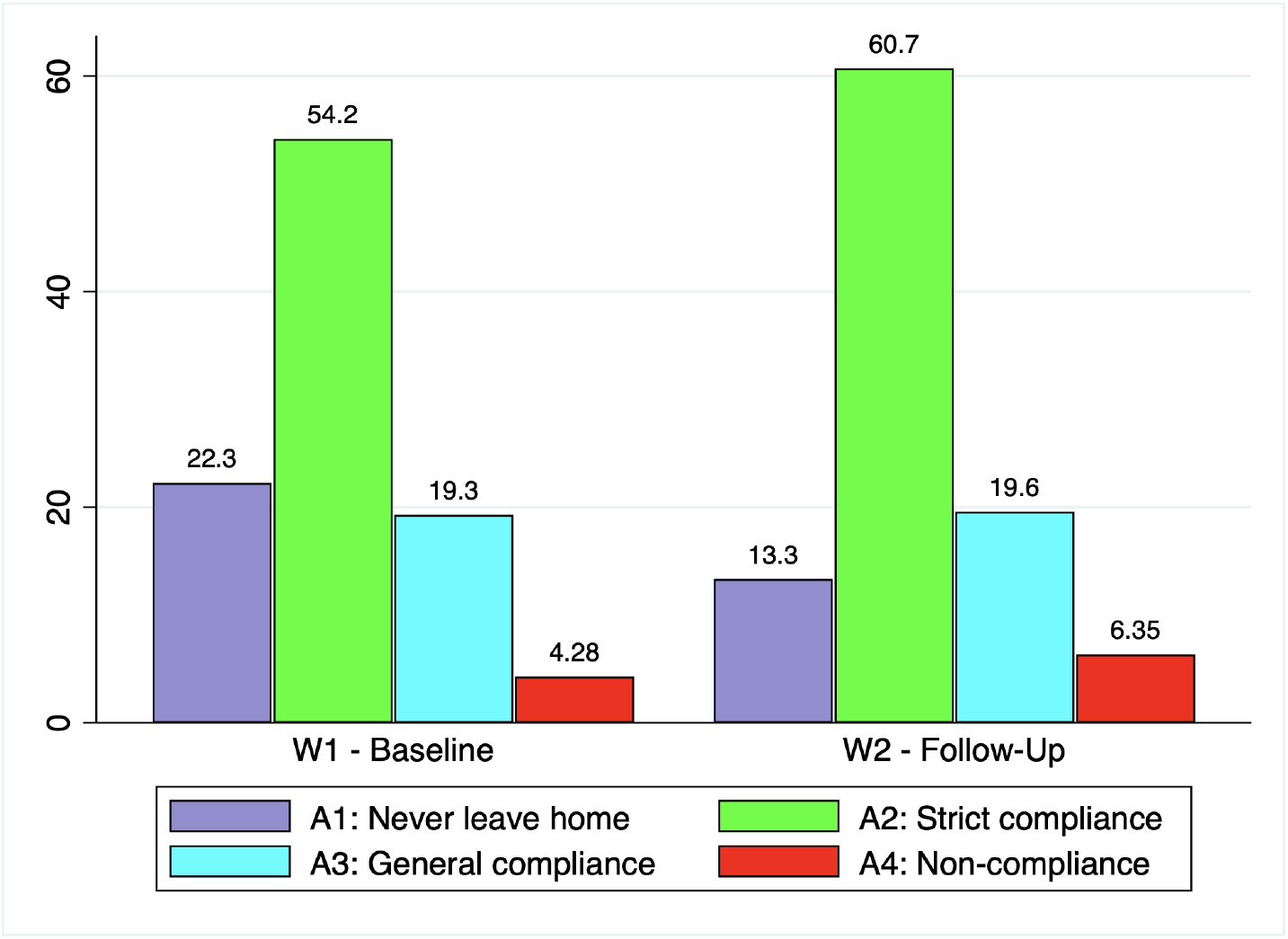
Heterogeneity in Compliance Probabilities by Wave

**Figure A9:**
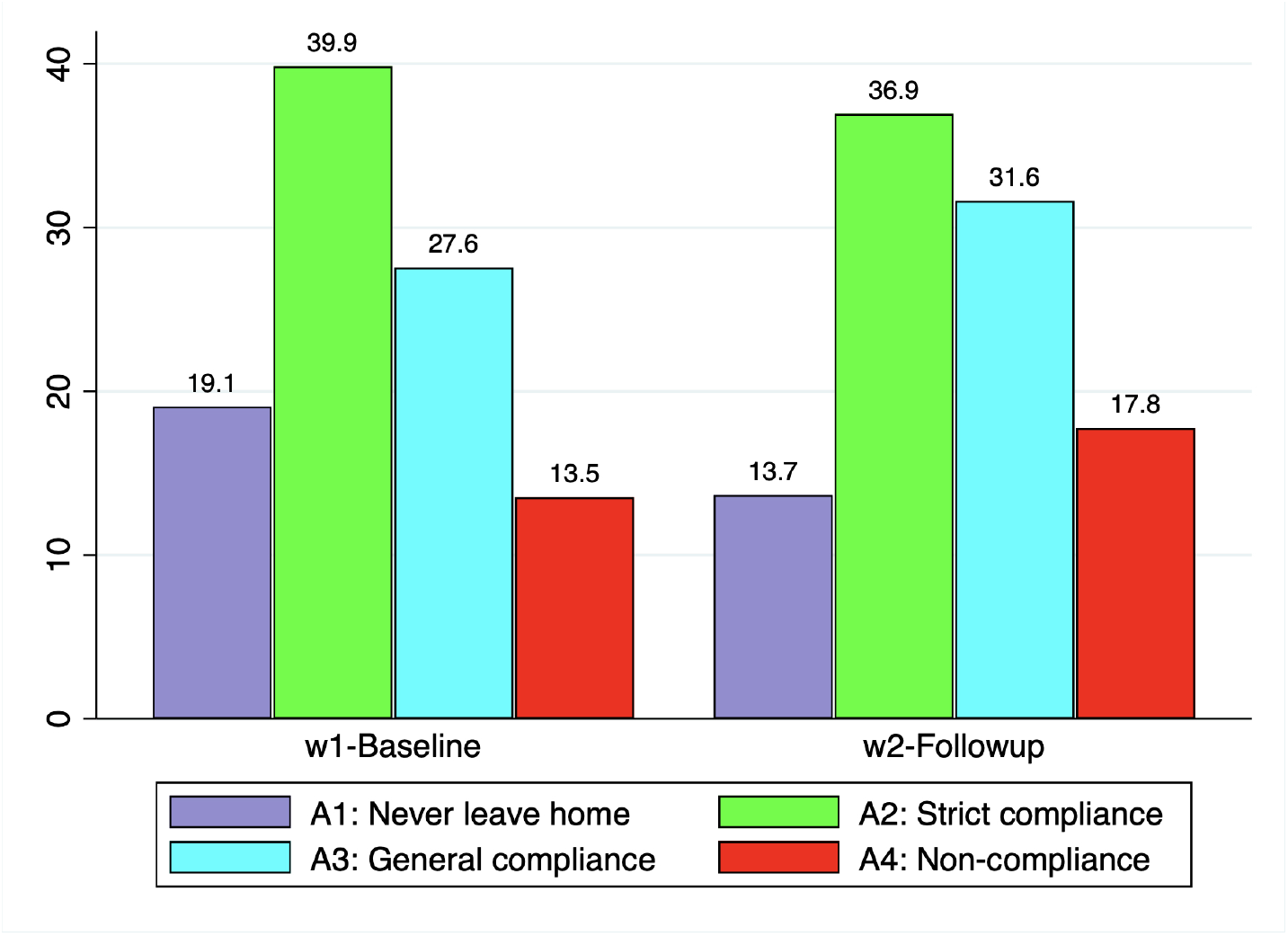
Heterogeneity in Perceptions of Others’ Compliance Behavior by Wave

**Figure A10:**
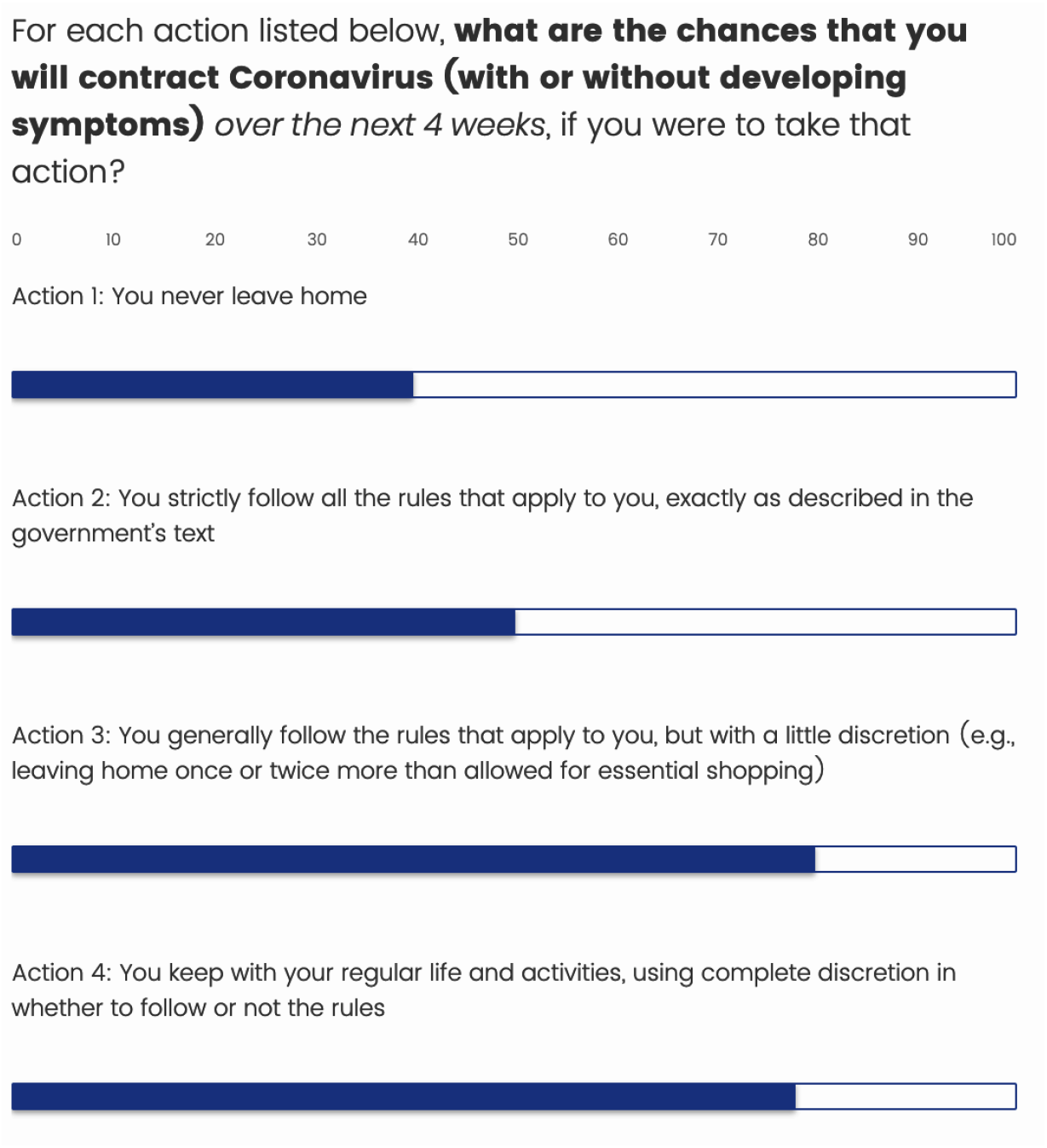
Example of Expectation Elicitation Question, Conditional on Alternative Compliance Behaviors

**Figure A11:**
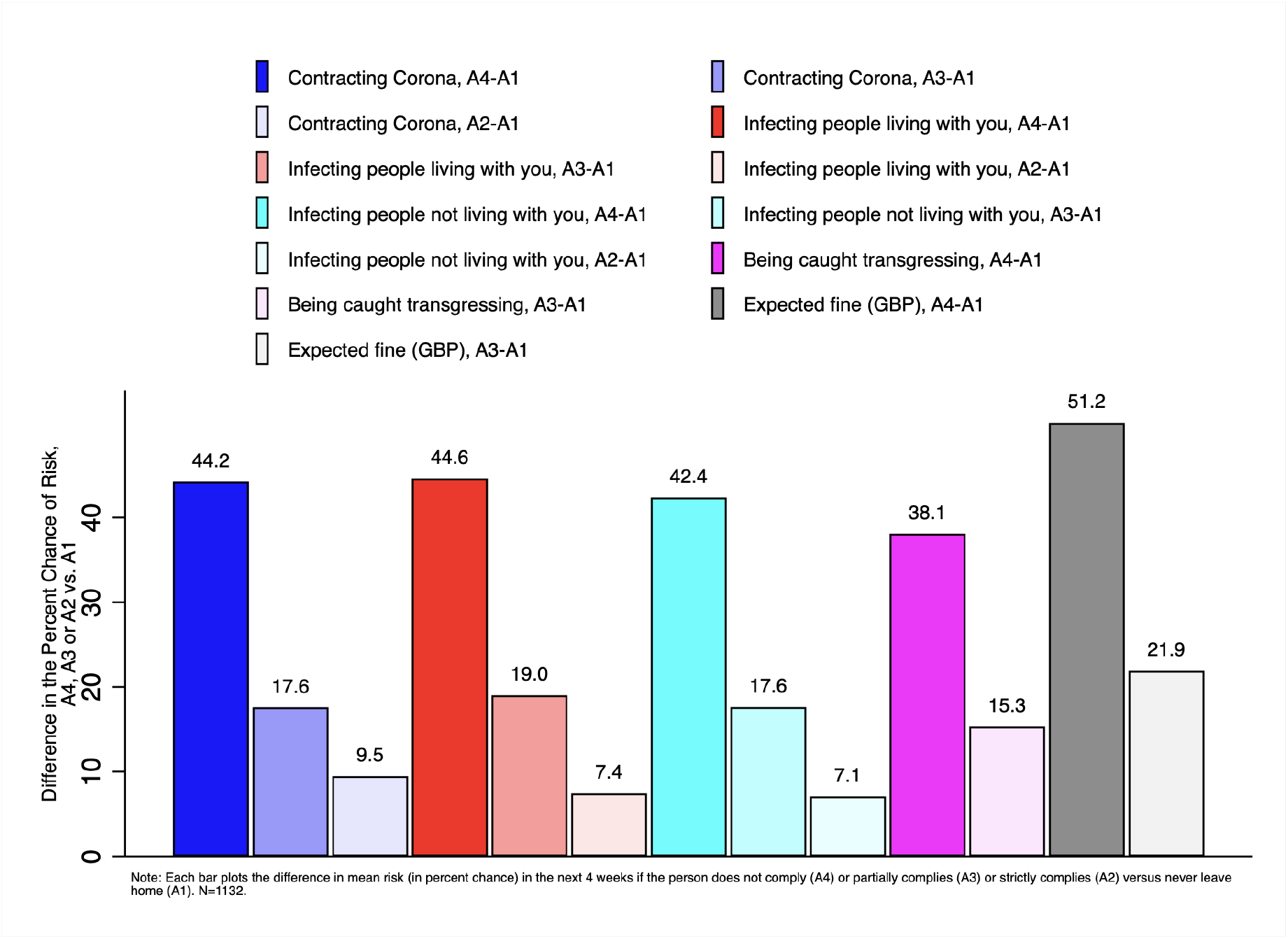
Perceived Risks to Non-Compliance and Partial Compliance

**Figure A12:**
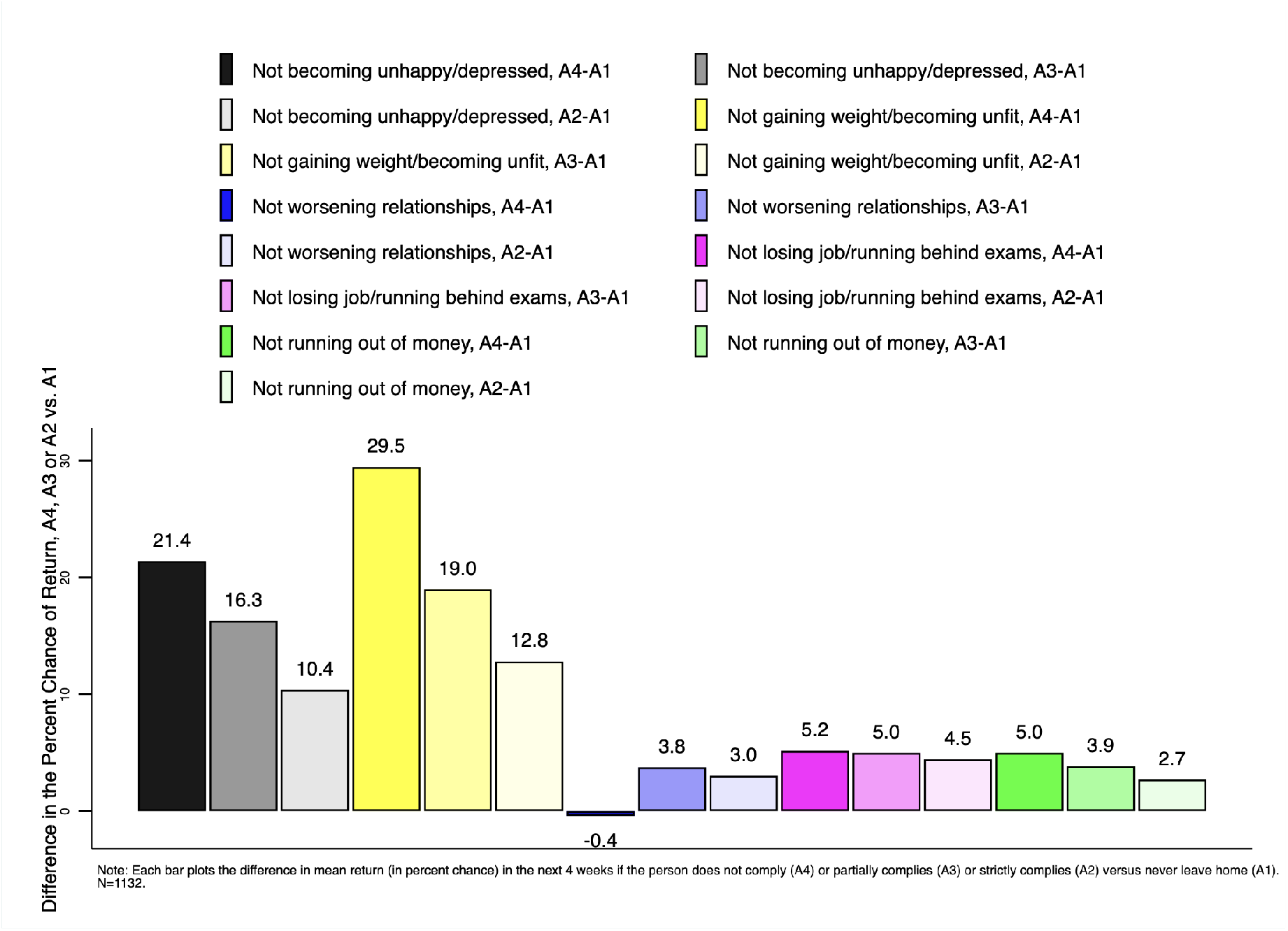
Perceived Returns to Non-Compliance and Partial Compliance

**Table A1:** Model With Homogeneous Utilities – LAD Estimates

| $\beta_k$ | Exp. Sign | Estimate |
| --- | --- | --- |
| **Risks** |  |  |
| $\beta_1$ (contracting Coronavirus) | - | 0.324 (0.401) |
| $\beta_2$ (unable to find ICU with acute COVID) | - | -1.987 (1.599) |
| $\beta_3$ (passing away for COVID) | - | -1.119 (0.694) |
| $\beta_4$ (infecting people living with) | - | -0.999 (0.352)*** |
| $\beta_5$ (infecting people not living with) | - | -1.592 (0.367)*** |
| $\beta_6$ (being caught transgressing) | - | -2.754 (0.362)*** |
| $\beta_7$ (expected fine) | - | -0.300 (0.130)** |
| **Benefits** |  |  |
| $\beta_8$ (not unhappy/depressed) | + | 1.933 (0.287)*** |
| $\beta_9$ (not unfit/gain weight) | + | 0.639 (0.286)** |
| $\beta_{10}$ (no worse relationships) | + | 0.899 (0.291)*** |
| $\beta_{11}$ (not losing job) | + | 1.382 (0.349)*** |
| $\beta_{12}$ (not running behind with exams) | + | 0.615 (0.954) |
| $\beta_{13}$ (not running out of money) | + | -0.850 (0.364)** |
| Constant |  | 0.776 (0.100)*** |
| Observations |  | 1,132 |

Note: k=1: subjective probability of contracting Coronavirus; k=2: subjective probability of not finding space in ICU after contracting Corona & getting COVID-19 with severe symptoms; k=3: subjective probability of dying after contracting Coronavirus; k=4: subjective probability of infecting someone living with you; k=5: subjective probability of infecting someone not living with you; k=6: subjective probability of being caught transgressing; k=7: expected fine (weighted by subjective probability of being caught transgressing); k=8: subjective probability of not becoming unhappy/depressed; k=9: subjective probability of not gaining weight/becoming unfit; k=10: subjective probability of relationship not deteriorating; k=11: subjective probability of not losing job; k=12: subjective probability of not falling behind with exams; k=13: subjective probability of not running out of money. Standard errors in parentheses.
\*\*\*: $p < 0.01$ ; \*\*: $p < 0.05$ ; \*: $p < 0.1$ .

**Table A2:** Model With Heterogeneous Utilities by Gender, Vulnerability, and COVID-19 Experience – LAD Estimates

| $\beta_k$ | Female<br>Non-Vulnerable<br>No COVID Exp. | Female<br>Non-Vulnerable<br>w/ COVID Exp. | Female<br>Vulnerable<br>No COVID Exp. | Female<br>Vulnerable<br>w/ COVID Exp. | Male<br>Non-Vulnerable<br>No COVID Exp. | Male<br>Non-Vulnerable<br>w/ COVID Exp. | Male<br>Vulnerable<br>No COVID Exp. | Male<br>Vulnerable<br>w/ COVID Exp. |
| --- | --- | --- | --- | --- | --- | --- | --- | --- |
| **Risks** |  |  |  |  |  |  |  |  |
| $\beta_1$ (contracting Coronavirus) | -0.851<br>(1.012) | 0.407<br>(0.608) | -1.651<br>(1.932) | -0.394<br>(1.743) | -0.088<br>(0.937) | 1.169<br>(0.727) | -0.889<br>(1.898) | 0.369<br>(1.792) |
| $\beta_2$ (unable to find ICU with acute COVID) | -0.207<br>(3.658) | -2.853<br>(2.507) | 6.749<br>(4.765) | 4.104<br>(3.799) | -1.843<br>(4.001) | -4.489<br>(3.438) | 5.114<br>(5.355) | 2.468<br>(4.827) |
| $\beta_3$ (passing away for COVID) | 0.075<br>(1.737) | -0.666<br>(1.121) | -0.922<br>(2.637) | -1.662<br>(2.245) | 0.340<br>(1.717) | -0.399<br>(1.312) | -0.657<br>(2.743) | -1.397<br>(2.478) |
| $\beta_4$ (infecting people living with) | -0.237<br>(0.802) | -0.704<br>(0.549) | -5.129***<br>(1.348) | -5.597***<br>(1.226) | -0.364<br>(0.789) | -0.831<br>(0.637) | -5.257***<br>(1.452) | -5.724***<br>(1.385) |
| $\beta_5$ (infecting people not living with) | -2.899***<br>(0.889) | -1.981***<br>(0.548) | 2.309*<br>(1.400) | 3.227***<br>(1.187) | -2.610***<br>(0.835) | -1.693**<br>(0.715) | 2.598*<br>(1.459) | 3.516***<br>(1.372) |
| $\beta_6$ (being caught transgressing) | -3.243***<br>(0.842) | -2.642***<br>(0.553) | -5.402***<br>(1.359) | -4.802***<br>(1.232) | -3.636***<br>(0.827) | -3.035***<br>(0.672) | -5.795***<br>(1.398) | -5.195***<br>(1.339) |
| $\beta_7$ (expected fine) | 0.00003<br>(0.003) | -0.003*<br>(0.002) | -0.009*<br>(0.005) | -0.013***<br>(0.005) | 0.002<br>(0.003) | -0.001<br>(0.003) | -0.008<br>(0.005) | -0.011**<br>(0.005) |
| **Benefits** |  |  |  |  |  |  |  |  |
| $\beta_8$ (not unhappy/depressed) | 2.675***<br>(0.644) | 2.067***<br>(0.451) | 1.093<br>(1.098) | 0.475<br>(0.988) | 2.446***<br>(0.657) | 1.838***<br>(0.506) | 0.854<br>(1.163) | 0.247<br>(1.075) |
| $\beta_9$ (not unfit/gain weight) | 0.198<br>(0.642) | 0.116<br>(0.446) | -0.287<br>(1.162) | -0.369<br>(1.153) | 0.863<br>(0.627) | 0.781<br>(0.493) | 0.378<br>(1.249) | 0.296<br>(1.266) |
| $\beta_{10}$ (no worse relationships) | 1.022<br>(0.709) | 0.795*<br>(0.462) | 2.936**<br>(1.247) | 2.709**<br>(1.094) | 0.283<br>(0.673) | 0.056<br>(0.496) | 2.197*<br>(1.237) | 1.969*<br>(1.119) |
| $\beta_{11}$ (not losing job) | 4.591***<br>(0.983) | 1.689***<br>(0.552) | 1.415<br>(2.231) | -1.486<br>(2.133) | 2.931***<br>(0.848) | 0.030<br>(0.566) | -0.245<br>(2.240) | -3.146<br>(2.203) |
| $\beta_{12}$ (not running behind with exams) | -2.283<br>(3.209) | -2.271<br>(1.694) | -1.354<br>(7.268) | -1.342<br>(6.381) | 3.208<br>(3.717) | 3.220**<br>(1.395) | 4.137<br>(7.826) | 4.149<br>(6.686) |
| $\beta_{13}$ (not running out of £) | -3.038***<br>(0.974) | -0.869<br>(0.561) | -4.211**<br>(2.033) | -2.043<br>(1.914) | -2.041**<br>(0.849) | 0.127<br>(0.634) | -3.214<br>(2.082) | -1.046<br>(2.044) |
| Constant | 0.854***<br>(0.105) |  |  |  |  |  |  |  |
| **Observations** | 1,127 |  |  |  |  |  |  |  |
| **Pseudo R<sup>2</sup>** | 0.083 |  |  |  |  |  |  |  |

Note: k=1: subjective probability of contracting Coronavirus; k=2: subjective probability of not finding space in ICU after contracting Corona & getting COVID-19 with severe symptoms; k=3: subjective probability of dying after contracting Coronavirus; k=4: subjective probability of infecting someone living with you; k=5: subj prob of infecting someone not living with you; k=6: subj prob of being caught transgressing; k=7: expected fine (weighted by subjective probability of being caught transgressing); k=8: subjective probability of not becoming unhappy/depressed; k=9: subjective probability of not gaining weight/becoming unfit; k=10: subjective probability of relationship not deteriorating; k=11: subjective probability of not losing job; k=12: subjective probability of not running behind with exams; k=13: subjective probability of not running out of money. Standard errors in parentheses.
\*\*\*: $p < 0.01$ ; \*\*: $p < 0.05$ ; \*: $p < 0.1$ .

**Figure A13:**
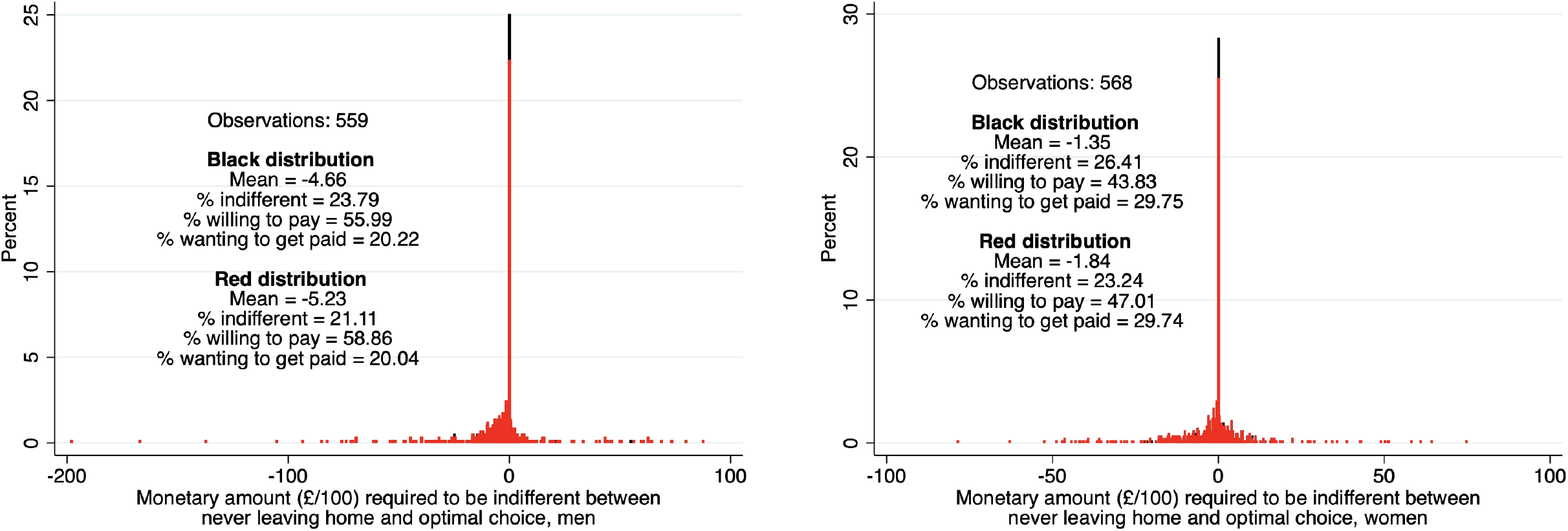
Distribution of the Monetary Amount Required to be Indifferent Between Never Leaving Home and Optimal Choice, by Gender

**Figure A14:**
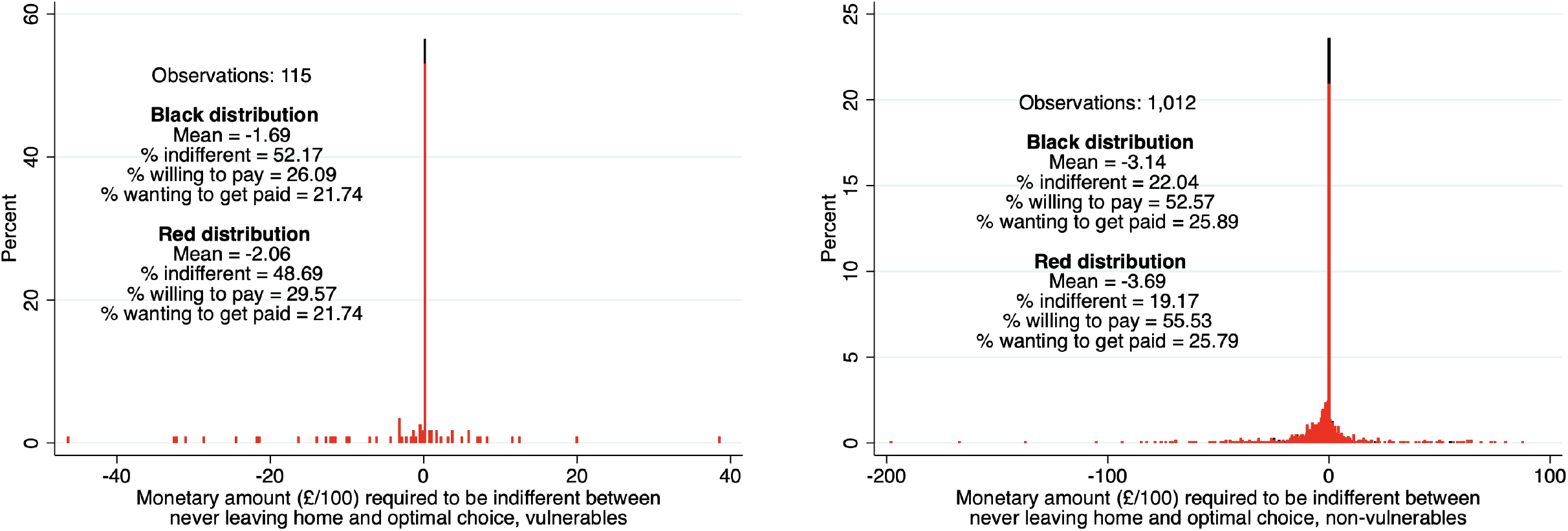
Distribution of the Monetary Amount Required to be Indifferent Between Never Leaving Home and Optimal Choice, by Vulnerability Status

**Figure A15:**
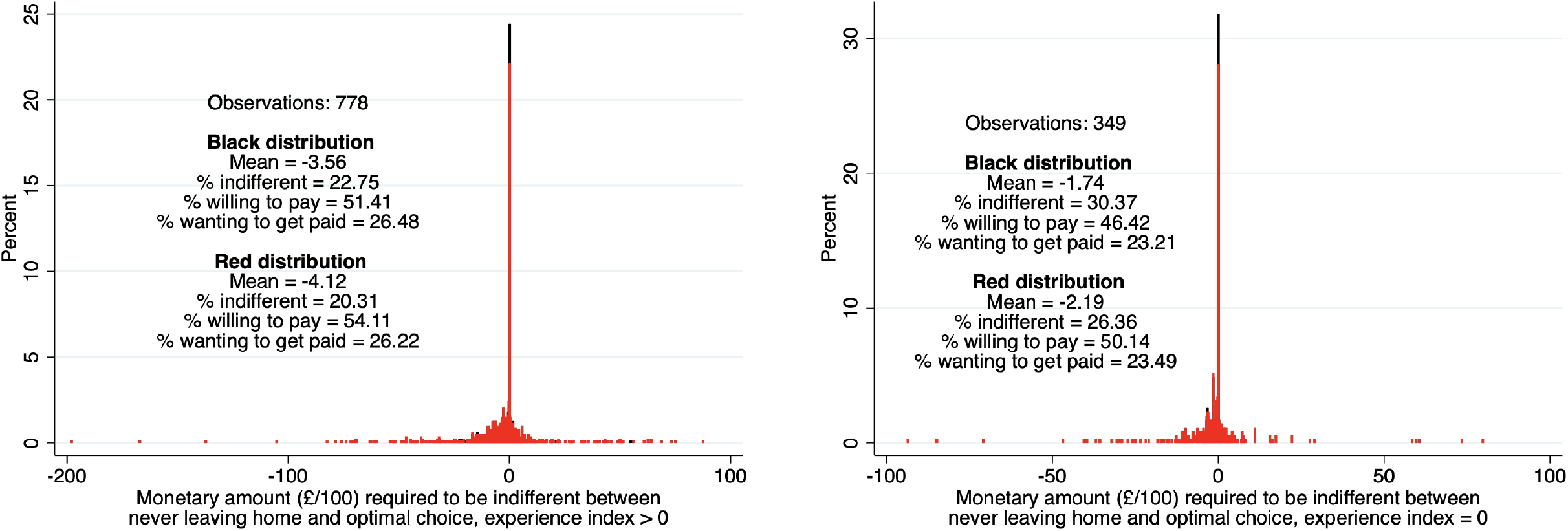
Distribution of the Monetary Amount Required to be Indifferent Between Never Leaving Home and Optimal Choice, by COVID- 19 Experience

## Footnotes

1 Notable exceptions are Sloan and Platt (2011), Delavande (2008), Dupas (2011), Miller, De Paula and Valente (2020), Biroli et al. (2022), Bhalotra et al. (2020) and Arni et al. (2021).

2 The “Cummings scandal” refers to a series of events involving Boris Johnson’s senior adviser Dominic Cummings, during the first UK lockdown. The events include at least one trip by car from London to Durham that Cummings made with his wife and son to reach his parents and sister, while presenting symptoms consistent with a COVID-19 infection, thus allegedly violating multiple lockdown rules.

3 Stated choices include forced choices, choice rankings, choice ratings, choice intentions, choice probabilities, and related measures. The term “stated preferences” refers to the fact that respondents state what choices they would make in hypothetical choice situations specified in a survey or experiment, or predict what choices they will make in the future. It is used in contrast to the term “revealed preferences” (RP), which refers to people’s actual choices in real-world situations.

4 For introductory treatments of discrete choice modelling with SP data, alone or combined with RP data, see for instance Train (2009), Ben-Akiva, McFadden and Train (2019), Ben-Akiva et al. (1994), and references therein. The strengths and weaknesses of SP and RP data and related approaches are well known. As put by Train (2009), pp. 152-153, “*Revealed-preference data have the advantage that they reflect actual choices.* (…) *However, such data are limited to the choice situations and attributes of alternatives that currently exist or have existed historically.* (…) *Even for choice situations that currently exist, there may be insufficient variation in relevant factors to allow estimation with revealed- preference data.* (…) *The advantage of stated-preference data is that the experiments can be designed to contain as much variation in each attribute as the researcher thinks is appropriate.* (…) *The limitations of stated-preference data are obvious: what people say they will do is often not the same as what they actually do.*”

5 As such, they tend to share the same strengths and weaknesses as other types of SP data relative to RP data. At the same time, subjective choice probabilities can have distinctive advantages over more traditional forms of SP data. Traditional elicitation of stated choices has asked respondents to select from a given list of options the alternative that they prefer, or expect to choose, in the future or in some hypothetical scenario. Manski (1990) shows that responses to this kind of questions have limited information content. Intuitively, observing that a respondent selects a certain option only reveals that he or she assigns a subjective probability higher than a certain threshold to the event of choosing that option, where the threshold depends on the loss function used by the respondent in forming his or her prediction. Unfortunately, the respondent’s subjective probability, threshold, and loss function are typically unobserved. Thus, Manski (1990, 1999) recommends directly eliciting respondents’ subjective probabilities over choice alternatives. See Juster (1966), Stinebrickner and Stinebrickner (2014), and Manski (2016) for additional theoretical arguments, empirical evidence, and in-depth discussions about the greater informativeness of subjective choice probabilities over stated intention data.

6 The first form of uncertainty is called resolvable, as it is resolved by the time of an actual choice. The latter form of uncertainty is called unresolvable, as it is present at the time of an actual choice. See Manski (1999, 2004), Blass, Lach and Manski (2010), Stinebrickner and Stinebrickner (2014), and others, discussion and applications.

7 The analysis in the first part of the paper does not amount to a discrete choice experiment, since we do not manipulate the characteristics of the choice alternatives or choice environment via hypothetical scenarios or vignettes.

8 We use measurement and econometric tools developed in a growing economic literature on survey expectations, reviewed from various perspectives by Manski (2004, 2023), Attanasio (2009), Attanasio, Almas and Jervis (2020), Delavande (2014, 2023), Hurd (2009), Hudomiet, Hurd and Rohwedder (2023), Giustinelli and Manski (2018), Giustinelli (2022, 2023), Bruine de Bruin et al. (2023), Fuster and Zafar (2023), and Koşar and O’Dea (2023), among others.

9 We review the restrictions in detail in Section 2.

10 Shielding was applied to those citizens who were considered vulnerables due to their age or health conditions; these individuals were expected not to leave their home for 12 weeks. Self-isolation for one week (two weeks) was required of individuals (households) testing positive to the Coronavirus or manifesting symptoms consistent with COVID-19.

11 The police issued more than 117,000 fixed penalty notices for breaches of Coronavirus restrictions up to 20 June 2021, see https://news.npcc.police.uk/releases/ update-on-coronavirus-fpns-issued-by-forces-in-england-and-wales-and-the-payment-of-fpns.

12 https://www.gov.uk/government/news/new-package-to-support-and-enforce-self-isolation.

13 These examples illustrate a well-known identification problem in the analysis of choice behaviors under uncertainty, whereby different combinations of expectations and preferences can be observationally equivalent choice-wise. They also illustrate the importance of measuring expectations (or preferences) in addition to choices, to reduce the risk of wrong inferences such as attributing a certain behavior to preferences instead of expectations, or viceversa. For more examples and formal treatments of these issues, see Manski (2004), Delavande (2008), Arcidiacono, Hotz and Kang (2012), van der Klaauw (2012), Zafar (2013), Stinebrickner and Stinebrickner (2014), Giustinelli (2016), and Koşar and O’Dea (2023), among others.

14 Mainly the vulnerables, since the quarantine period applied to the self-isolating was shorter than 4 weeks (unless of repeated infections within a household).

15 As further discussed below, we see this conceptualization as striking a balance between realism and tractability. Wright, Steptoe and Fancourt (2022) perform a latent class analysis of patterns of compliance behavior and assign respondents to four classes: full compliers, frequent compliers, occasional compliers, and household mixers – which provides support to our modelling choice.

16 Galasso et al. (2020) show large gender differences in COVID-19-related beliefs and behaviors, on the basis of survey evidence from eight OECD countries at the initial stages of the COVID-19 lockdown.

17 Finland and Italy were the first to declare national lockdowns, which became effective on March 8th and 9th respectively. By March 23rd, the vast majority of EU member countries were already in lockdown, with the exception of Cyprus, Romania, and Hungary, which entered lockdown between March 24th and March 28th. A minority of countries, including Latvia, Luxembourg, Malta, Slovakia, Slovenia, and Sweden, avoided extreme lockdown measures, at least in the first wave of the pandemic.

18 Support with care, medicine supplies and daily living was provided by the government, via the local GP practices and volunteer groups.

19 Wright, Steptoe and Fancourt (2022) elicit self-reported compliance along six dimensions (hand-washing, mask- wearing, social distancing, etc.) in a large online survey in November-December 2020 and find that most individuals reported similar levels of compliance across the six behaviour measures, providing further support to our parsimonious modelling choice.

20 Additive separability rules out interactions effects between outcomes assumed separable. Nevertheless, individual elements of *θ⃗* can be joint events, as it is indeed the case for certain outcomes in our empirical SEU specification, described below. Our use of additive separability is partly motivated by tractability in data collection, since measuring subjective probabilities of separable binary outcomes only requires elicitation of one marginal probability per outcome and choice alternative, i.e., *P_ij_*(*b_k_* = 1) for all (k, j) pairs, instead of joint belief distributions. In specific cases, because we elicited the probability of certain outcomes conditional on others (e.g., the likelihood of developing COVID-19 symptoms of varying intensity conditional on contracting the Coronavirus), we can construct probabilities for joint events using probability laws. An implication of additive separability is that the decision maker is risk-neutral with respect to continuous outcomes, in our case the fine an individual would get if caught transgressing. Relaxing this assumption would require eliciting multiple points on the respondent’s subjective fine distribution, instead of just the expected value.

21 Multiplicative separability between probabilities and utilities is a standard feature of canonical SEU (Savage, 1954). It rules out the possibility that a person subjective probability of an event depends on the (dis)utility the person assigns to the same event, as in models of utility-based or motivated beliefs (e.g., Brunnermeier (2005) or Benabou and Tirole (2016)). For instance, multiplicative separability would be violated if the decision maker’s subjective probability of contracting the Coronavirus were to depend on her disutility of that happening. A simple way to partially relax this assumption – and also that of outcome separability – would be to allow the utility of an outcome, say k^^^, to depend on the decision maker’s subjective probability of another outcome, say k^̌^, hypothesized to be related to the first. This is equivalent to introduce heterogeneity in the utility of k^^^ with respect to the person’s expectations for k^̌^, which would be simple to do in our setting since the latter are directly elicited in the survey.

22 Compared to the formulation in (2), in the econometric implementation of Section 4 we will limit the amount of heterogeneity in utility parameters across individuals, while still allowing for unrestricted heterogeneity of expectations across individuals. Parameter homogeneity across choice alternatives is customary in empirical models of discrete choice, but it could be relaxed with richer expectations data.

23 Note that the probability of being caught, *Pij* (caught), is only defined for non-compliance (actions A3 and A4).

24 Note that the expected fine is not action-specific.

25 Recall that the utility parameters do not vary across choice alternatives.

26 Detailed information about Prolific Academic can be found at https://www.prolific.co/.

27 For example, see https://www.prolific.co/prolific-vs-mturk/ for a comparison with M-Turk.

28 Information on how the demographic subgroups used for representative samples are created can be found on Prolific’s webpage: https://researcher-help.prolific.co/hc/en-gb/articles/ 360019238413-Representative-Samples-FAQ-limited-release-.

29 These are the rules we described in Subsection 2.1.

30 We pay respondents for taking the survey (according to the Prolific Academic regulations), but we don’t incentivize respondents for accurate reporting to specific questions. It is unusual in the survey subjective expectations literature to incentivize respondents for accurate reporting. For example, Botelho and Pinto (2004)’s elicitation procedure used a scoring rule to provide financial incentives for accurate reporting to a random subsample of respondents. The authors find no significant effects on the accuracy of respondents’ beliefs. Wiswall and Zafar (2015) do not incentivize their respondents on the ground that scoring rules tend to induce biased responses when responses are not risk neutral.

31 For instance, in an online survey on a nationally representative sample of the Dutch population, Bruine de Bruin and Carman (2018) found that elicitation of percent-chance probabilities using clickable sliders significantly reduced responses of 50 percent relative to a more traditional open-ended mode, without affecting the predictive validity of responses and survey satisfaction of respondents.

32 For an early discussion of rounding of numerical survey expectations, see Dominitz and Manski (1997); for a recent review and extensive analysis in the U.S. Health and Retirement Study, see Giustinelli, Manski and Molinari (2022).

33 A detailed description of the workings of the NHS TTS can be found at https://www.gov.uk/guidance/ nhs-test-and-trace-how-it-works. See also Fetzer and Graeber (2021).

34 Before seeing the Cummings screen, all respondents (including treated ones) were re-asked: (i) the battery on weekly habits during the lockdown from Section D of the baseline; (ii) the probability of contracting the Coronavirus with or without symptoms in the following 4 weeks from Section E of the baseline; (iii) basic demographic questions such as age, gender, and place of residence from Sections A and F of the baseline. We also asked every respondent whether they had seen the news about Dominic Cummings.

35 “*Do you think that Dominic Cummings broke the lockdown rules?* ” Possible answers included: (1) *Yes, but I was not aware that the government advice included an exception related to care of small children*; (2) *Yes, and I was aware that the government advice included an exception related to care of small children*; (3) *No, and I was aware that the government advice included an exception related to care of small children*; (4) *No, but I was not aware that the government advice included an exception related to care of small children*; (5) *Unsure*.

36 We have used 21 questions to construct this index, such as “Have you experienced any of the following symptoms since the beginning of February?”, “Have you been hospitalised since the beginning of February?”, “Do you personally know anyone who has tested positive for coronavirus?”. We have assigned value 1 to all the affirmative questions (indicating some experience with the coronavirus), and summed them up; we have then divided this sum by the total possible number of affirmative answers (i.e., by 21).

37 We have used 59 questions to construct this index, such as “Have you heard the expression “flatten the curve”?”, “Which of the following behaviours are effective at protecting you against coronavirus?”, “How many reported deaths from coronavirus disease (COVID-19) are there in the UK?”, “What are you allowed to do during the lockdown?”. We have assigned value 1 to all the affirmative questions (indicating some knowledge of the coronavirus), and summed them up; we have then divided this sum by the total possible number of correct answers (i.e., by 59).

38 Ganslmeier, Van Parys and Vlandas (2022) find a self-reported level of non-compliance for the same week of May 2020 of approximately 5%, using a large online sample of more than 100,000 UK individuals compiled by YouGov. Their variable of interest was based on the survey question “Which comes closer to describing you?”. Answer “’I will probably follow the advice of the government even if I don’t agree with it or find it pointless” was coded as compliance; while answer “I will probably do my own thing, regardless of government advice” was coded as non-compliance. Wu, Font and McCamley (2022) survey a MTurk-based sample and find that residents in England were more health-conscious and more altruistic during the first national lockdown.

39 https://ourworldindata.org/covid-google-mobility-trends

40 Ganslmeier, Van Parys and Vlandas (2022) find a high concordance between self-reported compliance and self-reported actual social distancing behaviors.

41 Because we are limited in the number of dimensions of heterogeneity we can allow for in the utility parameters of the model we estimate in Section 4 due to sample size considerations, we decided to focus on a few dimensions of heterogeneity that we deem particularly relevant, that is, vulnerability status, gender, and COVID-19 experience. Vulnerability status is the most relevant dimension in this context since the strictness of the lockdown rules differed between vulnerables and non-vulnerables; moreover, it should capture heterogeneity in characteristics based on which vulnerability is defined such as age and health. Gender is a highly relevant dimension since the COVID-19 literature has repeatedly documented the existence of significant systematic differences in risk perceptions, compliance behavior, and mortality across genders. Lastly, we consider respondents’ prior experience with COVID-19 for at least two reasons. First, prior experience with the Coronavirus and COVID-19 may be an important initial condition, as we fielded our baseline survey at the beginning of May 2020, a few months after the pandemic’s breakout and over a month inside the first lockdown. Second, the subjective expectations literature has found repeatedly that personal experiences are important drivers of belief formation and decisions under uncertainty. Nonetheless, we have also explored additional dimensions of heterogeneity in choice probabilities and underlying subjective expectations and preferences, including COVID-19 literacy, willingness to take risks, patience, age, and education, described in Table 1. Results are available upon request.

42 The percentage of vulnerables assigning the whole mass to A2 is 6.96%; hence, the percentage of vulnerables assigning the whole mass to A1 and/or A2 (excluding those giving corner responses) is 25.22%.

43 The complete list of behaviours is as follows: (1) Been outside (not in balcony/garden) for 15 minutes or more (2) Kept a distance of at least 2 meters to other people (when outside) (3) Avoided touching objects/surfaces (when outside) a DIY mask/face cover/snood (8) Met someone from another household (9) Went to work (10) Exercised outside (e.g. running) (11) Went sunbathing/suntanning (not in balcony/garden).

44 See Giustinelli and Shapiro (2023) for a similar exercise in the context of subjective working probabilities. See D’Haultfoeuille, Gaillac and Maurel (2021) and Crossley et al. (2021) for recent tests of rational expectations based on survey-elicited belief distributions over continuous variables and their application to subjective earnings expectations.

45 This set of questions was shown on a single screen and the answers were required to sum to 100 percent.

46 This was asked under scenarios A3 and A4 only.

47 https://www.gov.uk/government/collections/coronavirus-job-retention-scheme.

48 However, it should not be overlooked that standard deviations are large for all outcomes, revealing substantial heterogeneity around the mean values shown in the tables.

49 We will relax this assumption in the next subsection.

50 This is normal practice in econometric analysis of revealed preferences, dating back to McFadden (1973). Observing a single cross-section of discrete choices or behaviors, as common in micro data, does not generally suffice to recover individuals’ underlying preference distribution. To account for the fact that individuals with identical observable characteristics and choice environments are routinely observed to make distinct choices, an individual- and choice-specific random term is added to the utility specification to capture this unobserved heterogeneity, leading to the expressions *random utility model* or *random utility specification*. These terms refer to the fact that the decision maker’s utility is random from the viewpoint of the econometrician. Inference on individuals’ preferences typically requires assumptions on the distribution of unobserved heterogeneity. From the viewpoint of the decision maker, on the other hand, there is no randomness. The decision maker is typically assumed to know her utility function. Should her decision depend on some uncertain state or consequence, she will form expectations about the uncertainties and select the action or behavior that maximizes her SEU, as in (3). This econometric interpretation of revealed preference differs from the traditional psychological interpretation, whereby individuals are imagined to have a family of utility functions from which they draw each time they face a choice, thus viewing individual behavior as intrinsically probabilistic (see Thurstone (1927) and Luce and Suppes (1965)’s survey).

51 When the elicited choice probabilities have implied corner values of 0 or 1, we follow a common practice in the survey expectations literature and recode them to values just above 0/below 1.

52 To investigate the robustness of parameter estimates to our recoding of corner choice probabilities, we re-estimate (7) via least absolute deviations (LAD). Median regression is invariant to transformations that do not alter the ordering of values relative to the median. However, it requires that the unobserved υ*ij* are symmetrically distributed around zero conditional on the observables, implying a conditional median restriction on the unobservables.

53 Recall that the probability of outcome k = 2 is constructed as *Pi*(no ICU space|acute COVID, Corona) *× Pi*(acute COVID|Corona) *× Pij* (Corona) and the probability of outcome k = 3 is constructed as *Pi*(dying of COVID|Corona) *× Pij* (Corona).

54 While the large disutility associated with being caught transgressing might surprise at first instance, it can be rationalized with the political discourse which was reigning in the UK around the first lockdown, when experiences of shame, shaming and stigma dominated personal and public life, with “the government’s healthcare policies and rhetoric seemed to exacerbate experiences of shame, shaming and stigma, relying on a language and logic that intensified oppositional, antagonistic thinking, while dissimulating about its own responsibilities.” (Cooper, Dolezal and Rose, 2023).

55 Wright et al. (2022) analyze text data from 17,500 UK adults and find that, in November-December 2020, the main factors facilitating compliance were desires to reduce risk to oneself and one’s family and friends and to, a lesser extent, the general public. Wu, Font and McCamley (2022) also find that altruistic values played a consistently strong role to form behavioral intentions to comply with social distancing measures in the first lockdown (“[…] I want to help others” “[…] I care for people in my country”).

56 Keyworth et al. (2021) find that mental health was the most prevalent challenge reported by UK adults (reported by 41.4% of the sample) when adhering to COVID-19 related restrictions at the end of April 2020, the same time covered in our survey.

57 As mentioned in 3.5, since 1st of March 2020, the government had introduced the Coronavirus Job Retention Scheme. This should have lessened individuals’ concerns with regard to the possibility of running out of money.

58 Table A2 in the Supplementary Appendix reports LAD estimates for the same model with heterogeneous utilities by gender, vulnerability status, and prior experience with COVID-19.

59 The LAD estimates reported in Table A2 in the Supplementary Appendix are very similar, with a couple of exceptions: the disutility of contracting the coronavirus (β1) loses significance when estimated via LAD, while the disutility of infecting people not living with (β5) instead gains statistical significance when estimated via LAD.

60 Source: https://www.gov.uk/government/news/new-payment-for-people-self-isolating-in-highest-risk-areas.

61 Eligibility required that the person was employed or self-employed, could not work from home, and would lose income as a result of self-isolation. Moreover, eligible individuals could only apply if they were legally required to self-isolate because told so by the NHS Test and Trace Service, notified by the NHS COVID-19 App, or as the parent or guardian of a child who was told to self-isolate. This program was scheduled to run from September 28, 2020 until March 31, 2021, but was extended by the government through June 30, 2021.

62 The exact estimates depends on how we treat ties in choice probabilities. Figure 5 shows the distribution of the monetary amount making the individuals indifferent between their optimal choice and never leaving home, under two alternative ways of treating ties in choice probabilities. The black distribution breaks each observed tie in favor of low- index alternatives. Under this distribution, action *j* = 1 is selected as optimal for a larger fraction of respondents, leading to the higher observed spike at 0. The red distribution breaks each observed tie in favor of high-index alternatives. Reassuringly, the two distributions are very close to each other, indicating that results are not particularly sensitive to how ties in choice probabilities are dealt with.

63 Analogously, Blayac et al. (2022) use a discrete choice experiment (DCE) to study acceptance to the social distancing restrictions, and find that vulnerables are averse to the idea of monetary compensation to accept them.

64 The figures showing the empirical distributions of the amount needed to make individuals indifferent between their optimal choice and never leaving home disaggregated by respondent’s characteristics are provided in the Supplementary Appendix, Figures A13, A14 and A15.

65 Giustinelli and Shapiro (2023) call measures of this type “subjective *ex ante* treatment effects” (S*ea*TE) and investigate their properties in the context of the effect of health on the probability of working among older workers. See also Arcidiacono et al. (2020) for an application to the ex ante returns to college majors and occupations.

66 Of these, 26.91% reported being aware that the government advice included an exception related to care of small children.

67 Using a high-frequency large-scale online survey, Fancourt, Steptoe and Wright (2020) notice that trust in the government (but not in other dimensions) sharply fell soon after the Cummings episode in England (but not in the other nations belonging to the United Kingdom). The importance of political accountability and trust is also studied by Martinez-Bravo and Sanz (2023) in the Spain. Interestingly, Bird et al. (2023) find that, in Latin America, political trust predicted less compliance (more mobility), unlike in high-income countries.

68 Bargain and Aminjonov (2020) use a double difference approach around the time of lockdown announcements in Europe, and find that high-trust regions decrease their mobility related to non-necessary activities significantly more than low-trust regions.

